# Serum protein N-glycome patterns reveal alterations associated with endometrial cancer and its phenotypes of differentiation

**DOI:** 10.1101/2023.01.16.23284637

**Authors:** Zejian Zhang, Zhen Cao, Jinhui Wang, Zepeng Li, Tao Wang, Yang Xiang

## Abstract

**Background and aims:** Aberrant N-glycosylation and its involvement in pathogenesis have been reported in endometrial cancer (EC). Nevertheless, the serum N-glycomic signature of EC remains unknown. Here, we investigated serum N-glycome patterns of EC to identify candidate biomarkers.

**Materials and methods:** This study enrolled 34 untreated EC patients and 34 matched healthy controls (HC) from Peking Union Medical College Hospital. State-of-the-art MS-based methods were employed for N-glycans profiling. Multivariate and univariate statistical analyses were used to identify discriminative N-glycans driving classification. Receiver operating characteristic analyses were performed to evaluate classification accuracy.

**Results:** EC patients displayed distinct differences in serum N-glycome and had abnormal high-mannose and hybrid-type N-glycans, fucosylation, galactosylation, and linkage□specific sialylation compared with HC. The glycan panel built with the four most discriminative and biologically important derived N-glycan traits could accurately identify EC (random forest model, the area under the curve [AUC]=0.993 [95%CI 0.955-1]). The performance was validated by two other models. Total hybrid-type N-glycans significantly associated with the differentiation types of EC could effectively stratify EC into well- or poorly-differentiated subgroups (AUC>0.8).

**Conclusion:** This study to the best of our knowledge provides the initial evidence supporting the utility of serum N-glycomic signature as potential markers for the diagnosis and phenotyping of EC.

## 1. Introduction

Endometrial cancer (EC) is the most common gynecologic cancer. It is the sixth most common malignancy in women, with 417 000 estimated new cases and 97 370 estimated new deaths worldwide in 2020 [1]. Its overall incidence is rising globally and has increased by 132% over the past three decades. More women than ever are dying from EC [2].

Patients with advanced EC have a high mortality rate, with a 5-year survival rate of less than 20%. Early diagnosis and timely treatment can improve the 5-year survival rate to over 90% [3], emphasizing the importance of timely and accurate diagnosis. The main clinical presentation of EC is irregular bleeding. This is usually post-menopausal but may also be heavy, prolonged, or intermenstrual bleeding (around 15-20% of EC cases are diagnosed before menopause). However, such symptoms are so common that only a small percentage of cases are caused by EC [4, 5]. Incorrectly attributing the symptoms to benign etiology will lead to a delayed diagnosis of EC. Except for the clinical symptoms, the preoperative diagnosis of EC mainly relies on evaluating the results of imaging (e.g., transvaginal ultrasound and hysteroscopy) and endometrial tissue sampling [2]. Nevertheless, imaging methods suffer from insufficient accuracy [6]. While, endometrial tissue sampling is an invasive test and may result in anxiety, bleeding, infection, or uterine perforation [7]. As a result, the incorporation of non-invasive diagnostic biomarkers into routine diagnostic pathways is vitally important, which will identify suspected cases for further invasive tests to confirm the diagnosis and reassure low-risk women. However, due to the lack of specificity or sensitivity, the routinely used serological biomarkers in gynecological oncology such as carbohydrate antigen 125 (CA125), carbohydrate antigen 15-3 (CA15-3), and carbohydrate antigen 19-9 (CA19-9) have limited diagnostic value for EC [8, 9]. Thus, exploring new effective and non-invasive biomarkers for the timely diagnosis of EC is necessary and urgent.

Commonly, EC is classified into estrogen-dependent (type I) and non-estrogen-dependent (type II) types based on clinical and hormonal characteristics. Type I EC, which comprises primarily endometrioid carcinoma and accounts for approximately 85% of cases, is typically well-differentiated and low-grade and usually associated with a good prognosis. While type II is more likely to be high-grade and more aggressive, including histotypes such as serous, clear cell, squamous carcinoma, and carcinosarcoma. Type II tumors are always associated with a poor prognosis and a high risk of metastasis and relapse [10, 11]. Therefore, phenotyping, staging, and grading of EC are very important for the evaluation of patients’ prognoses. At present, the classification is obtained based on post-operative pathology, which is low-throughput, time-consuming, and only known after surgery [2, 12]. These limitations sometimes affect the proper management of patients. The discovery of more convenient and pre-operative approaches for the stratification/prognosis of EC has become a high priority.

Glycosylation is the most prevalent and structurally diverse post-translational modification. N-linked glycosylation occurs at Asn residues and N-glycans ‘decorations’ attached to various proteins have essential roles in many vital biological processes and pathological mechanisms. Aberrant N-glycans have been found to be associated with the pathogenesis of many types of cancer including EC [13]. Recently, Lin *et al*. found that IgG N-glycosylation was significantly correlated with EC prognostic risk factors [14]. Differences in N-glycans were also observed between tissues of EC tumors and normal endometrium, or between primary tumors with lymph node metastasis (LNM) and those without [15], further confirming the involvement of N-glycans in EC development and progression. Serological glycomic profiling is a burgeoning non-invasive method for the discovery of candidate biomarkers for a variety of diseases, especially cancers [16-19]. It has been reported that in a given physiological state, human serum glycoforms are stable and reproducible, but dramatical alterations in glycan profiles can occur in response to cancer states (the host responds to tumors or tumors secrete certain aberrant glycoproteins), demonstrating biomarker potential of the blood-based glycans [20]. However, at current, little is known regarding N-glycosylation in serum at the global level in EC.

Here, using a high-sensitive matrix-assisted laser desorption/ionization time-of-flight mass spectrometry (MALDI-TOF-MS) approach, we for the first time focused on the qualitative and quantitative alterations of serum N-glycome between EC patients and matched healthy controls (HC) and the associations of serum glycans with clinical features (phenotypes) of EC, in an attempt to characterize the serum N-glycomic signature of EC and investigate effective and non-invasive glycan biomarkers for EC diagnosis and stratification/prognosis.

## 2. Methods

### 2.1 Study population and sample collection

In this study, we collected 34 eligible untreated patients diagnosed with EC. Besides, 34 age- and sex-matched healthy women who had no history of tumors, precancerous lesions, autoimmune diseases, and other systematic diseases were included in the control group. EC patients enrolled in this study should meet the following criteria: (1) over 18 years old; and (2) diagnosed with EC (including type I and type II) and histologically confirmed by surgery. Exclusion criteria were as follows: (1) less than 18 years old; (2) had received chemotherapy or radiotherapy or had undergone surgery; (3) recurrent EC; (4) with other malignant tumors besides EC; and (5) with detectable infections. All serum samples of the participants were collected at Peking Union Medical College Hospital between November 2020 and November 2021 and were stored at −80 □ until N-glycome profiling. The regional ethics committee of Peking Union Medical College Hospital approved this study (No. S-K2059). All patients and healthy volunteers gave written informed consent. The study complied with the ethical standards for human experimentation in the latest version of the Declaration of Helsinki.

### 2.2 Enzymatical release of N□glycans from serum

Serum samples were treated with PNGase F to release N-glycans from serum glycoproteins according to our previous reports [17, 20]. In short, for each sample, 10 μL of 2% SDS and 5 μL of serum were thoroughly mixed and incubated for 15 min at 65 °C. Then the releasing buffer including 1 U PNGase F was added, followed by incubation at 37 °C for 12 hours [17, 20].

### 2.3 Glycan derivatization and purification

Released N-glycans were derivatized employing the linkage-specific sialic acid stabilization method allowing stabilization and mass-based differentiation of the sialic acid isomers (α2,3- and α2,6-linked sialic acids), as described in detail elsewhere [17, 20]. Briefly, the mixture of 1 μL of the released glycans and 20□μL of derivatization reagent containing 250□mM EDC and 250□mM HOBt in ethanol was incubated at 37□°C for 60□min, followed by adding acetonitrile (ACN) and further incubation at −20□°C. Thereafter, the derivatized N-glycans were enriched and purified by hydrophilic interaction liquid chromatography solid-phase extraction (HILIC-SPE) [21, 22].

### 2.4 MALDI-TOF-MS measurement of N-glycome

MALDI-TOF-MS in reflection positive-ion mode was used for the serum N-glycans profiling [17]. 1□μL of glycan samples was mixed with 1 μL of matrix containing 5□mg/mL super-DHB with 1□mM NaOH in 50% ACN on the AnchorChip target plate (Bruker Daltonics, Bremen, Germany) and the mixture was dried by air for 2 h. The N-glycomes were detected using a rapifleXtreme MALDI-TOF mass spectrometer with a Smartbeam-3D laser, controlled by flexControl 4.0 (Bruker Daltonics). A peptide calibration standard (Bruker Daltonics) was used for the calibration of the instrument. The mass range was set from *m/z* 1000 to *m/z* 5000. Laser shots were accumulated 5000 times for each spectrum. We chose an automatic acquisition mode and random walk pattern at a laser frequency of 5000□Hz for the acquisition of sample spectra [17].

### 2.5 Raw MALDI-TOF-MS data processing

The detailed procedure for the raw MALDI-TOF-MS data processing is described previously [17, 20]. In short, the raw data was first baseline-subtracted and smoothed and then transformed to .xy files by flexAnalysis 4.0 (Bruker Daltonics). Re-calibration of the .xy files was conducted using selected high-intensity glycan signals (Supplementary Table S1) as calibrants through the software MassyTools (version 0.1.8.1.2) [23]. The glyco-Peakfinder tool of GlycoWorkbench (version 1.1.3480) [24, 25] and previously confirmed glycan compositions [20, 21] were used for the manual assignment and confirmation of mono-isotopic peaks to N-glycan compositions. An N-glycan composition list including 109 N-glycan structures used for the subsequent targeted glycan extraction was generated applying the criteria of mono-isotopic peaks with good isotopic patterns, signal to noise (S/N) > six, and relative intensity > 0.1%. The peak areas (background-corrected) of the putative N-glycans together with quality parameters were then extracted into excel using the glycan composition list through MassyTools. Thereafter, curation was done for the extracted glycans by applying criteria of S/N > nine, ppm error < 20, QC score < 25%, and minimum percentage (> 50%) of presence in all spectra of HC, EC, or quality control serum samples (replications of a standard serum sample randomly distributed on the plates). Finally, 79 N-glycans out of the 109 passed the criteria for the next statistical analysis (Supplementary Table S1) and the sum of the areas of the remaining 79 N-glycans per spectrum was normalized to one.

As derived glycan traits represent glycosylation changes shared by a group of structurally related N-glycans and enable better interpretation of the biological effects of glycosylation [21, 26], we calculated derived glycan traits from these individual glycans (directly detected) using specific formulas based on the shared similar structures of individual glycans in RStudio (version 4.0.3) (Supplementary Table S2). The subject of the calculation is indicated by the last letter and the group on which it is calculated by the preceding letters. For instance, translates A4SF into ‘the fucosylation (F) within sialylated (S) tetra-antennary (A4) glycans’ (Supplementary Table S2).

### 2.6 Statistical analysis

The baseline clinical characteristics of the cohort were summarized using standard descriptive statistics. Continuous variables were presented as mean (standard deviation [SD]) or median (interquartile range [IQR]), as appropriate. Whereas categorical data were presented as frequency and percentage (Table 1). Multivariate and univariate statistical analyses were used to identify discriminative derived glycan traits driving the classification of EC and HC: Orthogonal partial least squares - discriminant analysis (OPLS-DA) and sparse partial least squares - discriminant analysis (sPLS-DA) were performed to show whether the two groups of EC and HC exist significant differences regarding the whole serum N-glycome through MetaboAnalyst 5.0. Meanwhile, the derived glycan traits that made significant contributions to the difference between EC and HC were identified by the S-plot of OPLS-DA. The most discriminative derived glycan traits ranked by variable importance in projection score (VIP) based on S-plot were visualized as the heatmap. Logistic regression was used to identify derived glycan traits significantly associated with the occurrence of EC in RStudio (age was included as a covariate in the models for the disease-related tests). Together with the results of the Mann-Whitney U test (non-normally distributed data) and OPLS-DA, the results of logistic regression were also used as the strict filter for screening the candidate biomarkers. After applying strict criteria (i.e. VIP>1, logistic regression p<0.000431 [0.05/116], and U test p<0.000431; 116 represents the number of derived glycan traits compared in this study) to filter the derived glycan traits, a set of derived glycan traits would be screened out and tentatively identified as candidate biomarkers for EC diagnosis.

**Table 1.**
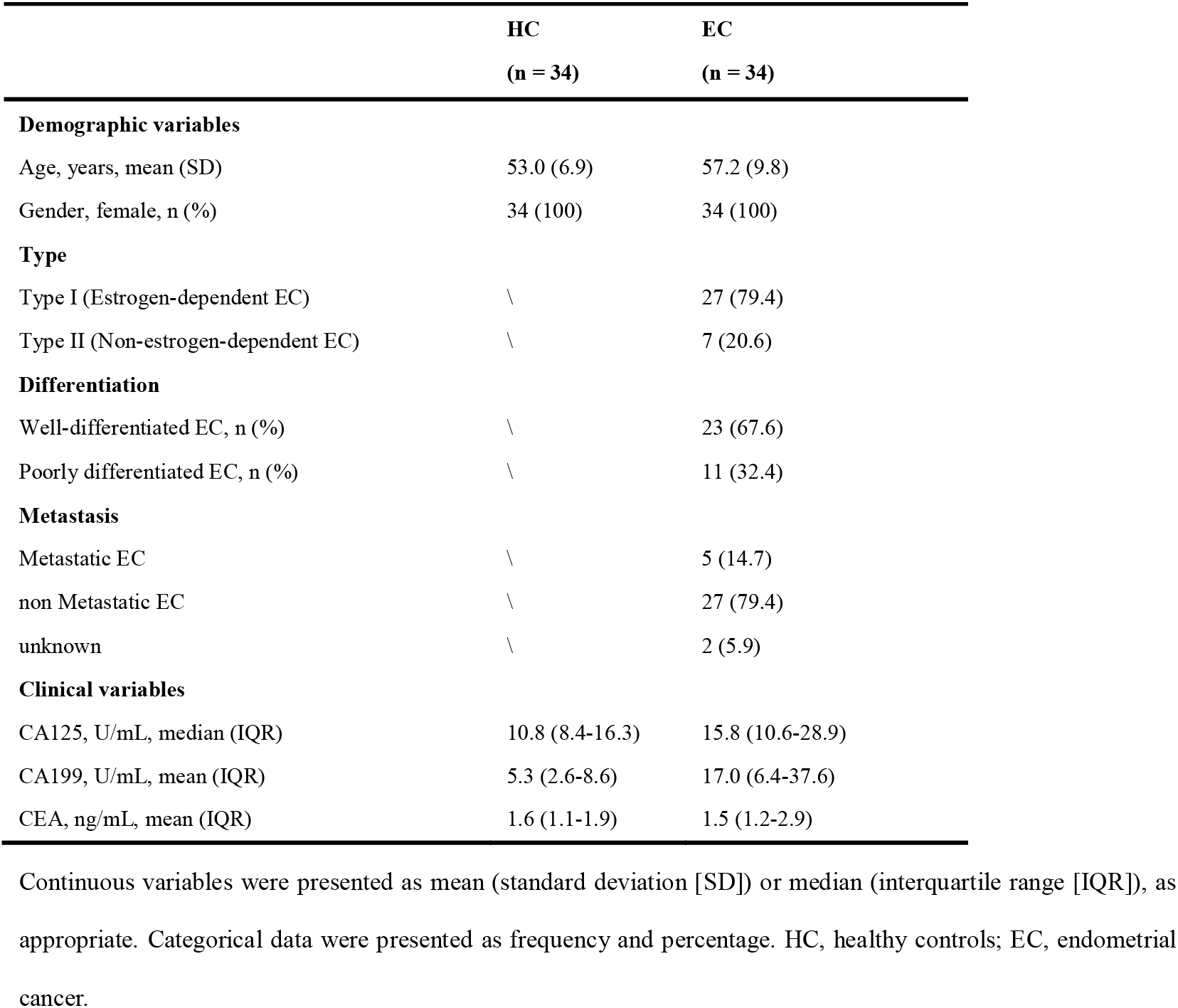
Clinicopathological characteristics of the cohort in the present study.

Thereafter, receiver operating characteristic (ROC) analyses were used to evaluate the classification accuracy of these candidate biomarkers and built the optimal glycan panel: Classical univariate ROC analyses can give the area under the curve (AUC) values and assess/rank the performance of each derived glycan trait; Multivariate ROC exploratory analyses were used to build the glycan panels based on derived glycan traits, test their performance in the classification, and screen out the most important derived glycan traits that may be used to build the optimal glycan panel. When doing the multivariate ROC exploratory analyses, we used three common algorithms/models (random forest [RF], support vector machine [SVM], and partial least squares [PLS]) as the classification method to build the glycan panels and to select the important glycan traits. Multivariate ROC curves were generated by Monte-Carlo cross-validation (MCCV) using balanced sub-sampling (in each MCCV, 2/3 of the samples are used to build classification models, and the left 1/3 of the samples are used for the validation). 100 cross-validations were repeated to calculate the AUC and 95% confidence interval (CI) as well as the average accuracy of each model. ‘AUCs ≥ 0.9’ represents highly accurate tests, ‘0.8 ≤ AUCs < 0.9’ accurate tests, ‘0.7 ≤ AUCs < 0.8’ moderately accurate tests, and ‘AUCs < 0.7’ uninformative tests [17, 27]. It is important to point out that, when exploring the biomarkers for EC differentiation, as only two derived glycan traits were found associated with EC differentiation, we did not perform the same analyses such as OPLS-DA and heatmap as when exploring diagnostic biomarkers.

## 3. Results

The serum N-glycomes of the EC cohort (Table 1) were profiled by MALDI-TOF-MS. At the global serum level, we identified 109 N-glycans, 79 of which passed the quality criteria for subsequent data analysis. It includes all typical types of N-glycans, including high-mannose-, hybrid-, and complex-type of N-glycans (refer to Supplementary Table S1 for the complete list of N-glycans analyzed in this study). A representative annotated spectrum of MALDI-TOF-MS for N-glycome of EC is displayed in Figure 1A. The 79 directly detected glycans were grouped into 116 derived glycan traits based on shared similar structures including the number of antennas (A), galactosylation (G), bisection (B), fucosylation (F), sialylation (S), and linkage-specific sialylation (E or L) using specific formulas (Supplementary Table S2, Figure 1B). As reported previously, derived glycan traits reflect biosynthetic pathways of glycans and enable better interpretation of the biological effects of glycosylation [21, 26], we mainly interpreted and presented the results of derived glycan traits in the present study.

**Figure 1.**
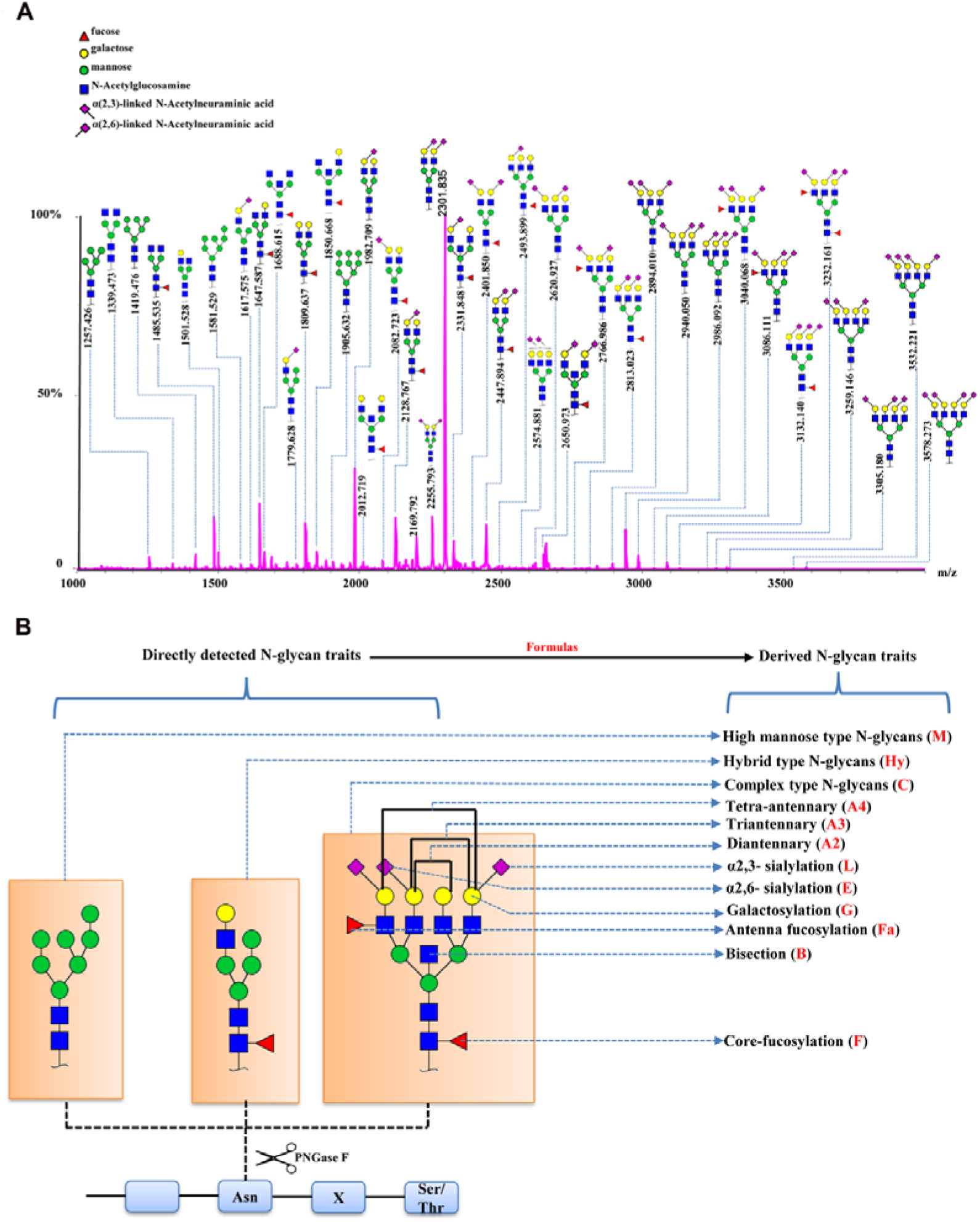
Serum N-glycome features of EC. **(A)** A representative annotated spectrum of MALDI-TOF-MS for N-glycome of EC presenting the major N-glycans. **(B)** Graphical representation of N-glycan types and derived N-glycan traits based on the shared similar structures of individual N-glycans.

### 3.1 Data reliability

As shown in Supplementary Table S3, the average RSD of the 116 derived glycan traits in the eight technical replicates was 4.06%, demonstrating good method repeatability.

### 3.2 Differences in serum N-glycan profiles between HC and EC

In order to evaluate the differential expressions of glycan profiles between HC and EC, OPLS-DA and sPLS-DA were conducted based on derived glycan traits. The scores plot of the OPLS-DA analysis showed that the HC and EC groups were well separated through N-glycan profiling (Figure 2A). The OPLS-DA of glycomic data showed cumulative values of R2 (Y) = 88.1% and Q2 = 82.8% and the permutation (cross-validation) test is shown in Supplementary Figure S1 (the intercept of the Q2 regression line and the vertical axis is less than zero), indicating that the model has a high separating capacity and is not overfitting. The sPLS-DA analysis was performed to validate the discriminating capacity of N-glycans. As displayed in Supplementary Figure S2, the sPLS-DA plot also showed that the two groups were well separated. Therefore, multivariate analyses demonstrated that distinct differences in N-glycan profiles existed between HC and EC.

**Figure 2.**
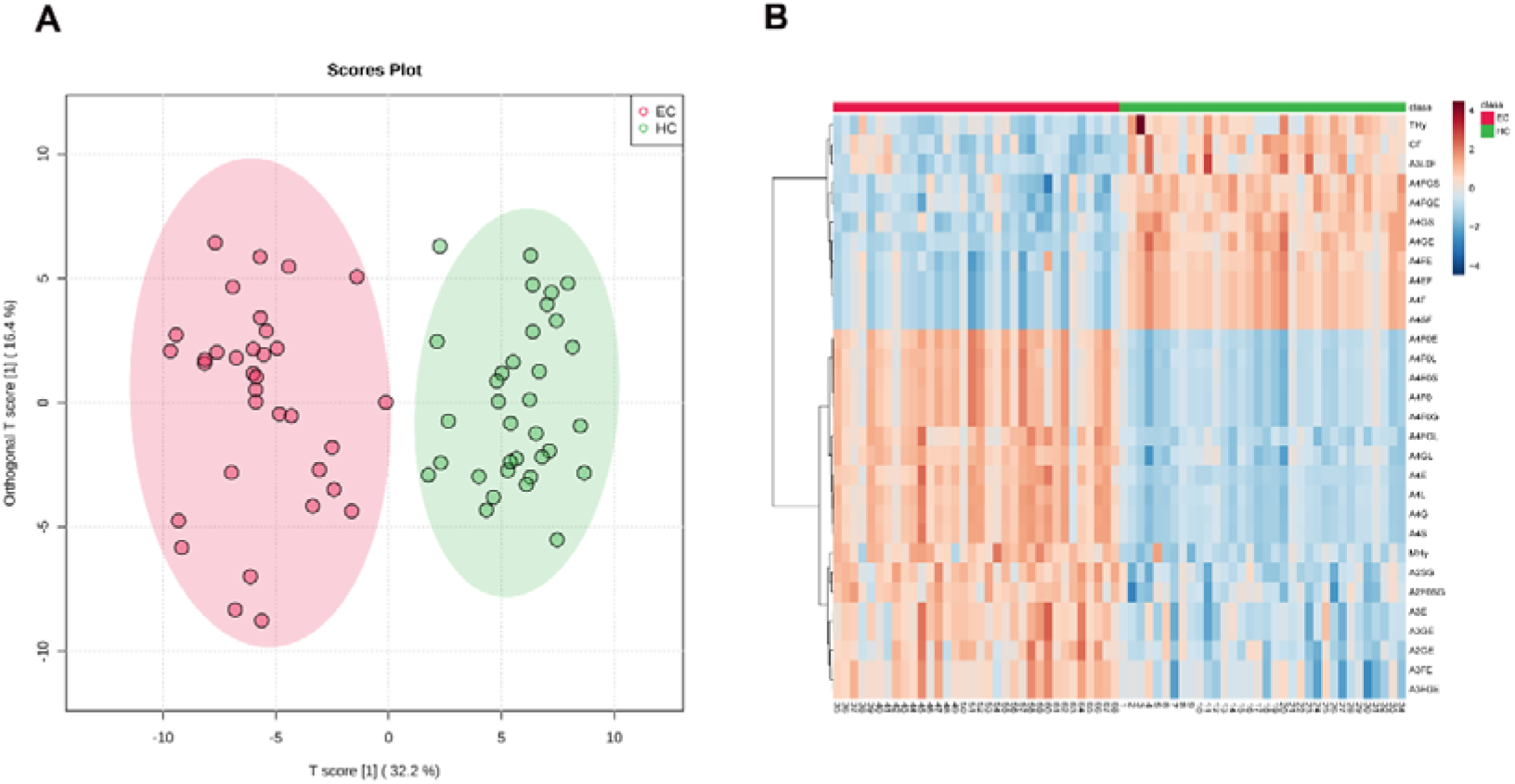
**(A)** Scores plot of the OPLS-DA between EC (red circle) and HC (green circle) groups. The scores plot is based on derived glycan traits. Areas of 95% confidence interval are highlighted in red and green for EC and HC, respectively. OPLS-DA, orthogonal partial least squares - discriminant analysis; EC, endometrial cancer; HC, healthy controls. **(B)** Heatmap of the top 30 discriminating derived glycan traits between EC and HC. The scale displays the log-transformed values of the relative intensities of the derived glycan traits. OPLS-DA, orthogonal partial least squares - discriminant analysis; EC, endometrial cancer; HC, healthy controls.

The derived glycan traits that made significant contributions to the difference between EC and HC were identified by the S-plot of OPLS-DA analysis (Supplementary Figure S3). The features (derived glycan traits) with VIP greater than 1 were screened out and the top 30 features (derived glycan traits) were plotted as the heatmap in Figure 2B, showing significant alterations of serum N-glycans in EC compared with that in HC (Figure 2B). Of note, the results of the multivariate analyses were consistent with that of the U test (Table 2).

**Table 2.**
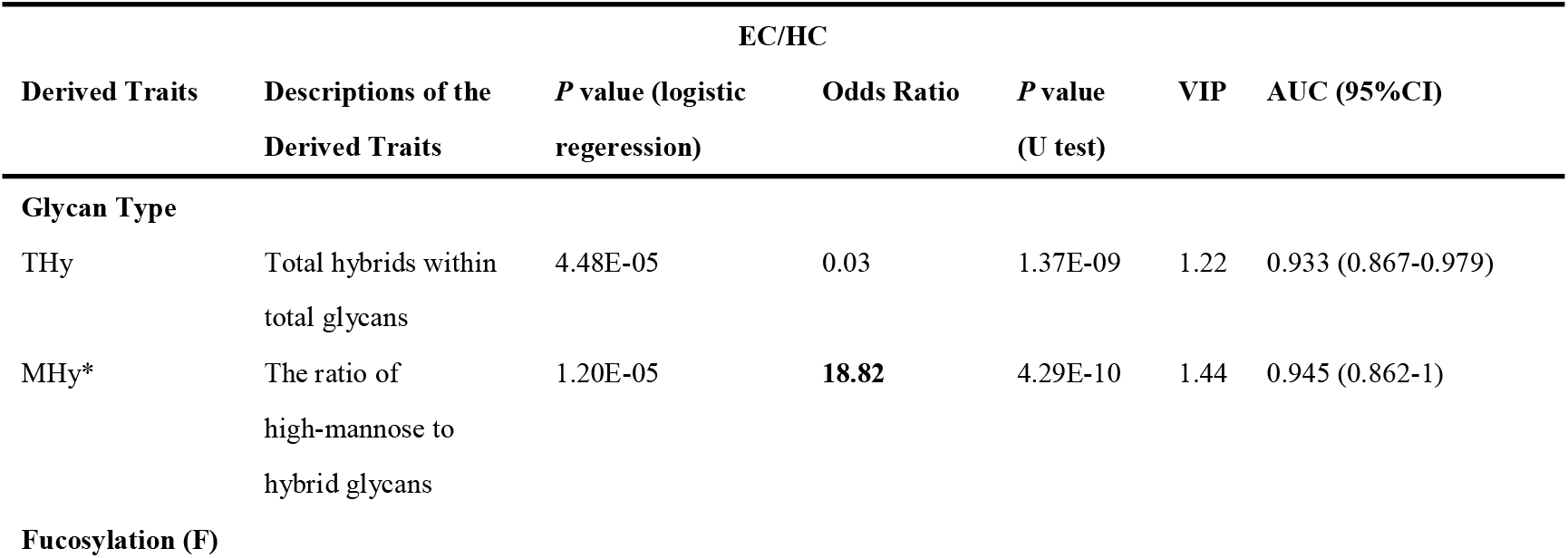
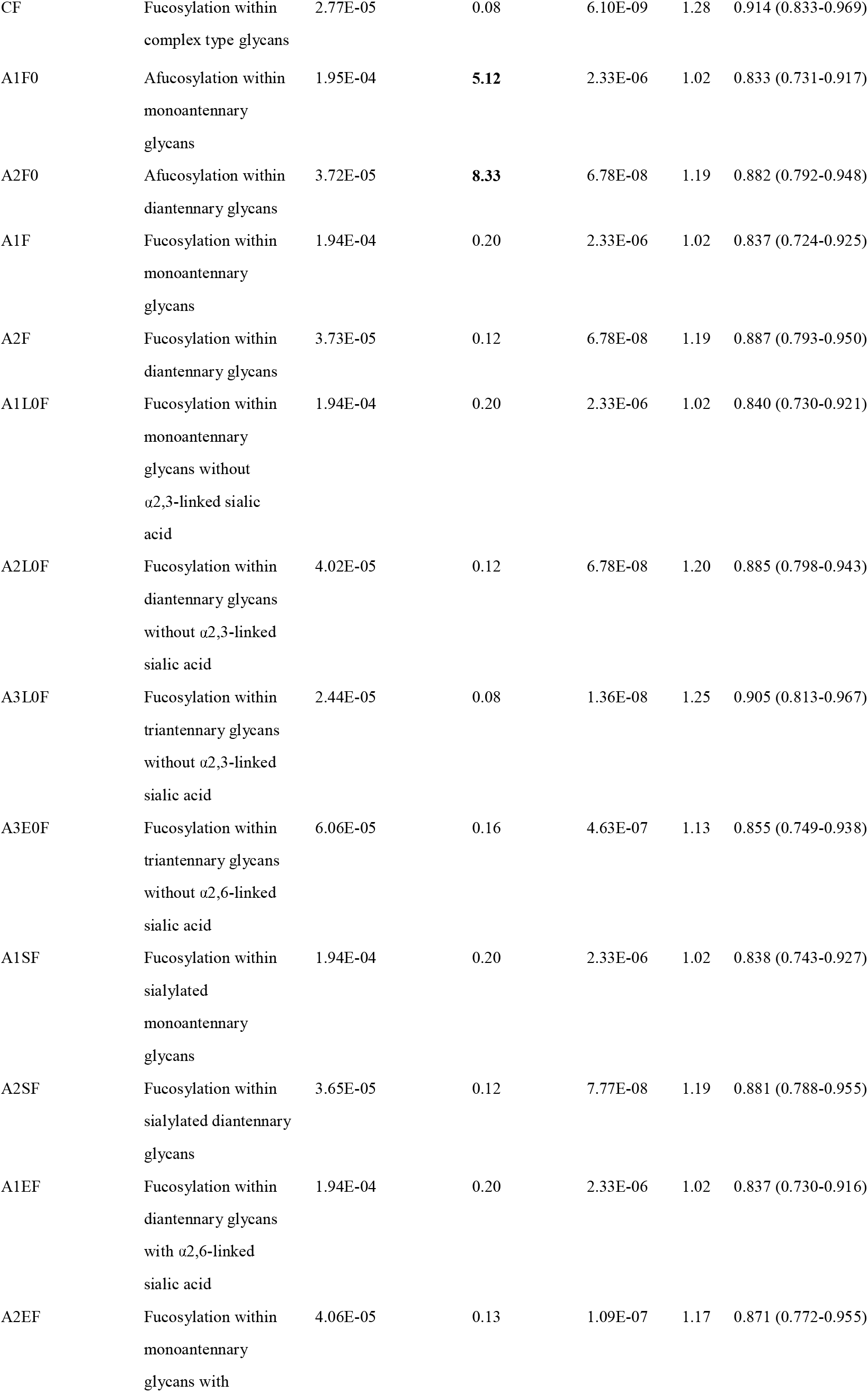
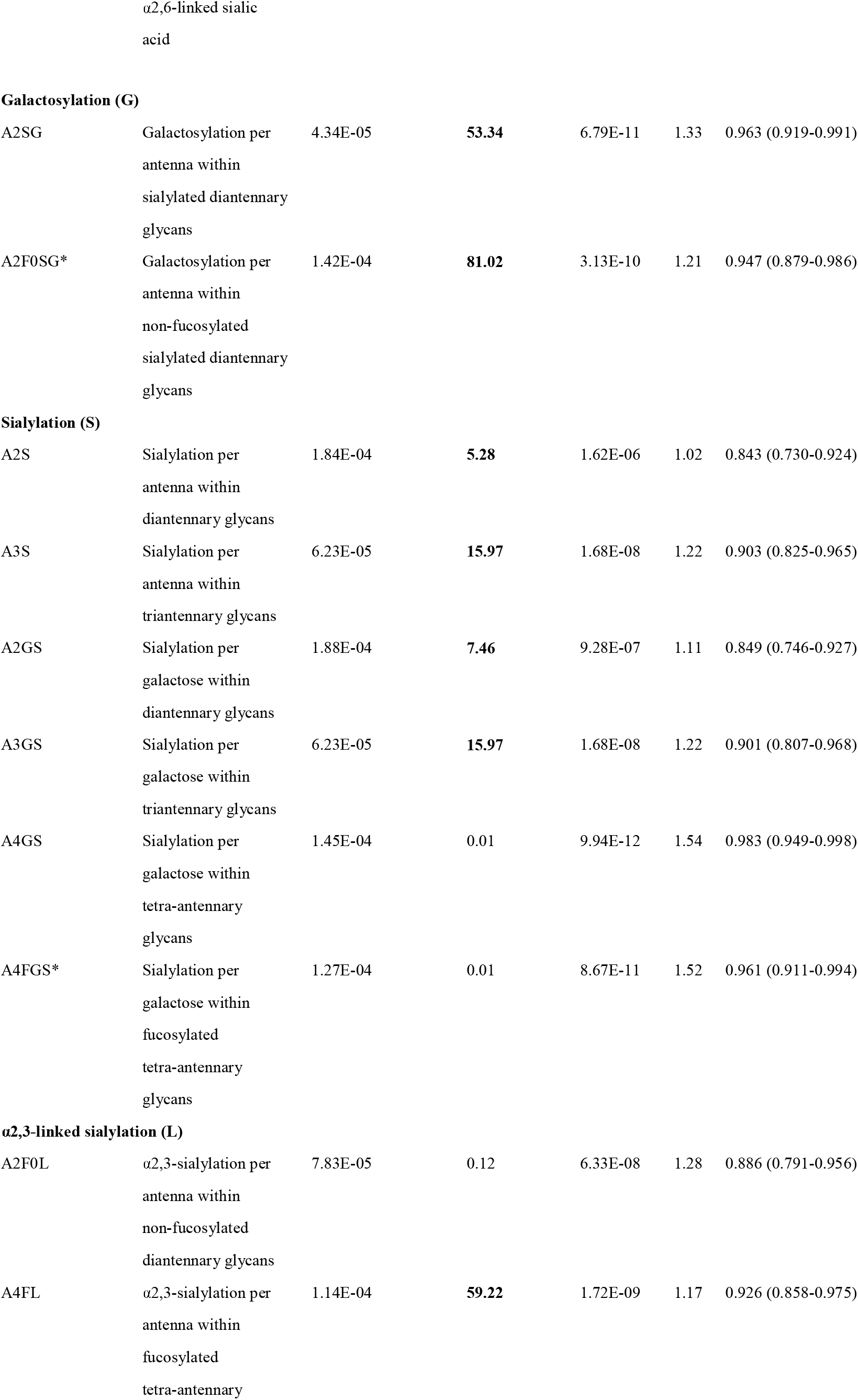
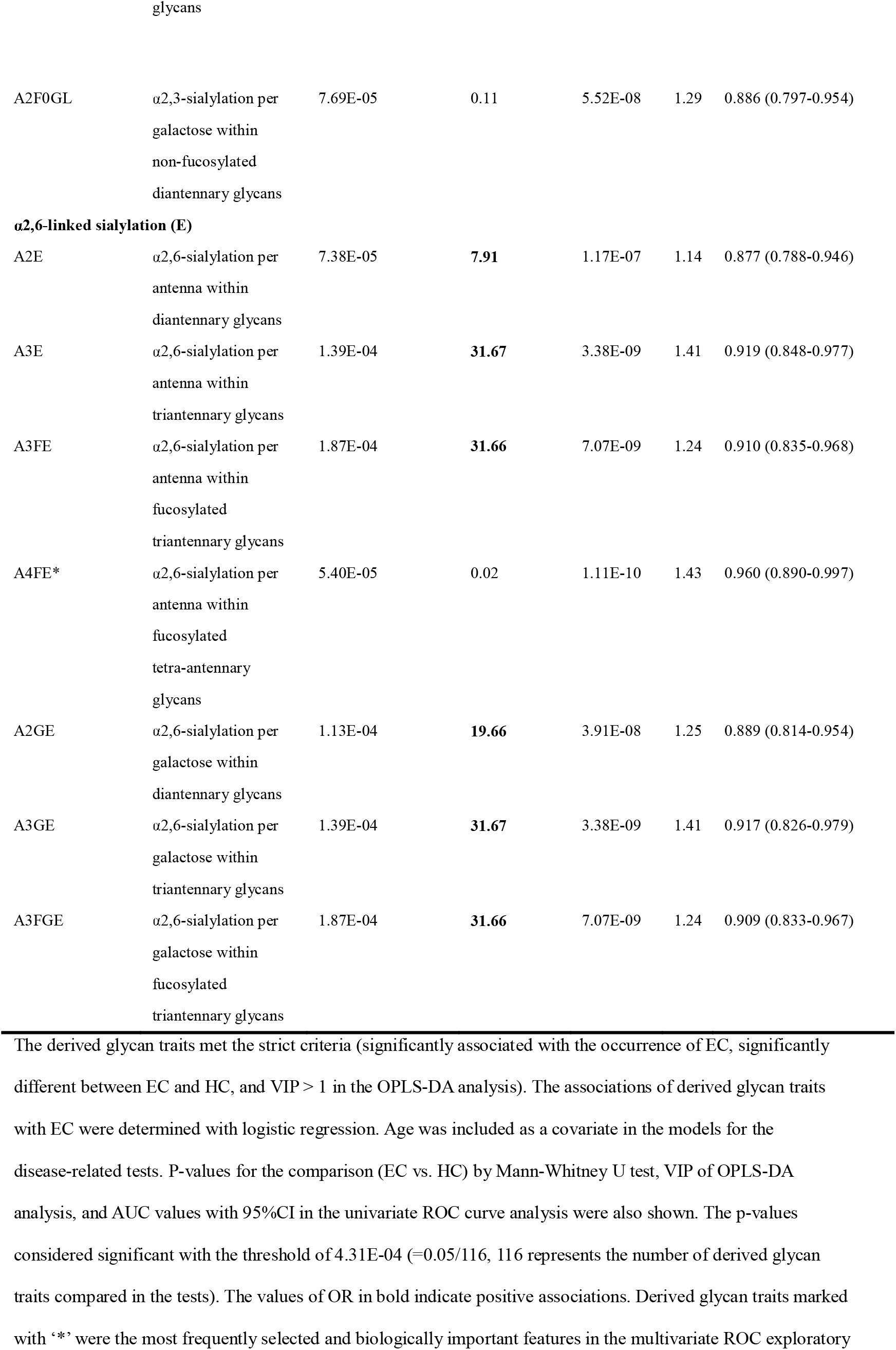
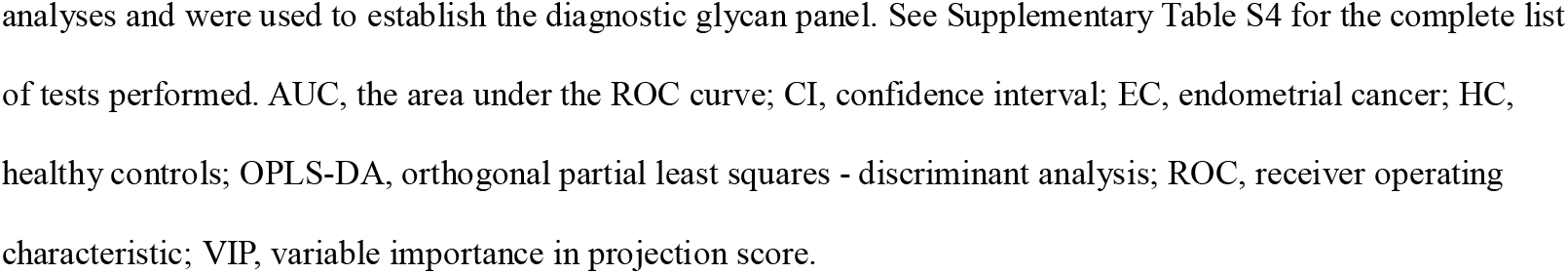
The list of a set of 33 derived N-glycan traits identified as candidate diagnostic biomarkers.

### 3.3 Identification of N-glycan traits associated with EC

By using logistic regression, derived glycan traits associated with the occurrence of EC were determined (Table 2, Supplementary Table S4). As shown in Table 2 and Supplementary Table S4, there were a series of derived glycan traits significantly associated with EC, mainly including the hybrid-type N-glycans and fucosylation-, galactosylation-, and sialylation-related glycans. From Table 2 we can see that sialylation within diantennary and tri-antennary glycans (A2S, A3S, A2GS, A3GS) was higher in EC than in controls and positively associated with EC, which was mainly due to the increase of α2,6-linked glycan species (A2E, A3E, A3FE, A2GE, A3GE, A3FGE). In contrast, sialyation within tetra-antennary glycans (A4GS, A4FGS) was lower in subjects with EC than in controls and negatively associated with EC, which was mainly driven by the decrease of α2,6-linked sialylation (A4FE). α2,3-linked sialylation showed opposite changes in EC compared with α2,6-linked glycan. In addition, fucosylation (CF, A1F, A1SF, A1EF, A1L0F, A2F, A2SF, A2EF, A2L0F, A3L0F, A3E0F) and hybrid-type N-glycans (THy) were decreased in the serum of EC patients compared to that of controls and negatively associated with EC (Table 2, Supplementary Table S4). Altered galactosylation was also found in EC compared to controls. Galactosylation within sialylated diantennary glycans (A2SG) showed a significant increase in EC patients and was positively associated with EC. This was mainly caused by the increase of galactosylation of afucosylated glycans (A2F0SG) (Table 2, Supplementary Table S4).

### 3.4 Glycan biomarker discovery and validation for EC diagnosis

After comprehensively characterizing the serum N-glycome of EC and identifying the derived glycan traits associated with the occurrence of EC, we attempted to screen out candidate biomarkers capable of classifying EC from HC. After applying strict criteria (VIP>1, logistic regression p<0.000431, and U test p<0.000431) to filter the most discriminative derived glycan traits, a set of 33 derived glycan traits were screened out and tentatively identified as candidate biomarkers (Table 2). The individual discriminative power of each derived glycan trait was also supported by classical univariate ROC analysis (the AUCs of all these traits were greater than 0.8) (Table 2). A4GS, A2SG, A4FGS, A4FE, A2F0SG, and MHy were the top-ranked (based on AUC) derived glycan traits contributing to discrimination between EC and HC, showing high capability as diagnostic biomarkers for EC (Table 2).

In order to optimize the candidate diagnostic biomarkers and build an optimal N-glycan panel, we performed multivariate ROC exploratory analyses based on the multivariate model/algorithm of RF. As presented in Figure 3A, ROC curves built with the variable number of features (the 33 candidate glycan traits mentioned above) show superior performance in distinguishing EC from HC with AUCs ranging from 0.970 to 0.996. The AUC values increased gradually with the increase of the number of derived glycan traits in the panels and reached the highest values with 10 derived glycan traits (Figure 3A). Then, adding more derived glycan traits no longer contributes to the AUC value (Figure 3A). At the same time, multivariate ROC exploratory analyses screened out the most discriminative derived glycan traits in the panels (Figure 3B, Supplementary Figure S4). Of note, the exploratory analysis didn’t point out which specific derived glycan traits were used to construct each glycan panel, but it indicated the most frequently selected derived glycan traits when building the panels, which facilitate the building of the optimal glycan panel. Importantly, the six most frequently selected derived glycan traits (A4FGS, A4GS, MHy, A4FE, A2F0SG, and A2SG) (Figure 3B, Supplementary Figure S4) in the multivariate ROC analyses were the top-ranked derived glycan traits in classical univariate ROC analysis (Table 2). Therefore, the optimal panel may be obtained by combining the six most discriminative derived glycan traits. However, considering the collinearity of derived glycan traits (A4FGS is a subset of A4GS and A2F0SG is a subset of A2SG, which causes the same effect to be included twice in the panel), we should include only one in each combination (A4FGS-A4GS, A2F0SG-A2SG) in the panel. As A4FGS and A2F0GS contain more biological information about the fucosylation state of the glycans, finally, we select the combination of A4FGS, A2F0SG, MHy, and A4FE as a new panel in an attempt to create the optimal panel for EC diagnosis. The AUC value of this panel in identifying EC is 0.993 (95%CI, 0.955-1) (Figure 3C), slightly lower than the values of panels consisting of 10, 20, and 33 features (Figure 3A). These results suggested that the glycan panel built with the four derived glycan traits is the optimal diagnostic glycan panel, which not only includes the least number of features and achieves very good performance but also is biologically reliable. In addition, the predictive accuracies of the panels with different numbers of features (derived glycan traits) were shown (Figure 3D, Supplementary Table S5). To evaluate the robustness of the panel built with the four glycan traits, we used another two classification methods (SVM and PLS models/algorithms) to further validate the results. The AUC value with 95%CI and average accuracy based on 100 cross-validations of each model were calculated (Supplementary Figure S5, Table S5). The AUCs for SVM and PLS models/algorithms were 0.987 (95%CI, 0.946-1) and 0.989 (95%CI, 0.950-1), respectively (Supplementary Figure S5, Table S5). In addition, the average predictive accuracies of this optimized panel were greater than 94% applying any of these three models/algorithms (Supplementary Figure S5, Table S5). These results confirm that the panel built based on the four most discriminative and biologically reliable derived glycan traits is promising as a biomarker for EC diagnosis. In contrast, the AUC values of classical gynecologic tumor markers (CA125, CA19-9, carcinoembryonic antigen [CEA], and the combination of the three markers) in detecting EC were only 0.716 (95%CI, 0.578-0.841), 0.520 (95%CI, 0.371-0.660), 0.688 (95%CI, 0.548-0.801), and 0.797 (95%CI, 0.627-0.925), respectively, lower than the AUC obtained with the glycan panel (Figure 3, Supplementary Figure S6).

**Figure 3.**
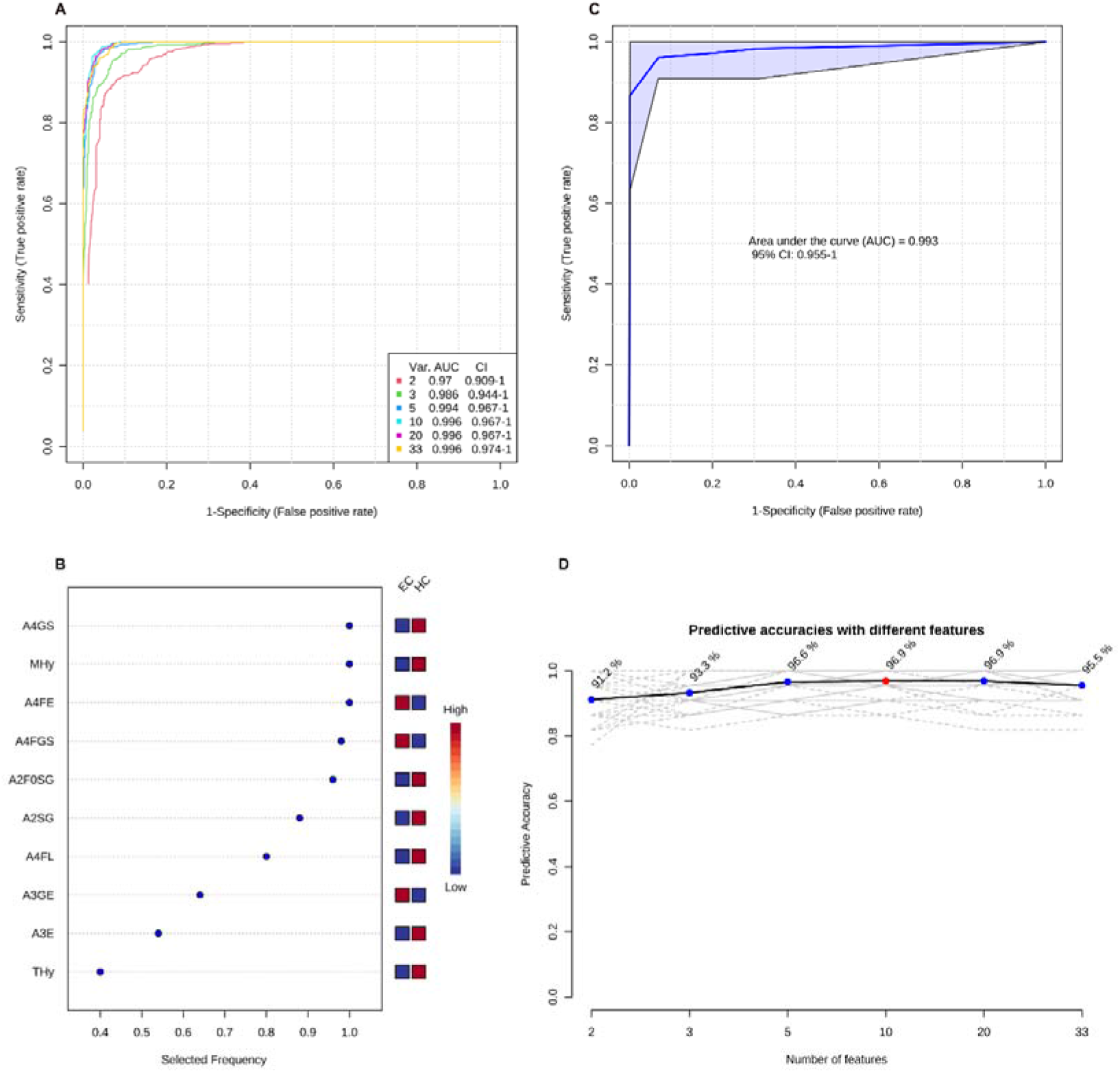
The establishment of the optimal diagnostic glycan panel based on the random forest model/algorithm. **(A)** Classification performance of the glycan panels built with the variable number of derived glycan traits. **(B)** The most frequently selected derived glycan traits when building the glycan panel constructed by 10 features (derived glycan traits). Please refer to Supplementary Figure S4 for the most frequently selected derived glycan traits when building all the six glycan panels (panel 1-panel 6) shown in Figure 3A. **(C)** the performance of the optimal diagnostic glycan panel built with the four most important (discriminative and biologically reliable) derived glycan traits. **(D)** The predicted accuracies of the panels built with the variable number of derived glycan traits.

### 3.5 Identification of N-glycan traits associated with phenotypes of EC differentiation

The associations of the derived N-glycan traits with well or poorly differentiated types of EC were tested by logistic regression on standardized data with age as a covariate in the model. In EC patients, poor differentiation was found negatively associated with hybrid-type N-glycans (derived glycan trait, THy; p = 0.018; OR = 0.11) (Supplementary Table S6), whereas positively associated with complex-type N-glycans (derived glycan trait, TC; p = 0.035; OR = 4.86) (Supplementary Table S6). Consistently, as shown in the boxplots, the levels of THy and TC decreased and increased from HC to well-differentiated to poorly-differentiated groups, but the increasing trend of TC was not statistically significant for ‘HC vs. well-differentiated EC’ (the mean values of TC in HC and well-differentiated EC were 0.9652 and 0.9654, respectively) (Figure 4A-B). Considering the total of THy, TC, and TM (total amount of high mannose type glycans) should be 100% (as relative amounts are used), we further assessed TM in the subgroups of the cohort. We found that in poorly differentiated EC, THy decreases at the expense of TC. While, in well-differentiated EC, THy decreases not at the expense of TC but TM (Figure 4C). We also tried to explore the associations of classical gynecologic tumor markers (CA125, CA19-9, and CEA) with the differentiation types of EC. However, no statistically significant associations were found between them (Supplementary Table S6).

**Figure 4.**
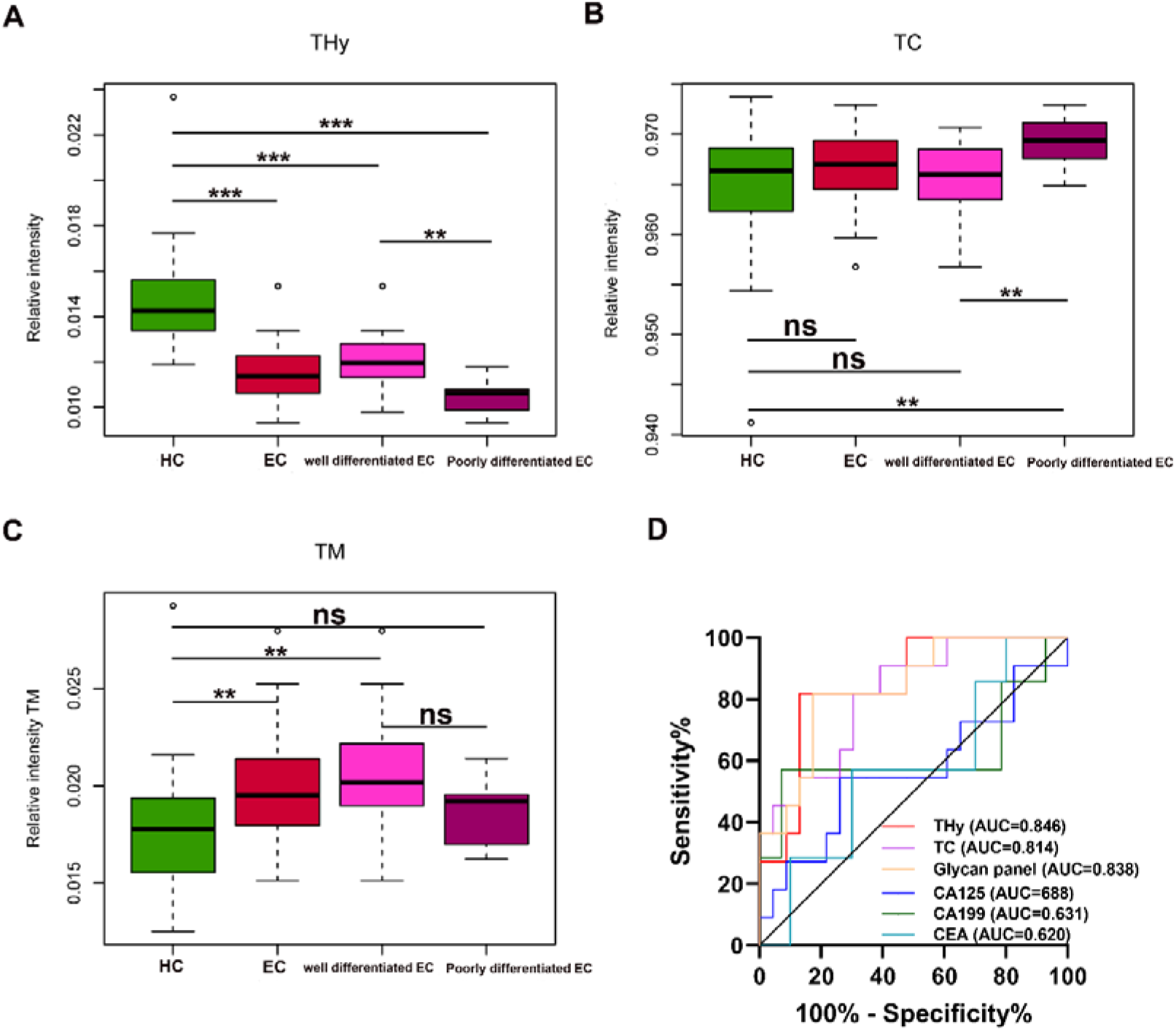
**(A-C)** Boxplots for THy, TC, and TM in HC and well and poorly differentiated EC. ***□=□p-value <0.001, **□=□p-value <0.01, * p-value <0.05, ns□=□not significant (U test, without multiple testing correction). THy, total amount of hybrid-type glycans; TC, total amount of complex-type glycans; TM, total amount of high mannose type glycans; EC, endometrial cancer; HC, healthy controls. **(D)** ROC curves of glycans and the classical markers in discriminating well and poorly differentiated EC. ROC: receiver operating characteristic; EC, endometrial cancer.

### 3.6 Glycan biomarker for EC stratification/ prognosis

As derived N-glycan traits THy and TC were strongly associated with the differentiation types of EC, ROC curve analyses were further conducted for THy, TC, and glycan panel built based on the combination of the two derived traits to determine their predictive values for stratifying EC to well or poorly differentiated groups. As displayed in Figure 4D, the AUCs of THy, TC, and the glycan panel were 0.846 (95%CI, 0.710-0.982), 0.814 (95%CI, 0.666-0.963), and 0.838 (95%CI, 0.697-0.979), respectively, suggesting that the glycans have a high predictive value for EC phenotyping (‘AUCs≥0.8’ represents accurate predictive tests). In addition, we also evaluated the performance of classical gynecologic tumor markers. Nevertheless, the results showed that these markers had no value in the stratification of EC (AUCs<0.7) (Figure 4D). As poorly differentiated EC patients are always associated with poor prognosis and a high risk of metastasis and relapse, the predictive glycan traits/panel we discovered in this section may be useful for patient stratification as well as the evaluation of patients’ prognoses.

## 4. Discussion

Timely and accurate diagnosis and patient stratification are crucial for the prognosis and management of EC patients. Due to the limitations of the existing methods (e.g., the routinely used gynecologic tumor markers have limited value for EC, and currently, the differentiation type of EC can only be known after surgery), exploring new and effective methods is vitally important and urgent.

The advent of high-throughput and highly sensitive technologies has greatly advanced our ability to comprehensively understand the complexity and heterogeneity of glycosylation in humans, which allowed pathological mechanisms to be elucidated at the level of glycome [28-31]. Besides, it also provides new approaches for discovering biomarkers [32, 33].

Glycosylation in EC has attracted much attention in recent years. For instance, Nishijima *et al*. compared the glycosylation of three cell lines (well-differentiated EC, poorly differentiated EC, and normal endometrium) using lectin microarray analysis and found apparent differences among these cell lines in glycans [34]. A recent study by Mittal *et al*. investigating tissue N-glycan changes in EC patients with LNM or without compared to normal endometrium confirmed the dysregulation of glycosylation in EC using MALDI MS imaging [15]. Besides, they found a core-fucosylated diantennary N-glycan (H5N4F1), which may originate from IgG to a large extent, was associated with the LNM of EC [15]. Consistently, another study by Lin *et al*. investigated the IgG N-glycome of EC patients. The results of this study revealed aberrant N-glycans on serum IgG in EC and their potential role in EC diagnosis, and that includes H5N4F1 (GP14) [14]. These pilot studies indicated alterations of glycosylation in EC and the potential of glycan-based biomarkers for EC. The previous studies, however, used low-sensitive approaches (e.g., lectin microarray) [34] or lost the linkage information of sialylation (without sialylation derivatization or protection) [14, 15], which allowed the detection of a limited number of N-glycans. Moreover, they focused on glycosylation in cells, tissues, or serum IgG. Nevertheless, to date little is known about the glycosylation patterns in serum at the global level, which has shown potential as biomarkers in various diseases and is often disease-specific. In the present study, we employed a high-throughput MALDI-TOF-MS platform to detect the serum N-glycome of EC patients (consisting of different types) and matched HC. The state-of-the-art MS-based N-glycomic method applied here was proven to be robust with good repeatability and the further linkage-specific sialic acids derivatization increased the effective depth of N-glycosylation analysis (more extensive coverage of glycan structures), which added more important glycobiological information. Thereby, for the first time, we provide the full features of serum N-glycome of EC. Importantly, we observed associations of N-glycans with EC or its phenotypes of differentiation, which may represent the specific N-glycomic fingerprint of EC. Furthermore, we screened and optimized glycan traits/panels that showed high promise as non-invasive biomarkers for EC diagnosis and stratification/prognosis.

Glycosylation is influenced by pathophysiological status. Changes in serum glycans in the present study can be partly attributed to the biosynthetic or metabolic dysregulation that occurs during the initiation and progression of EC. The six most discriminative derived glycan traits involve sialyation, galactosylation, and high-mannose- and hybrid-type glycans (i.e., A4GS, A4FGS, A4FE, A2SG, A2F0SG, and MHy). Sialyation within tetraantennary glycans (A4GS, A4FGS) was lower in subjects with EC than in controls, which was mainly driven by the decrease of α2,6-linked sialylation (A4FE). In contrast, sialyation within diantennary and tri-antennary glycans was higher in EC than in controls, which was mainly due to the increase of α2,6-linked glycan species. Therefore, the alterations of sialylation in EC were mainly caused by the difference in α2,6-linked sialylation species. The effect seen might largely be influenced by IgG, as IgG is the most abundant glycoprotein in serum and carries mainly diantennary glycans on which the terminal sialic acids are almost exclusively α2,6-linked species. There is growing evidence that demonstrated linkage-specific (α-2,3- or α-2,6-linkage) sialylated N-glycans are involved in the occurrence and progression of cancers including EC [35, 36]. It has been proposed that α2,6-linked sialic acids promote cancer cell survival by inhibiting galectin binding [37, 38]. Moreover, previous studies indicated that α2,6-sialylation may be linked to systemic inflammation [39], reflecting the possible inflammation changes in EC. Consistently, cancer-associated chronic inflammation has been established in gynecological malignancy, especially in EC [40]. One may speculate that the α2,6-sialylation changes in EC may be implicated in the chronic inflammation-related pathogenic mechanism of EC. Future work should determine the direct functions and mechanisms behind the changes in α2, 6-linked sialic acid in the serum. In addition to sialylation, one of the six most discriminative derived glycan traits - A2SG (galactosylation within sialylated diantennary glycans) showed a significant increase in EC patients. This was mainly caused by the increase of galactosylation of afucosylated glycans (A2F0SG). We can find that the variation tendency of galactosylation within diantennary glycans (A2SG and A2F0SG) was consistent with that of sialylation with diantennary glycans (A2S, A2GS, AE, and A2GE) (both increased in EC), as glycans with galactose are the substrate of sialylation. A2SG and A2F0SG are mainly derived from glycoproteins secreted by the liver (e.g., fibrinogen, hemopexin, haptoglobin, a1B-glycoprotein, and IgA) [26]. Importantly, some of these glycoproteins (e.g., fibrinogen and haptoglobin) have been proposed as markers for EC [41-43]. Therefore, the observed increase of galactosylation within diantennary glycans might reflect alterations in hepatic synthesis and/or aberrant glycosylation of liver-secreted glycoproteins in EC, which were also supported by previous studies [41-43]. At the same time, MHy (the ratio of high-mannose glycans to hybrid-type glycans, TM/THy) was found positively associated with EC in this study. Denominator THy was found negatively associated with EC and numerator TM (not statistically significant after the multiple testing correction) was found positively associated with EC (Figure 4, Supplementary Table S4). A higher abundance of high-mannose glycans in tumor regions of EC patients compared to adjacent normal regions was also reported recently [15], consistent with our findings here. The roles of high-mannose and hybrid-type glycans in EC have not been addressed in current studies.

Our results indicate poor differentiation of EC is accompanied by decreased total amount of hybrid-type glycans (THy) and increased total amount of complex-type glycans (TC). While, well differentiation of EC is accompanied by decreased THy and increased total amount of high mannose type glycans (TM). In line, evidence for distinct glycosylation between well or poorly differentiated EC cell lines has been found previously [34]. Though the role of hybrid-type glycans in EC has not yet been established, we assume that abnormal THy (along with the divergent trend in TC and TM) may be implicated in the mechanisms of cell differentiation of EC, and thus warrants further investigation to better interpret the glycomic data. It is worth mentioning that the TC group is large and encompasses more than 96% of all glycans. Though statistically this derived glycan trait was screened out as a discriminator for patient stratification, biologically this trait is non-informative. In addition, as THy and TC are strongly correlated (relative quantitative glycomics are used in this study), the glycan panel based on the two derived glycan traits includes the same effect twice, resulting in that there was no additive effect of the combination of both traits. The derived glycan trait THy associated with the phenotypes of differentiation of EC showed high promise to serve as a biomarker for EC stratification/prognosis and may play a critical role in predicting the types and guiding the treatment of EC. Whereas, this requires further long□term follow□up and mechanism studies to better validate their clinical utility.

The present study has several limitations. Firstly, promising glycan biomarkers for EC were identified, however, we did not directly verify the underlying pathophysiological mechanisms. The MS-based glycomic approach does not provide the protein origin of these glycans. Supportive evidence in this regard by quantifying protein□specific glycans and combining levels of the corresponding glycoproteins will give in□depth insights into the mechanisms, but it’s still technically challenging. Besides, glycopeptide analysis is also an additional technique, which should be considered in further studies. Secondly, the assignment of the glycan species does not exclude isomers (except for the linkage of sialylation). Thirdly, the Cancer Genome Atlas (TCGA) defined a new molecular classification system, which categorizes EC into four subgroups based on the molecular profile [44]. The incorporation of the molecular classification method can provide better prognostic predictions in EC [45]. In the present study, we investigated associations of glycan traits with the differentiation types of EC to explore promising glycan biomarkers for patient stratification. Considering the advantages of the new molecular classification system, associating the glycans traits with the molecular profiles may discover better glycan biomarkers for the prediction of patients’ prognoses. Molecular grouping is not widely used yet, since it adds a significant cost to histological interpretation [2]. Future studies can consider evaluating the associations of glycan traits with the new molecular grouping. Fourthly, the current sample size remains relatively small and further validations in larger multi-center cohorts are needed based on a prospective study design. Despite the limitations, our findings to the best of our knowledge provide the initial evidence supporting the utility of serum N-glycome signature as potential diagnosis and phenotyping markers for EC.

## 5. Conclusion

In conclusion, we for the first time provided the full signature of serum N-glycome of EC using MS-based glycomic approaches. Our results revealed distinct differences in serum glycan patterns between EC and HC and robust associations of N-glycans with EC or its phenotypes of differentiation, suggesting that EC has a specific glycomic fingerprint. The glycan panels built with the most discriminative glycan features hold high promise as non-invasive biomarkers for EC diagnosis and stratification/prognosis. Despite its limitations, this work provides new insights into the diagnosis, stratification/prognosis, and pathogenesis of EC.

## Data Availability

The data supporting the conclusion of the present study are included in the article. Additional data can be obtained from the corresponding author upon request.

## List of abbreviations

ACN: acetonitrile
AUC: area under the curve
CA125: carbohydrate antigen 125
CA15-3: carbohydrate antigen 15-3
CA19-9: carbohydrate antigen 19-9
CEA: carcinoembryonic antigen
CI: confidence interval
EC: endometrial cancer
HC: healthy controls
HILIC-SPE: hydrophilic interaction liquid chromatography solid-phase extraction
IQR: interquartile range
LNM: lymph node metastasis
MALDI-TOF-MS: matrix-assisted laser desorption/ionization time-of-flight mass spectrometry
MCCV: Monte-Carlo cross-validation
OPLS-DA: orthogonal partial least squares - discriminant analysis
PLS: partial least squares
RF: random forest
ROC: receiver operating characteristic
RSD: relative standard deviation
SD: standard deviation
S/N: signal to noise
sPLS-DA: sparse partial least squares - discriminant analysis
SVM: support vector machines
TCGA: the Cancer Genome Atlas
VIP: importance in projection score

## Funding

This work was funded by grants from Capital’s Funds for Health Improvement and Research (Grant number 2022-1-4011), National High Level Hospital Clinical Research Funding (Grant number 2022-PUMCH-A-200), and the National Natural Science Foundation of China (Grant numbers 32071436 and 31901041).

## Conflict of Interest

The authors declare no conflict of interest.

## Acknowledgements

The authors thank the patients and volunteers for their participation in the present study.

## Notes

### Competing Interest Statement

The authors have declared no competing interest.

### Funding Statement

This study was funded by Capital's Funds for Health Improvement and Research (Grant number 2022-1-4011), National High Level Hospital Clinical Research Funding (Grant number 2022-PUMCH-A-200), and the National Natural Science Foundation of China (Grant numbers 32071436 and 31901041).

### Author Declarations

Ethics committee of Peking Union Medical College Hospital gave ethical approval for this work (No. S-K2059).

