## Supplemental tables and figures for "Serum protein N-glycome patterns reveal alterations associated with endometrial cancer and its phenotypes of differentiation"

[a]Medical Research Center, State Key Laboratory of Complex Severe and Rare Diseases, Peking Union Medical College Hospital, Chinese Academy of Medical Sciences and Peking Union Medical College, Beijing 100730, China; [b]Department of General Surgery, Peking Union Medical College Hospital, Chinese Academy of Medical Sciences and Peking Union Medical College, Beijing 100730, China; [c]Department of Obstetrics and Gynecology, Peking Union Medical College Hospital, Chinese Academy of Medical Sciences and Peking Union Medical College; National Clinical Research Center for Obstetric and Gynecologic Diseases, Beijing 100730, China; [d]Department of Clinical Laboratory, Peking Union Medical College Hospital, Chinese Academy of Medical Sciences and Peking Union Medical College, Beijing 100730, China

[1]Contributed equally to this work

[*]**Correspondence**

Jinhui Wang, Department of Obstetrics and Gynecology, Peking Union Medical College Hospital, Chinese Academy of Medical Sciences and Peking Union Medical College; National Clinical Research Center for Obstetric and Gynecologic Diseases, No. 1 Shuaifuyuan Wangfujing, Dongcheng District, Beijing 100730, China.. Tel:+86-10-69156224

Zejian Zhang, Medical Research Center, State Key Laboratory of Complex Severe and Rare Diseases, Peking Union Medical College Hospital, Chinese Academy of Medical Sciences and Peking Union Medical College, No. 1 Shuaifuyuan Wangfujing, Dongcheng District, Beijing 100730, China.. Tel: +86-01-69154613

Yang Xiang, Department of Obstetrics and Gynecology, Peking Union Medical College Hospital, Chinese Academy of Medical Sciences and Peking Union Medical College; National Clinical Research Center for Obstetric and Gynecologic Diseases, No. 1 Shuaifuyuan Wangfujing, Dongcheng District, Beijing 100730, China.. Tel: +86-10-69156224

**Table of Contents**

Table S1. Overview of the directly detected N-glycans used for analysis in the present study.

Table S2. Description, depiction, and calculation of derived glycan traits.

Table S3. Data quality control.

Table S4. Associations of serum N-glycans with EC (complete list of tests performed).

Table S5. The diagnostic value of the optimized glycan panel built with the four most important (discriminative and biologically reliable) derived glycan traits using different models/algorithms.

Table S6. Associations of serum N-glycan traits and classical gynecologic tumor markers with the differentiation type (well or poorly differentiated) of EC.

Figure S1. The OPLS-DA model's permutation (cross-validation) test.

Figure S2. Scores plot of the sparse partial least squares - discriminant analysis (sPLS-DA) between endometrial cancer (EC, red circle) and healthy controls (HC, green circle) groups.

Figure S3. The S-plot of the orthogonal partial least squares - discriminant analysis (OPLS-DA).

Figure S4. The most frequently selected derived glycan traits when building the glycan panels (panel 1-panel 6, built with 2, 3, 5, 10, 20, or 33 derived glycan traits).

Figure S5. Diagnostic performance of the optimized glycan panel built with the four most important (discriminative and biologically reliable) glycan traits based on the support vector machines (SVM) model/algorithm and partial least squares (PLS) model/algorithm for endometrial cancer.

Figure S6. Diagnostic performance of the classical gynecologic tumor markers for endometrial cancer.

### Supplementary Tables

**Table S1. Overview of the directly detected N-glycans used for analysis in the present study.** Compositions were detected by MALDI-TOF-MS. The glycans were released from serum samples and the N-acetylneuraminicacids of the glycans were derivatized by ethyl esterification. All species are assigned $[M+Na]^+$. H = hexose; N = N-acetylhexosamine; F = deoxyhexose (fucose); L = lactonized N-acetylneuraminic acid (α2,3-linked); E = ethyl esterified N-acetylneuraminic acid (α2,6-linked).

| **Observed** | **Input composition** | | | | | **Modifiers** | | | **Calculated** | | **Error** | | **Used for calibration (marked with "*")** |
|---|---|---|---|---|---|---|---|---|---|---|---|---|---|
| Mass | H | N | F | L | E | $H_2O$ | Na+ | Ac | Composition | Mass | error (Da) | error (ppm) | |
| *m/z* | 162.0528234 | 203.0793725 | 146.0579088 | 273.0848518 | 319.1267166 | 18.01056468 | 22.9892207 | 42.01056 | | *m/z* | | | |
| 1257.42921 | 5 | 2 | 0 | 0 | 0 | 1 | 1 | 0 | H5N2 | 1257.42265 | 0.007 | 5.22 | |
| 1282.44957 | 3 | 3 | 1 | 0 | 0 | 1 | 1 | 0 | H3N3F1 | 1282.45428 | -0.005 | -3.68 | |
| 1339.46207 | 3 | 4 | 0 | 0 | 0 | 1 | 1 | 0 | H3N4 | 1339.47575 | -0.014 | -10.21 | |
| 1419.46371 | 6 | 2 | 0 | 0 | 0 | 1 | 1 | 0 | H6N2 | 1419.47547 | -0.012 | -8.28 | |
| 1444.51274 | 4 | 3 | 1 | 0 | 0 | 1 | 1 | 0 | H4N3F1 | 1444.50711 | 0.006 | 3.90 | |
| 1455.51697 | 3 | 3 | 0 | 0 | 1 | 1 | 1 | 0 | H3N3E1 | 1455.52309 | -0.006 | -4.20 | |
| 1460.52117 | 5 | 3 | 0 | 0 | 0 | 1 | 1 | 0 | H5N3 | 1460.50202 | 0.019 | 13.11 | |
| 1485.54111 | 3 | 4 | 1 | 0 | 0 | 1 | 1 | 0 | H3N4F1 | 1485.53365 | 0.007 | 5.02 | * |
| 1501.53349 | 4 | 4 | 0 | 0 | 0 | 1 | 1 | 0 | H4N4 | 1501.52857 | 0.005 | 3.28 | |
| 1542.54968 | 3 | 5 | 0 | 0 | 0 | 1 | 1 | 0 | H3N5 | 1542.55512 | -0.005 | -3.52 | |
| 1581.52308 | 7 | 2 | 0 | 0 | 0 | 1 | 1 | 0 | H7N2 | 1581.52829 | -0.005 | -3.29 | |
| 1601.59564 | 3 | 3 | 1 | 0 | 1 | 1 | 1 | 0 | H3N3F1E1 | 1601.58100 | 0.015 | 9.14 | |
| 1606.55678 | 5 | 3 | 1 | 0 | 0 | 1 | 1 | 0 | H5N3F1 | 1606.55993 | -0.003 | -1.96 | |
| 1617.58387 | 4 | 3 | 0 | 0 | 1 | 1 | 1 | 0 | H4N3E1 | 1617.57591 | 0.008 | 4.92 | |

| | | | | | | | | | | | | | |
|---|---|---|---|---|---|---|---|---|---|---|---|---|---|
| 1622.55001 | 6 | 3 | 0 | 0 | 0 | 1 | 1 | 0 | H6N3 | 1622.55484 | -0.005 | -2.98 | |
| 1647.59562 | 4 | 4 | 1 | 0 | 0 | 1 | 1 | 0 | H4N4F1 | 1647.58648 | 0.009 | 5.55 | * |
| 1663.59579 | 5 | 4 | 0 | 0 | 0 | 1 | 1 | 0 | H5N4 | 1663.58139 | 0.014 | 8.65 | |
| 1688.61932 | 3 | 5 | 1 | 0 | 0 | 1 | 1 | 0 | H3N5F1 | 1688.61303 | 0.006 | 3.73 | |
| 1704.60044 | 4 | 5 | 0 | 0 | 0 | 1 | 1 | 0 | H4N5 | 1704.60794 | -0.008 | -4.40 | |
| 1733.58716 | 5 | 3 | 0 | 1 | 0 | 1 | 1 | 0 | H5N3L1 | 1733.58687 | 0.000 | 0.17 | |
| 1743.57833 | 8 | 2 | 0 | 0 | 0 | 1 | 1 | 0 | H8N2 | 1743.58112 | -0.003 | -1.60 | |
| 1763.62662 | 4 | 3 | 1 | 0 | 1 | 1 | 1 | 0 | H4N3F1E1 | 1763.63382 | -0.007 | -4.08 | |
| 1774.61358 | 4 | 4 | 0 | 1 | 0 | 1 | 1 | 0 | H4N4L1 | 1774.61342 | 0.000 | 0.09 | |
| 1779.63464 | 5 | 3 | 0 | 0 | 1 | 1 | 1 | 0 | H5N3E1 | 1779.62874 | 0.006 | 3.32 | |
| 1809.64000 | 5 | 4 | 1 | 0 | 0 | 1 | 1 | 0 | H5N4F1 | 1809.63930 | 0.001 | 0.38 | * |
| 1820.64791 | 4 | 4 | 0 | 0 | 1 | 1 | 1 | 0 | H4N4E1 | 1820.65529 | -0.007 | -4.05 | |
| 1825.60996 | 6 | 4 | 0 | 0 | 0 | 1 | 1 | 0 | H6N4 | 1825.63422 | -0.024 | -13.28 | |
| 1850.66616 | 4 | 5 | 1 | 0 | 0 | 1 | 1 | 0 | H4N5F1 | 1850.66585 | 0.000 | 0.17 | |
| 1866.65007 | 5 | 5 | 0 | 0 | 0 | 1 | 1 | 0 | H5N5 | 1866.66077 | -0.011 | -5.73 | |
| 1905.63145 | 9 | 2 | 0 | 0 | 0 | 1 | 1 | 0 | H9N2 | 1905.63394 | -0.002 | -1.31 | |
| 1936.65929 | 5 | 4 | 0 | 1 | 0 | 1 | 1 | 0 | H5N4L1 | 1936.66624 | -0.007 | -3.59 | |
| 1941.67229 | 6 | 3 | 0 | 0 | 1 | 1 | 1 | 0 | H6N3E1 | 1941.68156 | -0.009 | -4.77 | |
| 1966.72426 | 4 | 4 | 1 | 0 | 1 | 1 | 1 | 0 | H4N4F1E1 | 1966.71319 | 0.011 | 5.62 | |
| 1982.71002 | 5 | 4 | 0 | 0 | 1 | 1 | 1 | 0 | H5N4E1 | 1982.70811 | 0.002 | 0.96 | * |
| 2012.72088 | 5 | 5 | 1 | 0 | 0 | 1 | 1 | 0 | H5N5F1 | 2012.71867 | 0.002 | 1.09 | |
| 2023.74760 | 4 | 5 | 0 | 0 | 1 | 1 | 1 | 0 | H4N5E1 | 2023.73466 | 0.013 | 6.40 | |
| 2082.74927 | 5 | 4 | 1 | 1 | 0 | 1 | 1 | 0 | H5N4F1L1 | 2082.72415 | 0.025 | 12.06 | |
| 2098.71588 | 6 | 4 | 0 | 1 | 0 | 1 | 1 | 0 | H6N4L1 | 2098.71907 | -0.003 | -1.52 | |
| 2110.77031 | 4 | 7 | 0 | 0 | 0 | 1 | 1 | 0 | H4N7 | 2110.76669 | 0.004 | 1.72 | |

| | | | | | | | | | | | | | |
|---|---|---|---|---|---|---|---|---|---|---|---|---|---|
| 2128.77245 | 5 | 4 | 1 | 0 | 1 | 1 | 1 | 0 | H5N4F1E1 | 2128.76602 | 0.006 | 3.02 | * |
| 2169.81455 | 4 | 5 | 1 | 0 | 1 | 1 | 1 | 0 | H4N5F1E1 | 2169.79257 | 0.022 | 10.13 | |
| 2185.79705 | 5 | 5 | 0 | 0 | 1 | 1 | 1 | 0 | H5N5E1 | 2185.78748 | 0.010 | 4.38 | |
| 2209.75876 | 5 | 4 | 0 | 2 | 0 | 1 | 1 | 0 | H5N4L2 | 2209.75110 | 0.008 | 3.47 | |
| 2255.79246 | 5 | 4 | 0 | 1 | 1 | 1 | 1 | 0 | H5N4E1L1 | 2255.79296 | 0.000 | -0.22 | |
| 2301.83999 | 5 | 4 | 0 | 0 | 2 | 1 | 1 | 0 | H5N4E2 | 2301.83483 | 0.005 | 2.25 | |
| 2331.85508 | 5 | 5 | 1 | 0 | 1 | 1 | 1 | 0 | H5N5F1E1 | 2331.84539 | 0.010 | 4.15 | |
| 2347.84336 | 6 | 5 | 0 | 0 | 1 | 1 | 1 | 0 | H6N5E1 | 2347.84031 | 0.003 | 1.30 | |
| 2355.78851 | 5 | 4 | 1 | 2 | 0 | 1 | 1 | 0 | H5N4F1L2 | 2355.80900 | -0.020 | -8.70 | |
| 2372.86451 | 4 | 6 | 1 | 0 | 1 | 1 | 1 | 0 | H4N6F1E1 | 2372.87194 | -0.007 | -3.13 | |
| 2401.85567 | 5 | 4 | 1 | 1 | 1 | 1 | 1 | 0 | H5N4F1E1L1 | 2401.85087 | 0.005 | 2.00 | |
| 2429.90998 | 4 | 7 | 0 | 0 | 1 | 1 | 1 | 0 | H4N7E1 | 2429.89340 | 0.017 | 6.82 | |
| 2447.89465 | 5 | 4 | 1 | 0 | 2 | 1 | 1 | 0 | H5N4F1E2 | 2447.89273 | 0.002 | 0.78 | |
| 2493.89248 | 6 | 5 | 1 | 0 | 1 | 1 | 1 | 0 | H6N5F1E1 | 2493.89821 | -0.006 | -2.30 | |
| 2504.91878 | 5 | 5 | 0 | 0 | 2 | 1 | 1 | 0 | H5N5E2 | 2504.91420 | 0.005 | 1.83 | |
| 2574.89939 | 6 | 5 | 0 | 2 | 0 | 1 | 1 | 0 | H6N5L2 | 2574.88329 | 0.016 | 6.25 | |
| 2604.92781 | 5 | 5 | 1 | 1 | 1 | 1 | 1 | 0 | H5N5F1E1L1 | 2604.93024 | -0.002 | -0.93 | |
| 2620.91774 | 6 | 5 | 0 | 1 | 1 | 1 | 1 | 0 | H6N5E1L1 | 2620.92516 | -0.007 | -2.83 | |
| 2650.96604 | 5 | 5 | 1 | 0 | 2 | 1 | 1 | 0 | H5N5F1E2 | 2650.97211 | -0.006 | -2.29 | |
| 2666.94791 | 6 | 5 | 0 | 0 | 2 | 1 | 1 | 0 | H6N5E2 | 2666.96702 | -0.019 | -7.16 | |
| 2720.91983 | 6 | 5 | 1 | 2 | 0 | 1 | 1 | 0 | H6N5F1L2 | 2720.94120 | -0.021 | -7.85 | |
| 2766.98562 | 6 | 5 | 1 | 1 | 1 | 1 | 1 | 0 | H6N5F1E1L1 | 2766.98307 | 0.003 | 0.92 | |
| 2813.00526 | 6 | 5 | 1 | 0 | 2 | 1 | 1 | 0 | H6N5F1E2 | 2813.02493 | -0.020 | -6.99 | |
| 2848.00774 | 8 | 6 | 2 | 0 | 0 | 1 | 1 | 0 | H8N6F2 | 2848.01443 | -0.007 | -2.35 | |
| 2894.02407 | 6 | 5 | 0 | 2 | 1 | 1 | 1 | 0 | H6N5E1L2 | 2894.01001 | 0.014 | 4.86 | |

| | | | | | | | | | | | | | |
|---|---|---|---|---|---|---|---|---|---|---|---|---|---|
| 2940.04523 | 6 | 5 | 0 | 1 | 2 | 1 | 1 | 0 | H6N5E2L1 | 2940.05187 | -0.007 | -2.26 | * |
| 2986.06421 | 6 | 5 | 0 | 0 | 3 | 1 | 1 | 0 | H6N5E3 | 2986.09374 | -0.030 | -9.89 | |
| 3040.04173 | 6 | 5 | 1 | 2 | 1 | 1 | 1 | 0 | H6N5F1E1L2 | 3040.06792 | -0.026 | -8.61 | |
| 3086.11533 | 6 | 5 | 1 | 1 | 2 | 1 | 1 | 0 | H6N5F1E2L1 | 3086.10978 | 0.006 | 1.80 | * |
| 3132.13278 | 6 | 5 | 1 | 0 | 3 | 1 | 1 | 0 | H6N5F1E3 | 3132.15165 | -0.019 | -6.02 | |
| 3232.18571 | 6 | 5 | 2 | 1 | 2 | 1 | 1 | 0 | H6N5F2E2L1 | 3232.16769 | 0.018 | 5.58 | |
| 3259.13248 | 7 | 6 | 0 | 2 | 1 | 1 | 1 | 0 | H7N6E1L2 | 3259.14220 | -0.010 | -2.98 | |
| 3305.20626 | 7 | 6 | 0 | 1 | 2 | 1 | 1 | 0 | H7N6E2L1 | 3305.18407 | 0.022 | 6.71 | |
| 3405.16982 | 7 | 6 | 1 | 2 | 1 | 1 | 1 | 0 | H7N6F1E1L2 | 3405.20011 | -0.030 | -8.90 | |
| 3451.24806 | 7 | 6 | 1 | 1 | 2 | 1 | 1 | 0 | H7N6F1E2L1 | 3451.24198 | 0.006 | 1.76 | |
| 3532.21170 | 7 | 6 | 0 | 3 | 1 | 1 | 1 | 0 | H7N6E1L3 | 3532.22706 | -0.015 | -4.35 | * |
| 3578.25147 | 7 | 6 | 0 | 2 | 2 | 1 | 1 | 0 | H7N6E2L2 | 3578.26892 | -0.017 | -4.88 | |
| 3624.30007 | 7 | 6 | 0 | 1 | 3 | 1 | 1 | 0 | H7N6E3L1 | 3624.31079 | -0.011 | -2.96 | |
| 3678.30979 | 7 | 6 | 1 | 3 | 1 | 1 | 1 | 0 | H7N6F1E1L3 | 3678.28497 | 0.025 | 6.75 | |
| 3724.32403 | 7 | 6 | 1 | 2 | 2 | 1 | 1 | 0 | H7N6F1E2L2 | 3724.32683 | -0.003 | -0.75 | |

**Table S2. Description, depiction, and calculation of derived glycan traits.** M = mannose; Hy = hybrid species; T = within total spectrum; C = within complex species; F = deoxyhexose (fucose); G = galactose; S = N-acetylneuraminic acid (sialic acid); E = α2,6-linked sialic acid; L = α2,3-linked sialic acid; H = hexose (mannose or galactose); N = N-acetylhexosamine (N-acetylglucosamine: GlcNAc).

| Derived traits | Description | Formular of calculation (derived glycan traits were calculated from the directly detected glycans) |
|---|---|---|
| **Glycan type** | | |
| **TM** | Relative abundance of high mannose type glycans within total spectrum | TM = ( H5N2 + H6N2 + H7N2 + H8N2 + H9N2 ) / ( H5N2 + H3N3F1 + H3N4 + H6N2 + H4N3F1 + H3N3E1 + H5N3 + H3N4F1 + H4N4 + H3N5 + H7N2 + H3N3F1E1 + H5N3F1 + H4N3E1 + H6N3 + H4N4F1 + H5N4 + H3N5F1 + H4N5 + H5N3L1 + H8N2 + H4N3F1E1 + H4N4L1 + H5N3E1 + H5N4F1 + H4N4E1 + H6N4 + H4N5F1 + H5N5 + H9N2 + H5N4L1 + H6N3E1 + H4N4F1E1 + H5N4E1 + H5N5F1 + H4N5E1 + H5N4F1L1 + H6N4L1 + H4N7 + H5N4F1E1 + H4N5F1E1 + H5N5E1 + H5N4L2 + H5N4E1L1 + H5N4E2 + H5N5F1E1 + H6N5E1 + H5N4F1L2 + H4N6F1E1 + H5N4F1E1L1 + H4N7E1 + H5N4F1E2 + H6N5F1E1 + H5N5E2 + H6N5L2 + H5N5F1E1L1 + H6N5E1L1 + H5N5F1E2 + H6N5E2 + H6N5F1L2 + H6N5F1E1L1 + H6N5F1E2 + H8N6F2 + H6N5E1L2 + H6N5E2L1 + H6N5E3 + H6N5F1E1L2 + H6N5F1E2L1 + H6N5F1E3 + H6N5F2E2L1 + H7N6E1L2 + H7N6E2L1 + H7N6F1E1L2 + H7N6F1E2L1 + H7N6E1L3 + H7N6E2L2 + H7N6E3L1 + H7N6F1E1L3 + H7N6F1E2L2 ) |
| **THy** | Relative abundance of hybrid type glycans within total spectrum | THy = ( H5N3 + H5N3F1 + H6N3 + H5N3L1 + H5N3E1 + H6N4 + H6N3E1 + H6N4L1 + H8N6F2 ) / ( H5N2 + H3N3F1 + H3N4 + H6N2 + H4N3F1 + H3N3E1 + H5N3 + H3N4F1 + H4N4 + H3N5 + H7N2 + H3N3F1E1 + H5N3F1 + H4N3E1 + H6N3 + H4N4F1 + H5N4 + H3N5F1 + H4N5 + H5N3L1 + H8N2 + H4N3F1E1 + H4N4L1 + H5N3E1 + H5N4F1 + H4N4E1 + H6N4 + H4N5F1 + H5N5 + H9N2 + H5N4L1 + H6N3E1 + H4N4F1E1 + H5N4E1 + H5N5F1 + H4N5E1 + H5N4F1L1 + H6N4L1 + H4N7 + H5N4F1E1 + H4N5F1E1 + H5N5E1 + H5N4L2 + H5N4E1L1 + H5N4E2 + H5N5F1E1 + H6N5E1 + H5N4F1L2 + H4N6F1E1 + H5N4F1E1L1 + H4N7E1 + H5N4F1E2 + H6N5F1E1 + H5N5E2 + H6N5L2 + H5N5F1E1L1 + H6N5E1L1 + H5N5F1E2 + H6N5E2 + H6N5F1L2 + H6N5F1E1L1 + H6N5F1E2 + H8N6F2 + H6N5E1L2 + H6N5E2L1 + H6N5E3 + H6N5F1E1L2 + H6N5F1E2L1 + H6N5F1E3 + H6N5F2E2L1 + H7N6E1L2 + H7N6E2L1 + H7N6F1E1L2 + H7N6F1E2L1 + H7N6E1L3 + H7N6E2L2 + H7N6E3L1 + H7N6F1E1L3 + H7N6F1E2L2 ) |

| | | |
|---|---|---|
| **TC** | Total complex glycans | TC = ( H3N4 + H3N3E1 + H3N4F1 + H4N4 + H3N5 + H3N3F1E1 + H4N3E1 + H4N4F1 + H5N4 + H3N5F1 + H4N5 + H4N3F1E1 + H4N4L1 + H5N4F1 + H4N4E1 + H4N5F1 + H5N5 + H5N4L1 + H4N4F1E1 + H5N4E1 + H5N5F1 + H4N5E1 + H5N4F1L1 + H4N7 + H5N4F1E1 + H4N5F1E1 + H5N5E1 + H5N4L2 + H5N4E1L1 + H5N4E2 + H5N5F1E1 + H6N5E1 + H5N4F1L2 + H4N6F1E1 + H5N4F1E1L1 + H4N7E1 + H5N4F1E2 + H6N5F1E1 + H5N5E2 + H6N5L2 + H5N5F1E1L1 + H6N5E1L1 + H5N5F1E2 + H6N5E2 + H6N5F1L2 + H6N5F1E1L1 + H6N5F1E2 + H6N5E1L2 + H6N5E2L1 + H6N5E3 + H6N5F1E1L2 + H6N5F1E2L1 + H6N5F1E3 + H6N5F2E2L1 + H7N6E1L2 + H7N6E2L1 + H7N6F1E1L2 + H7N6F1E2L1 + H7N6E1L3 + H7N6E2L2 + H7N6E3L1 + H7N6F1E1L3 + H7N6F1E2L2 ) / ( H5N2 + H3N3F1 + H3N4 + H6N2 + H4N3F1 + H3N3E1 + H5N3 + H3N4F1 + H4N4 + H3N5 + H7N2 + H3N3F1E1 + H5N3F1 + H4N3E1 + H6N3 + H4N4F1 + H5N4 + H3N5F1 + H4N5 + H5N3L1 + H8N2 + H4N3F1E1 + H4N4L1 + H5N3E1 + H5N4F1 + H4N4E1 + H6N4 + H4N5F1 + H5N5 + H9N2 + H5N4L1 + H6N3E1 + H4N4F1E1 + H5N4E1 + H5N5F1 + H4N5E1 + H5N4F1L1 + H6N4L1 + H4N7 + H5N4F1E1 + H4N5F1E1 + H5N5E1 + H5N4L2 + H5N4E1L1 + H5N4E2 + H5N5F1E1 + H6N5E1 + H5N4F1L2 + H4N6F1E1 + H5N4F1E1L1 + H4N7E1 + H5N4F1E2 + H6N5F1E1 + H5N5E2 + H6N5L2 + H5N5F1E1L1 + H6N5E1L1 + H5N5F1E2 + H6N5E2 + H6N5F1L2 + H6N5F1E1L1 + H6N5F1E2 + H8N6F2 + H6N5E1L2 + H6N5E2L1 + H6N5E3 + H6N5F1E1L2 + H6N5F1E2L1 + H6N5F1E3 + H6N5F2E2L1 + H7N6E1L2 + H7N6E2L1 + H7N6F1E1L2 + H7N6F1E2L1 + H7N6E1L3 + H7N6E2L2 + H7N6E3L1 + H7N6F1E1L3 + H7N6F1E2L2 ) |
| **MHy** | The ratio of high-mannose to hybrid glycans | MHy = ( H5N2 + H6N2 + H7N2 + H8N2 + H9N2 ) / ( H5N3 + H5N3F1 + H6N3 + H5N3L1 + H5N3E1 + H6N4 + H6N3E1 + H6N4L1 + H8N6F2 ) |
| **MM** | Average number of mannoses on high mannose type glycans | MM = ( 5 * ( H5N2 ) + 6 * ( H6N2 ) + 7 * ( H7N2 ) + 8 * ( H8N2 ) + 9 * ( H9N2 ) + 10 * (0) ) / ( H5N2 + H6N2 + H7N2 + H8N2 + H9N2 ) |
| **CA1** | Relative abundance of monoantennary glycans within complex type glycans | CA1 = ( H3N3E1 + H3N3F1E1 + H4N3E1 + H4N3F1E1 ) / ( H3N4 + H3N3E1 + H3N4F1 + H4N4 + H3N5 + H3N3F1E1 + H4N3E1 + H4N4F1 + H5N4 + H3N5F1 + H4N5 + H4N3F1E1 + H4N4L1 + H5N4F1 + H4N4E1 + H4N5F1 + H5N5 + H5N4L1 + H4N4F1E1 + H5N4E1 + H5N5F1 + H4N5E1 + H5N4F1L1 + H4N7 + H5N4F1E1 + H4N5F1E1 + H5N5E1 + H5N4L2 + H5N4E1L1 + H5N4E2 + H5N5F1E1 + H6N5E1 + H5N4F1L2 + H4N6F1E1 + H5N4F1E1L1 + H4N7E1 + H5N4F1E2 + H6N5F1E1 + H5N5E2 + H6N5L2 + H5N5F1E1L1 + H6N5E1L1 + H5N5F1E2 + H6N5E2 + H6N5F1L2 + H6N5F1E1L1 + H6N5F1E2 + H6N5E1L2 + H6N5E2L1 + H6N5E3 + H6N5F1E1L2 + H6N5F1E2L1 + H6N5F1E3 + H6N5F2E2L1 + H7N6E1L2 + H7N6E2L1 + H7N6F1E1L2 + H7N6F1E2L1 + H7N6E1L3 + H7N6E2L2 + H7N6E3L1 + H7N6F1E1L3 + H7N6F1E2L2 ) |

| | | |
|---|---|---|
| **CA2** | Relative abundance of diantennary glycans within complex type glycans | CA2 = ( H3N4 + H3N4F1 + H4N4 + H3N5 + H4N4F1 + H5N4 + H3N5F1 + H4N5 + H4N4L1 + H5N4F1 + H4N4E1 + H4N5F1 + H5N5 + H5N4L1 + H4N4F1E1 + H5N4E1 + H5N5F1 + H4N5E1 + H5N4F1L1 + H5N4F1E1 + H4N5F1E1 + H5N5E1 + H5N4L2 + H5N4E1L1 + H5N4E2 + H5N5F1E1 + H5N4F1L2 + H5N4F1E1L1 + H5N4F1E2 + H5N5E2 + H5N5F1E1L1 + H5N5F1E2 ) / ( H3N4 + H3N3E1 + H3N4F1 + H4N4 + H3N5 + H3N3F1E1 + H4N3E1 + H4N4F1 + H5N4 + H3N5F1 + H4N5 + H4N3F1E1 + H4N4L1 + H5N4F1 + H4N4E1 + H4N5F1 + H5N5 + H5N4L1 + H4N4F1E1 + H5N4E1 + H5N5F1 + H4N5E1 + H5N4F1L1 + H4N7 + H5N4F1E1 + H4N5F1E1 + H5N5E1 + H5N4L2 + H5N4E1L1 + H5N4E2 + H5N5F1E1 + H6N5E1 + H5N4F1L2 + H4N6F1E1 + H5N4F1E1L1 + H4N7E1 + H5N4F1E2 + H6N5F1E1 + H5N5E2 + H6N5L2 + H5N5F1E1L1 + H6N5E1L1 + H5N5F1E2 + H6N5E2 + H6N5F1L2 + H6N5F1E1L1 + H6N5F1E2 + H6N5E1L2 + H6N5E2L1 + H6N5E3 + H6N5F1E1L2 + H6N5F1E2L1 + H6N5F1E3 + H6N5F2E2L1 + H7N6E1L2 + H7N6E2L1 + H7N6F1E1L2 + H7N6F1E2L1 + H7N6E1L3 + H7N6E2L2 + H7N6E3L1 + H7N6F1E1L3 + H7N6F1E2L2 ) |
| **CA3** | Relative abundance of triantennary glycans within complex type glycans | CA3 = ( H6N5E1 + H6N5F1E1 + H6N5L2 + H6N5E1L1 + H6N5E2 + H6N5F1L2 + H6N5F1E1L1 + H6N5F1E2 + H6N5E1L2 + H6N5E2L1 + H6N5E3 + H6N5F1E1L2 + H6N5F1E2L1 + H6N5F1E3 + H6N5F2E2L1 ) / ( H3N4 + H3N3E1 + H3N4F1 + H4N4 + H3N5 + H3N3F1E1 + H4N3E1 + H4N4F1 + H5N4 + H3N5F1 + H4N5 + H4N3F1E1 + H4N4L1 + H5N4F1 + H4N4E1 + H4N5F1 + H5N5 + H5N4L1 + H4N4F1E1 + H5N4E1 + H5N5F1 + H4N5E1 + H5N4F1L1 + H4N7 + H5N4F1E1 + H4N5F1E1 + H5N5E1 + H5N4L2 + H5N4E1L1 + H5N4E2 + H5N5F1E1 + H6N5E1 + H5N4F1L2 + H4N6F1E1 + H5N4F1E1L1 + H4N7E1 + H5N4F1E2 + H6N5F1E1 + H5N5E2 + H6N5L2 + H5N5F1E1L1 + H6N5E1L1 + H5N5F1E2 + H6N5E2 + H6N5F1L2 + H6N5F1E1L1 + H6N5F1E2 + H6N5E1L2 + H6N5E2L1 + H6N5E3 + H6N5F1E1L2 + H6N5F1E2L1 + H6N5F1E3 + H6N5F2E2L1 + H7N6E1L2 + H7N6E2L1 + H7N6F1E1L2 + H7N6F1E2L1 + H7N6E1L3 + H7N6E2L2 + H7N6E3L1 + H7N6F1E1L3 + H7N6F1E2L2 ) |
| **CA4** | Relative abundance of tetra-antennary glycans within complex type glycans | CA4 = ( H4N6F1E1 + H7N6E1L2 + H7N6E2L1 + H7N6F1E1L2 + H7N6F1E2L1 + H7N6E1L3 + H7N6E2L2 + H7N6E3L1 + H7N6F1E1L3 + H7N6F1E2L2 ) / ( H3N4 + H3N3E1 + H3N4F1 + H4N4 + H3N5 + H3N3F1E1 + H4N3E1 + H4N4F1 + H5N4 + H3N5F1 + H4N5 + H4N3F1E1 + H4N4L1 + H5N4F1 + H4N4E1 + H4N5F1 + H5N5 + H5N4L1 + H4N4F1E1 + H5N4E1 + H5N5F1 + H4N5E1 + H5N4F1L1 + H4N7 + H5N4F1E1 + H4N5F1E1 + H5N5E1 + H5N4L2 + H5N4E1L1 + H5N4E2 + H5N5F1E1 + H6N5E1 + H5N4F1L2 + H4N6F1E1 + H5N4F1E1L1 + H4N7E1 + H5N4F1E2 + H6N5F1E1 + H5N5E2 + H6N5L2 + H5N5F1E1L1 + H6N5E1L1 + H5N5F1E2 + H6N5E2 + H6N5F1L2 + H6N5F1E1L1 + H6N5F1E2 + H6N5E1L2 + H6N5E2L1 + H6N5E3 + H6N5F1E1L2 + H6N5F1E2L1 + H6N5F1E3 + H6N5F2E2L1 + H7N6E1L2 + H7N6E2L1 + H7N6F1E1L2 + H7N6F1E2L1 + H7N6E1L3 + H7N6E2L2 + H7N6E3L1 + H7N6F1E1L3 + H7N6F1E2L2 ) |
| **TA2FS0 (IgG specific glycans)** | Total asialo fucosylated A2 | TA2FS0 = ( H3N4F1 + H4N4F1 + H3N5F1 + H5N4F1 + H4N5F1 + H5N5F1 ) |
| **Fucosylation** | | |

| | | |
|---|---|---|
| **CF** | Fucosylation within complex type glycans | CF = ( H3N4F1 + H3N3F1E1 + H4N4F1 + H3N5F1 + H4N3F1E1 + H5N4F1 + H4N5F1 + H4N4F1E1 + H5N5F1 + H5N4F1L1 + H5N4F1E1 + H4N5F1E1 + H5N5F1E1 + H5N4F1L2 + H4N6F1E1 + H5N4F1E1L1 + H5N4F1E2 + H6N5F1E1 + H5N5F1E1L1 + H5N5F1E2 + H6N5F1L2 + H6N5F1E1L1 + H6N5F1E2 + H6N5F1E1L2 + H6N5F1E2L1 + H6N5F1E3 + H6N5F2E2L1 + H7N6F1E1L2 + H7N6F1E2L1 + H7N6F1E1L3 + H7N6F1E2L2 ) / ( H3N4 + H3N3E1 + H3N4F1 + H4N4 + H3N5 + H3N3F1E1 + H4N3E1 + H4N4F1 + H5N4 + H3N5F1 + H4N5 + H4N3F1E1 + H4N4L1 + H5N4F1 + H4N4E1 + H4N5F1 + H5N5 + H5N4L1 + H4N4F1E1 + H5N4E1 + H5N5F1 + H4N5E1 + H5N4F1L1 + H4N7 + H5N4F1E1 + H4N5F1E1 + H5N5E1 + H5N4L2 + H5N4E1L1 + H5N4E2 + H5N5F1E1 + H6N5E1 + H5N4F1L2 + H4N6F1E1 + H5N4F1E1L1 + H4N7E1 + H5N4F1E2 + H6N5F1E1 + H5N5E2 + H6N5L2 + H5N5F1E1L1 + H6N5E1L1 + H5N5F1E2 + H6N5E2 + H6N5F1L2 + H6N5F1E1L1 + H6N5F1E2 + H6N5E1L2 + H6N5E2L1 + H6N5E3 + H6N5F1E1L2 + H6N5F1E2L1 + H6N5F1E3 + H6N5F2E2L1 + H7N6E1L2 + H7N6E2L1 + H7N6F1E1L2 + H7N6F1E2L1 + H7N6E1L3 + H7N6E2L2 + H7N6E3L1 + H7N6F1E1L3 + H7N6F1E2L2 ) |
| **CFa** | Relative abundance of species with 2 fucoses (i.e. at least one antennary fucose) within all complex type glycans | CFa = ( H6N5F2E2L1 ) / ( H3N4 + H3N3E1 + H3N4F1 + H4N4 + H3N5 + H3N3F1E1 + H4N3E1 + H4N4F1 + H5N4 + H3N5F1 + H4N5 + H4N3F1E1 + H4N4L1 + H5N4F1 + H4N4E1 + H4N5F1 + H5N5 + H5N4L1 + H4N4F1E1 + H5N4E1 + H5N5F1 + H4N5E1 + H5N4F1L1 + H4N7 + H5N4F1E1 + H4N5F1E1 + H5N5E1 + H5N4L2 + H5N4E1L1 + H5N4E2 + H5N5F1E1 + H6N5E1 + H5N4F1L2 + H4N6F1E1 + H5N4F1E1L1 + H4N7E1 + H5N4F1E2 + H6N5F1E1 + H5N5E2 + H6N5L2 + H5N5F1E1L1 + H6N5E1L1 + H5N5F1E2 + H6N5E2 + H6N5F1L2 + H6N5F1E1L1 + H6N5F1E2 + H6N5E1L2 + H6N5E2L1 + H6N5E3 + H6N5F1E1L2 + H6N5F1E2L1 + H6N5F1E3 + H6N5F2E2L1 + H7N6E1L2 + H7N6E2L1 + H7N6F1E1L2 + H7N6F1E2L1 + H7N6E1L3 + H7N6E2L2 + H7N6E3L1 + H7N6F1E1L3 + H7N6F1E2L2 ) |
| **A1F0** | Afucosylated monoantennary glycans | A1F0 = ( H3N3E1 + H4N3E1 ) / ( H3N3E1 + H3N3F1E1 + H4N3E1 + H4N3F1E1 ) |
| **A2F0** | Afucosylated diantennary glycans | A2F0 = ( H3N4 + H4N4 + H3N5 + H5N4 + H4N5 + H4N4L1 + H4N4E1 + H5N5 + H5N4L1 + H5N4E1 + H4N5E1 + H5N5E1 + H5N4L2 + H5N4E1L1 + H5N4E2 + H5N5E2 ) / ( H3N4 + H3N4F1 + H4N4 + H3N5 + H4N4F1 + H5N4 + H3N5F1 + H4N5 + H4N4L1 + H5N4F1 + H4N4E1 + H4N5F1 + H5N5 + H5N4L1 + H4N4F1E1 + H5N4E1 + H5N5F1 + H4N5E1 + H5N4F1L1 + H5N4F1E1 + H4N5F1E1 + H5N5E1 + H5N4L2 + H5N4E1L1 + H5N4E2 + H5N5F1E1 + H5N4F1L2 + H5N4F1E1L1 + H5N4F1E2 + H5N5E2 + H5N5F1E1L1 + H5N5F1E2 ) |
| **A3F0** | Afucosylated triantennary glycans | A3F0 = ( H6N5E1 + H6N5L2 + H6N5E1L1 + H6N5E2 + H6N5E1L2 + H6N5E2L1 + H6N5E3 ) / ( H6N5E1 + H6N5F1E1 + H6N5L2 + H6N5E1L1 + H6N5E2 + H6N5F1L2 + H6N5F1E1L1 + H6N5F1E2 + H6N5E1L2 + H6N5E2L1 + H6N5E3 + H6N5F1E1L2 + H6N5F1E2L1 + H6N5F1E3 + H6N5F2E2L1 ) |
| **A4F0** | Afucosylated tetra-antennary glycans | A4F0 = ( H7N6E1L2 + H7N6E2L1 + H7N6E1L3 + H7N6E2L2 + H7N6E3L1 ) / ( H4N6F1E1 + H7N6E1L2 + H7N6E2L1 + H7N6F1E1L2 + H7N6F1E2L1 + H7N6E1L3 + H7N6E2L2 + H7N6E3L1 + H7N6F1E1L3 + H7N6F1E2L2 ) |
| **A1F** | Fucosylation within monoantennary glycans | A1F = ( H3N3F1E1 + H4N3F1E1 ) / ( H3N3E1 + H3N3F1E1 + H4N3E1 + H4N3F1E1 ) |

| | | |
|---|---|---|
| **A2F** | Fucosylation within diantennary glycans | A2F = ( H3N4F1 + H4N4F1 + H3N5F1 + H5N4F1 + H4N5F1 + H4N4F1E1 + H5N5F1 + H5N4F1L1 + H5N4F1E1 + H4N5F1E1 + H5N5F1E1 + H5N4F1L2 + H5N4F1E1L1 + H5N4F1E2 + H5N5F1E1L1 + H5N5F1E2 ) / ( H3N4 + H3N4F1 + H4N4 + H3N5 + H4N4F1 + H5N4 + H3N5F1 + H4N5 + H4N4L1 + H5N4F1 + H4N4E1 + H4N5F1 + H5N5 + H5N4L1 + H4N4F1E1 + H5N4E1 + H5N5F1 + H4N5E1 + H5N4F1L1 + H5N4F1E1 + H4N5F1E1 + H5N5E1 + H5N4L2 + H5N4E1L1 + H5N4E2 + H5N5F1E1 + H5N4F1L2 + H5N4F1E1L1 + H5N4F1E2 + H5N5E2 + H5N5F1E1L1 + H5N5F1E2 ) |
| **A3F** | Fucosylation within triantennary glycans | A3F = ( H6N5F1E1 + H6N5F1L2 + H6N5F1E1L1 + H6N5F1E2 + H6N5F1E1L2 + H6N5F1E2L1 + H6N5F1E3 + H6N5F2E2L1 ) / ( H6N5E1 + H6N5F1E1 + H6N5L2 + H6N5E1L1 + H6N5E2 + H6N5F1L2 + H6N5F1E1L1 + H6N5F1E2 + H6N5E1L2 + H6N5E2L1 + H6N5E3 + H6N5F1E1L2 + H6N5F1E2L1 + H6N5F1E3 + H6N5F2E2L1 ) |
| **A4F** | Fucosylation within tetra-antennary glycans | A4F = ( H4N6F1E1 + H7N6F1E1L2 + H7N6F1E2L1 + H7N6F1E1L3 + H7N6F1E2L2 ) / ( H4N6F1E1 + H7N6E1L2 + H7N6E2L1 + H7N6F1E1L2 + H7N6F1E2L1 + H7N6E1L3 + H7N6E2L2 + H7N6E3L1 + H7N6F1E1L3 + H7N6F1E2L2 ) |
| **A3Fa** | Relative abundance of species with 2 fucoses (i.e. at least one antennary fucose) within triantennary glycans | A3Fa = ( H6N5F2E2L1 ) / ( H6N5E1 + H6N5F1E1 + H6N5L2 + H6N5E1L1 + H6N5E2 + H6N5F1L2 + H6N5F1E1L1 + H6N5F1E2 + H6N5E1L2 + H6N5E2L1 + H6N5E3 + H6N5F1E1L2 + H6N5F1E2L1 + H6N5F1E3 + H6N5F2E2L1 ) |
| **A2S0F** | Fucosylation within non-sialylated diantennary glycans | A2S0F = ( H3N4F1 + H4N4F1 + H3N5F1 + H5N4F1 + H4N5F1 + H5N5F1 ) / ( H3N4 + H3N4F1 + H4N4 + H3N5 + H4N4F1 + H5N4 + H3N5F1 + H4N5 + H5N4F1 + H4N5F1 + H5N5 + H5N5F1 ) |
| **A1L0F** | Fucosylation within monoantennary glycans without α2,3-linked sialic acid | A1L0F = ( H3N3F1E1 + H4N3F1E1 ) / ( H3N3E1 + H3N3F1E1 + H4N3E1 + H4N3F1E1 ) |
| **A2L0F** | Fucosylation within diantennary glycans without α2,3-linked sialic acid | A2L0F = ( H3N4F1 + H4N4F1 + H3N5F1 + H5N4F1 + H4N5F1 + H4N4F1E1 + H5N5F1 + H5N4F1E1 + H4N5F1E1 + H5N5F1E1 + H5N4F1E2 + H5N5F1E2 ) / ( H3N4 + H3N4F1 + H4N4 + H3N5 + H4N4F1 + H5N4 + H3N5F1 + H4N5 + H5N4F1 + H4N4E1 + H4N5F1 + H5N5 + H4N4F1E1 + H5N4E1 + H5N5F1 + H4N5E1 + H5N4F1E1 + H4N5F1E1 + H5N5E1 + H5N4E2 + H5N5F1E1 + H5N4F1E2 + H5N5E2 + H5N5F1E2 ) |
| **A3L0F** | Fucosylation within triantennary glycans without α2,3-linked sialic acid | A3L0F = ( H6N5F1E1 + H6N5F1E2 + H6N5F1E3 ) / ( H6N5E1 + H6N5F1E1 + H6N5E2 + H6N5F1E2 + H6N5E3 + H6N5F1E3 ) |

| | | |
|---|---|---|
| **A2E0F** | Fucosylation within diantennary glycans without α2,6-linked sialic acid | A2E0F = ( H3N4F1 + H4N4F1 + H3N5F1 + H5N4F1 + H4N5F1 + H5N5F1 + H5N4F1L1 + H5N4F1L2 ) / ( H3N4 + H3N4F1 + H4N4 + H3N5 + H4N4F1 + H5N4 + H3N5F1 + H4N5 + H4N4L1 + H5N4F1 + H4N5F1 + H5N5 + H5N4L1 + H5N5F1 + H5N4F1L1 + H5N4L2 + H5N4F1L2 ) |
| **A3E0F** | Fucosylation within triantennary glycans without α2,6-linked sialic acid | A3E0F = ( H6N5F1L2 ) / ( H6N5L2 + H6N5F1L2 ) |
| **A1SF** | Fucosylation within sialylated monoantennary glycans | A1SF = ( H3N3F1E1 + H4N3F1E1 ) / ( H3N3E1 + H3N3F1E1 + H4N3E1 + H4N3F1E1 ) |
| **A2SF** | Fucosylation within sialylated diantennary glycans | A2SF = ( H4N4F1E1 + H5N4F1L1 + H5N4F1E1 + H4N5F1E1 + H5N5F1E1 + H5N4F1L2 + H5N4F1E1L1 + H5N4F1E2 + H5N5F1E1L1 + H5N5F1E2 ) / ( H4N4L1 + H4N4E1 + H5N4L1 + H4N4F1E1 + H5N4E1 + H4N5E1 + H5N4F1L1 + H5N4F1E1 + H4N5F1E1 + H5N5E1 + H5N4L2 + H5N4E1L1 + H5N4E2 + H5N5F1E1 + H5N4F1L2 + H5N4F1E1L1 + H5N4F1E2 + H5N5E2 + H5N5F1E1L1 + H5N5F1E2 ) |
| **A3SF** | Fucosylation within sialylated triantennary glycans | A3SF = ( H6N5F1E1 + H6N5F1L2 + H6N5F1E1L1 + H6N5F1E2 + H6N5F1E1L2 + H6N5F1E2L1 + H6N5F1E3 + H6N5F2E2L1 ) / ( H6N5E1 + H6N5F1E1 + H6N5L2 + H6N5E1L1 + H6N5E2 + H6N5F1L2 + H6N5F1E1L1 + H6N5F1E2 + H6N5E1L2 + H6N5E2L1 + H6N5E3 + H6N5F1E1L2 + H6N5F1E2L1 + H6N5F1E3 + H6N5F2E2L1 ) |
| **A4SF** | Fucosylation within sialylated tetra-antennary glycans | A4SF = ( H4N6F1E1 + H7N6F1E1L2 + H7N6F1E2L1 + H7N6F1E1L3 + H7N6F1E2L2 ) / ( H4N6F1E1 + H7N6E1L2 + H7N6E2L1 + H7N6F1E1L2 + H7N6F1E2L1 + H7N6E1L3 + H7N6E2L2 + H7N6E3L1 + H7N6F1E1L3 + H7N6F1E2L2 ) |
| **A2LF** | Fucosylation within diantennary glycans with α2,3-linked sialic acid | A2LF = ( H5N4F1L1 + H5N4F1L2 + H5N4F1E1L1 + H5N5F1E1L1 ) / ( H4N4L1 + H5N4L1 + H5N4F1L1 + H5N4L2 + H5N4E1L1 + H5N4F1L2 + H5N4F1E1L1 + H5N5F1E1L1 ) |
| **A3LF** | Fucosylation within triantennary glycans with α2,3-linked sialic acid | A3LF = ( H6N5F1L2 + H6N5F1E1L1 + H6N5F1E1L2 + H6N5F1E2L1 + H6N5F2E2L1 ) / ( H6N5L2 + H6N5E1L1 + H6N5F1L2 + H6N5F1E1L1 + H6N5E1L2 + H6N5E2L1 + H6N5F1E1L2 + H6N5F1E2L1 + H6N5F2E2L1 ) |
| **A4LF** | Fucosylation within tetra-antennary glycans with α2,3-linked sialic acid | A4LF = ( H7N6F1E1L2 + H7N6F1E2L1 + H7N6F1E1L3 + H7N6F1E2L2 ) / ( H7N6E1L2 + H7N6E2L1 + H7N6F1E1L2 + H7N6F1E2L1 + H7N6E1L3 + H7N6E2L2 + H7N6E3L1 + H7N6F1E1L3 + H7N6F1E2L2 ) |

| | | |
|---|---|---|
| **A1EF** | Fucosylation within monoantennary glycans with α2,6-linked sialic acid | A1EF = ( H3N3F1E1 + H4N3F1E1 ) / ( H3N3E1 + H3N3F1E1 + H4N3E1 + H4N3F1E1 ) |
| **A2EF** | Fucosylation within diantennary glycans with α2,6-linked sialic acid | A2EF = ( H4N4F1E1 + H5N4F1E1 + H4N5F1E1 + H5N5F1E1 + H5N4F1E1L1 + H5N4F1E2 + H5N5F1E1L1 + H5N5F1E2 ) / ( H4N4E1 + H4N4F1E1 + H5N4E1 + H4N5E1 + H5N4F1E1 + H4N5F1E1 + H5N5E1 + H5N4E1L1 + H5N4E2 + H5N5F1E1 + H5N4F1E1L1 + H5N4F1E2 + H5N5E2 + H5N5F1E1L1 + H5N5F1E2 ) |
| **A3EF** | Fucosylation within triantennary glycans with α2,6-linked sialic acid | A3EF = ( H6N5F1E1 + H6N5F1E1L1 + H6N5F1E2 + H6N5F1E1L2 + H6N5F1E2L1 + H6N5F1E3 + H6N5F2E2L1 ) / ( H6N5E1 + H6N5F1E1 + H6N5E1L1 + H6N5E2 + H6N5F1E1L1 + H6N5F1E2 + H6N5E1L2 + H6N5E2L1 + H6N5E3 + H6N5F1E1L2 + H6N5F1E2L1 + H6N5F1E3 + H6N5F2E2L1 ) |
| **A4EF** | Fucosylation within tetra-antennary glycans with α2,6-linked sialic acid | A4EF = ( H4N6F1E1 + H7N6F1E1L2 + H7N6F1E2L1 + H7N6F1E1L3 + H7N6F1E2L2 ) / ( H4N6F1E1 + H7N6E1L2 + H7N6E2L1 + H7N6F1E1L2 + H7N6F1E2L1 + H7N6E1L3 + H7N6E2L2 + H7N6E3L1 + H7N6F1E1L3 + H7N6F1E2L2 ) |
| **Bisection** | | |
| **CB** | Relative abundance of species with a bisecting GlcNAc within all complex glycans | CB = ( H3N5 + H3N5F1 + H4N5 + H4N5F1 + H5N5 + H5N5F1 + H4N5E1 + H4N5F1E1 + H5N5E1 + H5N5F1E1 + H5N5E2 + H5N5F1E1L1 + H5N5F1E2 ) / ( H3N4 + H3N3E1 + H3N4F1 + H4N4 + H3N5 + H3N3F1E1 + H4N3E1 + H4N4F1 + H5N4 + H3N5F1 + H4N5 + H4N3F1E1 + H4N4L1 + H5N4F1 + H4N4E1 + H4N5F1 + H5N5 + H5N4L1 + H4N4F1E1 + H5N4E1 + H5N5F1 + H4N5E1 + H5N4F1L1 + H4N7 + H5N4F1E1 + H4N5F1E1 + H5N5E1 + H5N4L2 + H5N4E1L1 + H5N4E2 + H5N5F1E1 + H6N5E1 + H5N4F1L2 + H4N6F1E1 + H5N4F1E1L1 + H4N7E1 + H5N4F1E2 + H6N5F1E1 + H5N5E2 + H6N5L2 + H5N5F1E1L1 + H6N5E1L1 + H5N5F1E2 + H6N5E2 + H6N5F1L2 + H6N5F1E1L1 + H6N5F1E2 + H6N5E1L2 + H6N5E2L1 + H6N5E3 + H6N5F1E1L2 + H6N5F1E2L1 + H6N5F1E3 + H6N5F2E2L1 + H7N6E1L2 + H7N6E2L1 + H7N6F1E1L2 + H7N6F1E2L1 + H7N6E1L3 + H7N6E2L2 + H7N6E3L1 + H7N6F1E1L3 + H7N6F1E2L2 ) |
| **A2B** | Relative abundance of species with a bisecting GlcNAc within diantennary glycans | A2B = ( H3N5 + H3N5F1 + H4N5 + H4N5F1 + H5N5 + H5N5F1 + H4N5E1 + H4N5F1E1 + H5N5E1 + H5N5F1E1 + H5N5E2 + H5N5F1E1L1 + H5N5F1E2 ) / ( H3N4 + H3N4F1 + H4N4 + H3N5 + H4N4F1 + H5N4 + H3N5F1 + H4N5 + H4N4L1 + H5N4F1 + H4N4E1 + H4N5F1 + H5N5 + H5N4L1 + H4N4F1E1 + H5N4E1 + H5N5F1 + H4N5E1 + H5N4F1L1 + H5N4F1E1 + H4N5F1E1 + H5N5E1 + H5N4L2 + H5N4E1L1 + H5N4E2 + H5N5F1E1 + H5N4F1L2 + H5N4F1E1L1 + H5N4F1E2 + H5N5E2 + H5N5F1E1L1 + H5N5F1E2 ) |

| | | |
|---|---|---|
| **A2F0B** | Relative abundance of species with a bisecting GlcNAc within non-fucosylated diantennary glycans | A2F0B = ( H3N5 + H4N5 + H5N5 + H4N5E1 + H5N5E1 + H5N5E2 ) / ( H3N4 + H4N4 + H3N5 + H5N4 + H4N5 + H4N4L1 + H4N4E1 + H5N5 + H5N4L1 + H5N4E1 + H4N5E1 + H5N5E1 + H5N4L2 + H5N4E1L1 + H5N4E2 + H5N5E2 ) |
| **A2FB** | Relative abundance of species with a bisecting GlcNAc within fucosylated diantennary | A2FB = ( H3N5F1 + H4N5F1 + H5N5F1 + H4N5F1E1 + H5N5F1E1 + H5N5F1E1L1 + H5N5F1E2 ) / ( H3N4F1 + H4N4F1 + H3N5F1 + H5N4F1 + H4N5F1 + H4N4F1E1 + H5N5F1 + H5N4F1L1 + H5N4F1E1 + H4N5F1E1 + H5N5F1E1 + H5N4F1L2 + H5N4F1E1L1 + H5N4F1E2 + H5N5F1E1L1 + H5N5F1E2 ) |
| **A2S0B** | Relative abundance of species with a bisecting GlcNAc within non-sialylated diantennary glycans | A2S0B = ( H3N5 + H3N5F1 + H4N5 + H4N5F1 + H5N5 + H5N5F1 ) / ( H3N4 + H3N4F1 + H4N4 + H3N5 + H4N4F1 + H5N4 + H3N5F1 + H4N5 + H5N4F1 + H4N5F1 + H5N5 + H5N5F1 ) |
| **A2SB** | Relative abundance of species with a bisecting GlcNAc within sialylated diantennary glycans | A2SB = ( H4N5E1 + H4N5F1E1 + H5N5E1 + H5N5F1E1 + H5N5E2 + H5N5F1E1L1 + H5N5F1E2 ) / ( H4N4L1 + H4N4E1 + H5N4L1 + H4N4F1E1 + H5N4E1 + H4N5E1 + H5N4F1L1 + H5N4F1E1 + H4N5F1E1 + H5N5E1 + H5N4L2 + H5N4E1L1 + H5N4E2 + H5N5F1E1 + H5N4F1L2 + H5N4F1E1L1 + H5N4F1E2 + H5N5E2 + H5N5F1E1L1 + H5N5F1E2 ) |
| **A2F0S0B** | Relative abundance of species with a bisecting GlcNAc within non-fucosylated non-sialylated diantennary glycans | A2F0S0B = ( H3N5 + H4N5 + H5N5 ) / ( H3N4 + H4N4 + H3N5 + H5N4 + H4N5 + H5N5 ) |
| **A2F0SB** | Relative abundance of species with a bisecting GlcNAc within non-fucosylated sialylated diantennary glycans | A2F0SB = ( H4N5E1 + H5N5E1 + H5N5E2 ) / ( H4N4L1 + H4N4E1 + H5N4L1 + H5N4E1 + H4N5E1 + H5N5E1 + H5N4L2 + H5N4E1L1 + H5N4E2 + H5N5E2 ) |

| | | |
|---|---|---|
| **A2FS0B** | Relative abundance of species with a bisecting GlcNAc within fucosylated non-sialylated diantennary glycans | A2FS0B = ( H3N5F1 + H4N5F1 + H5N5F1 ) / ( H3N4F1 + H4N4F1 + H3N5F1 + H5N4F1 + H4N5F1 + H5N5F1 ) |
| **A2FSB** | Relative abundance of species with a bisecting GlcNAc within fucosylated sialylated diantennary glycans | A2FSB = ( H4N5F1E1 + H5N5F1E1 + H5N5F1E1L1 + H5N5F1E2 ) / ( H4N4F1E1 + H5N4F1L1 + H5N4F1E1 + H4N5F1E1 + H5N5F1E1 + H5N4F1L2 + H5N4F1E1L1 + H5N4F1E2 + H5N5F1E1L1 + H5N5F1E2 ) |
| **Galactosylation** | | |
| **CG** | Galactosylation within all complex glycans | CG = ( H3N3E1 + H4N4 + H3N3F1E1 + H4N3E1 + H4N4F1 + H5N4 + H4N5 + H4N3F1E1 + H4N4L1 + H5N4F1 + H4N4E1 + H4N5F1 + H5N5 + H5N4L1 + H4N4F1E1 + H5N4E1 + H5N5F1 + H4N5E1 + H5N4F1L1 + H5N4F1E1 + H4N5F1E1 + H5N5E1 + H5N4L2 + H5N4E1L1 + H5N4E2 + H5N5F1E1 + H6N5E1 + H5N4F1L2 + H4N6F1E1 + H5N4F1E1L1 + H4N7E1 + H5N4F1E2 + H6N5F1E1 + H5N5E2 + H6N5L2 + H5N5F1E1L1 + H6N5E1L1 + H5N5F1E2 + H6N5E2 + H6N5F1L2 + H6N5F1E1L1 + H6N5F1E2 + H6N5E1L2 + H6N5E2L1 + H6N5E3 + H6N5F1E1L2 + H6N5F1E2L1 + H6N5F1E3 + H6N5F2E2L1 + H7N6E1L2 + H7N6E2L1 + H7N6F1E1L2 + H7N6F1E2L1 + H7N6E1L3 + H7N6E2L2 + H7N6E3L1 + H7N6F1E1L3 + H7N6F1E2L2 ) / ( H3N4 + H3N3E1 + H3N4F1 + H4N4 + H3N5 + H3N3F1E1 + H4N3E1 + H4N4F1 + H5N4 + H3N5F1 + H4N5 + H4N3F1E1 + H4N4L1 + H5N4F1 + H4N4E1 + H4N5F1 + H5N5 + H5N4L1 + H4N4F1E1 + H5N4E1 + H5N5F1 + H4N5E1 + H5N4F1L1 + H4N7 + H5N4F1E1 + H4N5F1E1 + H5N5E1 + H5N4L2 + H5N4E1L1 + H5N4E2 + H5N5F1E1 + H6N5E1 + H5N4F1L2 + H4N6F1E1 + H5N4F1E1L1 + H4N7E1 + H5N4F1E2 + H6N5F1E1 + H5N5E2 + H6N5L2 + H5N5F1E1L1 + H6N5E1L1 + H5N5F1E2 + H6N5E2 + H6N5F1L2 + H6N5F1E1L1 + H6N5F1E2 + H6N5E1L2 + H6N5E2L1 + H6N5E3 + H6N5F1E1L2 + H6N5F1E2L1 + H6N5F1E3 + H6N5F2E2L1 + H7N6E1L2 + H7N6E2L1 + H7N6F1E1L2 + H7N6F1E2L1 + H7N6E1L3 + H7N6E2L2 + H7N6E3L1 + H7N6F1E1L3 + H7N6F1E2L2 ) |
| **A2G** | Galactosylation per antenna within diantennary glycans | A2G = ( 0/2 * ( H3N4 + H3N4F1 + H3N5 + H3N5F1 ) + 1/2 * ( H4N4 + H4N4F1 + H4N5 + H4N4L1 + H4N4E1 + H4N5F1 + H4N4F1E1 + H4N5E1 + H4N5F1E1 ) + 2/2 * ( H5N4 + H5N4F1 + H5N5 + H5N4L1 + H5N4E1 + H5N5F1 + H5N4F1L1 + H5N4F1E1 + H5N5E1 + H5N4L2 + H5N4E1L1 + H5N4E2 + H5N5F1E1 + H5N4F1L2 + H5N4F1E1L1 + H5N4F1E2 + H5N5E2 + H5N5F1E1L1 + H5N5F1E2 ) ) / ( H3N4 + H3N4F1 + H4N4 + H3N5 + H4N4F1 + H5N4 + H3N5F1 + H4N5 + H4N4L1 + H5N4F1 + H4N4E1 + H4N5F1 + H5N5 + H5N4L1 + H4N4F1E1 + H5N4E1 + H5N5F1 + H4N5E1 + H5N4F1L1 + H5N4F1E1 + H4N5F1E1 + H5N5E1 + H5N4L2 + H5N4E1L1 + H5N4E2 + H5N5F1E1 + H5N4F1L2 + H5N4F1E1L1 + H5N4F1E2 + H5N5E2 + H5N5F1E1L1 + H5N5F1E2 ) |

| | | |
|---|---|---|
| **A4G** | Galactosylation per antenna within tetra-antennary glycans | A4G = ( 0/4 * (0) + 1/4 * ( H4N6F1E1 ) + 2/4 * (0) + 3/4 * (0) + 4/4 * ( H7N6E1L2 + H7N6E2L1 + H7N6F1E1L2 + H7N6F1E2L1 + H7N6E1L3 + H7N6E2L2 + H7N6E3L1 + H7N6F1E1L3 + H7N6F1E2L2 ) ) / ( H4N6F1E1 + H7N6E1L2 + H7N6E2L1 + H7N6F1E1L2 + H7N6F1E2L1 + H7N6E1L3 + H7N6E2L2 + H7N6E3L1 + H7N6F1E1L3 + H7N6F1E2L2 ) |
| **A2F0G** | Galactosylation per antenna within non-fucosylated diantennary glycans | A2F0G = ( 0/2 * ( H3N4 + H3N5 ) + 1/2 * ( H4N4 + H4N5 + H4N4L1 + H4N4E1 + H4N5E1 ) + 2/2 * ( H5N4 + H5N5 + H5N4L1 + H5N4E1 + H5N5E1 + H5N4L2 + H5N4E1L1 + H5N4E2 + H5N5E2 ) ) / ( H3N4 + H4N4 + H3N5 + H5N4 + H4N5 + H4N4L1 + H4N4E1 + H5N5 + H5N4L1 + H5N4E1 + H4N5E1 + H5N5E1 + H5N4L2 + H5N4E1L1 + H5N4E2 + H5N5E2 ) |
| **A4F0G** | Galactosylation per antenna within non-fucosylated tetra-antennary glycans | A4F0G = ( 0/4 * (0) + 1/4 * (0) + 2/4 * (0) + 3/4 * (0) + 4/4 * ( H7N6E1L2 + H7N6E2L1 + H7N6E1L3 + H7N6E2L2 + H7N6E3L1 ) ) / ( H4N6F1E1 + H7N6E1L2 + H7N6E2L1 + H7N6F1E1L2 + H7N6F1E2L1 + H7N6E1L3 + H7N6E2L2 + H7N6E3L1 + H7N6F1E1L3 + H7N6F1E2L2 ) |
| **A2FG** | Galactosylation per antenna within fucosylated diantennary glycans | A2FG = ( 0/2 * ( H3N4F1 + H3N5F1 ) + 1/2 * ( H4N4F1 + H4N5F1 + H4N4F1E1 + H4N5F1E1 ) + 2/2 * ( H5N4F1 + H5N5F1 + H5N4F1L1 + H5N4F1E1 + H5N5F1E1 + H5N4F1L2 + H5N4F1E1L1 + H5N4F1E2 + H5N5F1E1L1 + H5N5F1E2 ) ) / ( H3N4F1 + H4N4F1 + H3N5F1 + H5N4F1 + H4N5F1 + H4N4F1E1 + H5N5F1 + H5N4F1L1 + H5N4F1E1 + H4N5F1E1 + H5N5F1E1 + H5N4F1L2 + H5N4F1E1L1 + H5N4F1E2 + H5N5F1E1L1 + H5N5F1E2 ) |
| **A4FG** | Galactosylation per antenna within fucosylated tetra-antennary glycans | A4FG = ( 0/4 * (0) + 1/4 * ( H4N6F1E1 ) + 2/4 * (0) + 3/4 * (0) + 4/4 * ( H7N6F1E1L2 + H7N6F1E2L1 + H7N6F1E1L3 + H7N6F1E2L2 ) ) / ( H4N6F1E1 + H7N6E1L2 + H7N6E2L1 + H7N6F1E1L2 + H7N6F1E2L1 + H7N6E1L3 + H7N6E2L2 + H7N6E3L1 + H7N6F1E1L3 + H7N6F1E2L2 ) |
| **A2S0G** | Galactosylation per antenna within non-sialylated diantennary glycans | A2S0G = ( 0/2 * ( H3N4 + H3N4F1 + H3N5 + H3N5F1 ) + 1/2 * ( H4N4 + H4N4F1 + H4N5 + H4N5F1 ) + 2/2 * ( H5N4 + H5N4F1 + H5N5 + H5N5F1 ) ) / ( H3N4 + H3N4F1 + H4N4 + H3N5 + H4N4F1 + H5N4 + H3N5F1 + H4N5 + H5N4F1 + H4N5F1 + H5N5 + H5N5F1 ) |
| **A2SG** | Galactosylation per antenna within sialylated diantennary glycans | A2SG = ( 0/2 * (0) + 1/2 * ( H4N4L1 + H4N4E1 + H4N4F1E1 + H4N5E1 + H4N5F1E1 ) + 2/2 * ( H5N4L1 + H5N4E1 + H5N4F1L1 + H5N4F1E1 + H5N5E1 + H5N4L2 + H5N4E1L1 + H5N4E2 + H5N5F1E1 + H5N4F1L2 + H5N4F1E1L1 + H5N4F1E2 + H5N5E2 + H5N5F1E1L1 + H5N5F1E2 ) ) / ( H4N4L1 + H4N4E1 + H5N4L1 + H4N4F1E1 + H5N4E1 + H4N5E1 + H5N4F1L1 + H5N4F1E1 + H4N5F1E1 + H5N5E1 + H5N4L2 + H5N4E1L1 + H5N4E2 + H5N5F1E1 + H5N4F1L2 + H5N4F1E1L1 + H5N4F1E2 + H5N5E2 + H5N5F1E1L1 + H5N5F1E2 ) |
| **A2F0S0G** | Galactosylation per antenna within non-fucosylated, non-sialylated diantennary glycans | A2F0S0G = ( 0/2 * ( H3N4 + H3N5 ) + 1/2 * ( H4N4 + H4N5 ) + 2/2 * ( H5N4 + H5N5 ) ) / ( H3N4 + H4N4 + H3N5 + H5N4 + H4N5 + H5N5 ) |

| | | |
|---|---|---|
| **A2FS0G** | Galactosylation per antenna within fucosylated non-sialylated diantennary glycans | A2FS0G = ( 0/2 * ( H3N4F1 + H3N5F1 ) + 1/2 * ( H4N4F1 + H4N5F1 ) + 2/2 * ( H5N4F1 + H5N5F1 ) ) / ( H3N4F1 + H4N4F1 + H3N5F1 + H5N4F1 + H4N5F1 + H5N5F1 ) |
| **A2F0SG** | Galactosylation per antenna within non-fucosylated sialylated diantennary glycans | A2F0SG = ( 0/2 * (0) + 1/2 * ( H4N4L1 + H4N4E1 + H4N5E1 ) + 2/2 * ( H5N4L1 + H5N4E1 + H5N5E1 + H5N4L2 + H5N4E1L1 + H5N4E2 + H5N5E2 ) ) / ( H4N4L1 + H4N4E1 + H5N4L1 + H5N4E1 + H4N5E1 + H5N5E1 + H5N4L2 + H5N4E1L1 + H5N4E2 + H5N5E2 ) |
| **A2FSG** | Galactosylation per antenna within fucosylated sialylated diantennary glycans | A2FSG = ( 0/2 * (0) + 1/2 * ( H4N4F1E1 + H4N5F1E1 ) + 2/2 * ( H5N4F1L1 + H5N4F1E1 + H5N5F1E1 + H5N4F1L2 + H5N4F1E1L1 + H5N4F1E2 + H5N5F1E1L1 + H5N5F1E2 ) ) / ( H4N4F1E1 + H5N4F1L1 + H5N4F1E1 + H4N5F1E1 + H5N5F1E1 + H5N4F1L2 + H5N4F1E1L1 + H5N4F1E2 + H5N5F1E1L1 + H5N5F1E2 ) |
| **Sialylation** | | |
| **CS** | Sialylation per antenna within all complex glycans | CS = ( H3N3E1 + H3N3F1E1 + H4N3E1 + H4N3F1E1 + H4N4L1 + H4N4E1 + H5N4L1 + H4N4F1E1 + H5N4E1 + H4N5E1 + H5N4F1L1 + H5N4F1E1 + H4N5F1E1 + H5N5E1 + H5N4L2 + H5N4E1L1 + H5N4E2 + H5N5F1E1 + H6N5E1 + H5N4F1L2 + H4N6F1E1 + H5N4F1E1L1 + H4N7E1 + H5N4F1E2 + H6N5F1E1 + H5N5E2 + H6N5L2 + H5N5F1E1L1 + H6N5E1L1 + H5N5F1E2 + H6N5E2 + H6N5F1L2 + H6N5F1E1L1 + H6N5F1E2 + H6N5E1L2 + H6N5E2L1 + H6N5E3 + H6N5F1E1L2 + H6N5F1E2L1 + H6N5F1E3 + H6N5F2E2L1 + H7N6E1L2 + H7N6E2L1 + H7N6F1E1L2 + H7N6F1E2L1 + H7N6E1L3 + H7N6E2L2 + H7N6E3L1 + H7N6F1E1L3 + H7N6F1E2L2 ) / ( H3N4 + H3N3E1 + H3N4F1 + H4N4 + H3N5 + H3N3F1E1 + H4N3E1 + H4N4F1 + H5N4 + H3N5F1 + H4N5 + H4N3F1E1 + H4N4L1 + H5N4F1 + H4N4E1 + H4N5F1 + H5N5 + H5N4L1 + H4N4F1E1 + H5N4E1 + H5N5F1 + H4N5E1 + H5N4F1L1 + H4N7 + H5N4F1E1 + H4N5F1E1 + H5N5E1 + H5N4L2 + H5N4E1L1 + H5N4E2 + H5N5F1E1 + H6N5E1 + H5N4F1L2 + H4N6F1E1 + H5N4F1E1L1 + H4N7E1 + H5N4F1E2 + H6N5F1E1 + H5N5E2 + H6N5L2 + H5N5F1E1L1 + H6N5E1L1 + H5N5F1E2 + H6N5E2 + H6N5F1L2 + H6N5F1E1L1 + H6N5F1E2 + H6N5E1L2 + H6N5E2L1 + H6N5E3 + H6N5F1E1L2 + H6N5F1E2L1 + H6N5F1E3 + H6N5F2E2L1 + H7N6E1L2 + H7N6E2L1 + H7N6F1E1L2 + H7N6F1E2L1 + H7N6E1L3 + H7N6E2L2 + H7N6E3L1 + H7N6F1E1L3 + H7N6F1E2L2 ) |

| | | |
|---|---|---|
| **A2S** | Sialylation per antenna within diantennary glycans | A2S = ( 0/2 * ( H3N4 + H3N4F1 + H4N4 + H3N5 + H4N4F1 + H5N4 + H3N5F1 + H4N5 + H5N4F1 + H4N5F1 + H5N5 + H5N5F1 ) + 1/2 * ( H4N4L1 + H4N4E1 + H5N4L1 + H4N4F1E1 + H5N4E1 + H4N5E1 + H5N4F1L1 + H5N4F1E1 + H4N5F1E1 + H5N5E1 + H5N5F1E1 ) + 2/2 * ( H5N4L2 + H5N4E1L1 + H5N4E2 + H5N4F1L2 + H5N4F1E1L1 + H5N4F1E2 + H5N5E2 + H5N5F1E1L1 + H5N5F1E2 ) ) / ( H3N4 + H3N4F1 + H4N4 + H3N5 + H4N4F1 + H5N4 + H3N5F1 + H4N5 + H4N4L1 + H5N4F1 + H4N4E1 + H4N5F1 + H5N5 + H5N4L1 + H4N4F1E1 + H5N4E1 + H5N5F1 + H4N5E1 + H5N4F1L1 + H5N4F1E1 + H4N5F1E1 + H5N5E1 + H5N4L2 + H5N4E1L1 + H5N4E2 + H5N5F1E1 + H5N4F1L2 + H5N4F1E1L1 + H5N4F1E2 + H5N5E2 + H5N5F1E1L1 + H5N5F1E2 ) |
| **A3S** | Sialylation per antenna within triantennary glycans | A3S = ( 0/3 * (0) + 1/3 * ( H6N5E1 + H6N5F1E1 ) + 2/3 * ( H6N5L2 + H6N5E1L1 + H6N5E2 + H6N5F1L2 + H6N5F1E1L1 + H6N5F1E2 ) + 3/3 * ( H6N5E1L2 + H6N5E2L1 + H6N5E3 + H6N5F1E1L2 + H6N5F1E2L1 + H6N5F1E3 + H6N5F2E2L1 ) ) / ( H6N5E1 + H6N5F1E1 + H6N5L2 + H6N5E1L1 + H6N5E2 + H6N5F1L2 + H6N5F1E1L1 + H6N5F1E2 + H6N5E1L2 + H6N5E2L1 + H6N5E3 + H6N5F1E1L2 + H6N5F1E2L1 + H6N5F1E3 + H6N5F2E2L1 ) |
| **A4S** | Sialylation per antenna within tetra-antennary glycans | A4S = ( 0/4 * (0) + 1/4 * ( H4N6F1E1 ) + 2/4 * (0) + 3/4 * ( H7N6E1L2 + H7N6E2L1 + H7N6F1E1L2 + H7N6F1E2L1 ) + 4/4 * ( H7N6E1L3 + H7N6E2L2 + H7N6E3L1 + H7N6F1E1L3 + H7N6F1E2L2 ) ) / ( H4N6F1E1 + H7N6E1L2 + H7N6E2L1 + H7N6F1E1L2 + H7N6F1E2L1 + H7N6E1L3 + H7N6E2L2 + H7N6E3L1 + H7N6F1E1L3 + H7N6F1E2L2 ) |
| **A2F0S** | Sialylation per antenna within non-fucosylated diantennary glycans | A2F0S = ( 0/2 * ( H3N4 + H4N4 + H3N5 + H5N4 + H4N5 + H5N5 ) + 1/2 * ( H4N4L1 + H4N4E1 + H5N4L1 + H5N4E1 + H4N5E1 + H5N5E1 ) + 2/2 * ( H5N4L2 + H5N4E1L1 + H5N4E2 + H5N5E2 ) ) / ( H3N4 + H4N4 + H3N5 + H5N4 + H4N5 + H4N4L1 + H4N4E1 + H5N5 + H5N4L1 + H5N4E1 + H4N5E1 + H5N5E1 + H5N4L2 + H5N4E1L1 + H5N4E2 + H5N5E2 ) |
| **A3F0S** | Sialylation per antenna within non-fucosylated triantennary glycans | A3F0S = ( 0/3 * (0) + 1/3 * ( H6N5E1 ) + 2/3 * ( H6N5L2 + H6N5E1L1 + H6N5E2 ) + 3/3 * ( H6N5E1L2 + H6N5E2L1 + H6N5E3 ) ) / ( H6N5E1 + H6N5L2 + H6N5E1L1 + H6N5E2 + H6N5E1L2 + H6N5E2L1 + H6N5E3 ) |
| **A4F0S** | Sialylation per antenna within non-fucosylated tetra-antennary glycans | A4F0S = ( 0/4 * (0) + 1/4 * (0) + 2/4 * (0) + 3/4 * ( H7N6E1L2 + H7N6E2L1 ) + 4/4 * ( H7N6E1L3 + H7N6E2L2 + H7N6E3L1 ) ) / ( H4N6F1E1 + H7N6E1L2 + H7N6E2L1 + H7N6F1E1L2 + H7N6F1E2L1 + H7N6E1L3 + H7N6E2L2 + H7N6E3L1 + H7N6F1E1L3 + H7N6F1E2L2 ) |
| **A2FS** | Sialylation per antenna within fucosylated diantennary glycans | A2FS = ( 0/2 * ( H3N4F1 + H4N4F1 + H3N5F1 + H5N4F1 + H4N5F1 + H5N5F1 ) + 1/2 * ( H4N4F1E1 + H5N4F1L1 + H5N4F1E1 + H4N5F1E1 + H5N5F1E1 ) + 2/2 * ( H5N4F1L2 + H5N4F1E1L1 + H5N4F1E2 + H5N5F1E1L1 + H5N5F1E2 ) ) / ( H3N4F1 + H4N4F1 + H3N5F1 + H5N4F1 + H4N5F1 + H4N4F1E1 + H5N5F1 + H5N4F1L1 + H5N4F1E1 + H4N5F1E1 + H5N5F1E1 + H5N4F1L2 + H5N4F1E1L1 + H5N4F1E2 + H5N5F1E1L1 + H5N5F1E2 ) |
| **A3FS** | Sialylation per antenna within fucosylated triantennary glycans | A3FS = ( 0/3 * (0) + 1/3 * ( H6N5F1E1 ) + 2/3 * ( H6N5F1L2 + H6N5F1E1L1 + H6N5F1E2 ) + 3/3 * ( H6N5F1E1L2 + H6N5F1E2L1 + H6N5F1E3 + H6N5F2E2L1 ) ) / ( H6N5F1E1 + H6N5F1L2 + H6N5F1E1L1 + H6N5F1E2 + H6N5F1E1L2 + H6N5F1E2L1 + H6N5F1E3 + H6N5F2E2L1 ) |

| | | |
|---|---|---|
| **A4FS** | Sialylation per antenna within fucosylated tetra-antennary glycans | A4FS = ( 0/4 * (0) + 1/4 * ( H4N6F1E1 ) + 2/4 * (0) + 3/4 * ( H7N6F1E1L2 + H7N6F1E2L1 ) + 4/4 * ( H7N6F1E1L3 + H7N6F1E2L2 ) ) / ( H4N6F1E1 + H7N6E1L2 + H7N6E2L1 + H7N6F1E1L2 + H7N6F1E2L1 + H7N6E1L3 + H7N6E2L2 + H7N6E3L1 + H7N6F1E1L3 + H7N6F1E2L2 ) |
| **A2GS** | Sialylation per galactose within diantennary glycans | A2GS = ( ( 0/2 * ( H3N4 + H3N4F1 + H4N4 + H3N5 + H4N4F1 + H5N4 + H3N5F1 + H4N5 + H5N4F1 + H4N5F1 + H5N5 + H5N5F1 ) + 1/2 * ( H4N4L1 + H4N4E1 + H5N4L1 + H4N4F1E1 + H5N4E1 + H4N5E1 + H5N4F1L1 + H5N4F1E1 + H4N5F1E1 + H5N5E1 + H5N5F1E1 ) + 2/2 * ( H5N4L2 + H5N4E1L1 + H5N4E2 + H5N4F1L2 + H5N4F1E1L1 + H5N4F1E2 + H5N5E2 + H5N5F1E1L1 + H5N5F1E2 ) ) / ( H3N4 + H3N4F1 + H4N4 + H3N5 + H4N4F1 + H5N4 + H3N5F1 + H4N5 + H4N4L1 + H5N4F1 + H4N4E1 + H4N5F1 + H5N5 + H5N4L1 + H4N4F1E1 + H5N4E1 + H5N5F1 + H4N5E1 + H5N4F1L1 + H5N4F1E1 + H4N5F1E1 + H5N5E1 + H5N4L2 + H5N4E1L1 + H5N4E2 + H5N5F1E1 + H5N4F1L2 + H5N4F1E1L1 + H5N4F1E2 + H5N5E2 + H5N5F1E1L1 + H5N5F1E2 ) ) / ( ( 0/2 * ( H3N4 + H3N4F1 + H3N5 + H3N5F1 ) + 1/2 * ( H4N4 + H4N4F1 + H4N5 + H4N4L1 + H4N4E1 + H4N5F1 + H4N4F1E1 + H4N5E1 + H4N5F1E1 ) + 2/2 * ( H5N4 + H5N4F1 + H5N5 + H5N4L1 + H5N4E1 + H5N5F1 + H5N4F1L1 + H5N4F1E1 + H5N5E1 + H5N4L2 + H5N4E1L1 + H5N4E2 + H5N5F1E1 + H5N4F1L2 + H5N4F1E1L1 + H5N4F1E2 + H5N5E2 + H5N5F1E1L1 + H5N5F1E2 ) ) / ( H3N4 + H3N4F1 + H4N4 + H3N5 + H4N4F1 + H5N4 + H3N5F1 + H4N5 + H4N4L1 + H5N4F1 + H4N4E1 + H4N5F1 + H5N5 + H5N4L1 + H4N4F1E1 + H5N4E1 + H5N5F1 + H4N5E1 + H5N4F1L1 + H5N4F1E1 + H4N5F1E1 + H5N5E1 + H5N4L2 + H5N4E1L1 + H5N4E2 + H5N5F1E1 + H5N4F1L2 + H5N4F1E1L1 + H5N4F1E2 + H5N5E2 + H5N5F1E1L1 + H5N5F1E2 ) ) |
| **A3GS** | Sialylation per galactose within triantennary glycans | A3GS = ( ( 0/3 * (0) + 1/3 * ( H6N5E1 + H6N5F1E1 ) + 2/3 * ( H6N5L2 + H6N5E1L1 + H6N5E2 + H6N5F1L2 + H6N5F1E1L1 + H6N5F1E2 ) + 3/3 * ( H6N5E1L2 + H6N5E2L1 + H6N5E3 + H6N5F1E1L2 + H6N5F1E2L1 + H6N5F1E3 + H6N5F2E2L1 ) ) / ( H6N5E1 + H6N5F1E1 + H6N5L2 + H6N5E1L1 + H6N5E2 + H6N5F1L2 + H6N5F1E1L1 + H6N5F1E2 + H6N5E1L2 + H6N5E2L1 + H6N5E3 + H6N5F1E1L2 + H6N5F1E2L1 + H6N5F1E3 + H6N5F2E2L1 ) ) / ( ( 0/3 * (0) + 1/3 * (0) + 2/3 * (0) + 3/3 * ( H6N5E1 + H6N5F1E1 + H6N5L2 + H6N5E1L1 + H6N5E2 + H6N5F1L2 + H6N5F1E1L1 + H6N5F1E2 + H6N5E1L2 + H6N5E2L1 + H6N5E3 + H6N5F1E1L2 + H6N5F1E2L1 + H6N5F1E3 + H6N5F2E2L1 ) ) / ( H6N5E1 + H6N5F1E1 + H6N5L2 + H6N5E1L1 + H6N5E2 + H6N5F1L2 + H6N5F1E1L1 + H6N5F1E2 + H6N5E1L2 + H6N5E2L1 + H6N5E3 + H6N5F1E1L2 + H6N5F1E2L1 + H6N5F1E3 + H6N5F2E2L1 ) ) |
| **A4GS** | Sialylation per galactose within tetra-antennary glycans | A4GS = ( ( 0/4 * (0) + 1/4 * ( H4N6F1E1 ) + 2/4 * (0) + 3/4 * ( H7N6E1L2 + H7N6E2L1 + H7N6F1E1L2 + H7N6F1E2L1 ) + 4/4 * ( H7N6E1L3 + H7N6E2L2 + H7N6E3L1 + H7N6F1E1L3 + H7N6F1E2L2 ) ) / ( H4N6F1E1 + H7N6E1L2 + H7N6E2L1 + H7N6F1E1L2 + H7N6F1E2L1 + H7N6E1L3 + H7N6E2L2 + H7N6E3L1 + H7N6F1E1L3 + H7N6F1E2L2 ) ) / ( ( 0/4 * (0) + 1/4 * ( H4N6F1E1 ) + 2/4 * (0) + 3/4 * (0) + 4/4 * ( H7N6E1L2 + H7N6E2L1 + H7N6F1E1L2 + H7N6F1E2L1 + H7N6E1L3 + H7N6E2L2 + H7N6E3L1 + H7N6F1E1L3 + H7N6F1E2L2 ) ) / ( H4N6F1E1 + H7N6E1L2 + H7N6E2L1 + H7N6F1E1L2 + H7N6F1E2L1 + H7N6E1L3 + H7N6E2L2 + H7N6E3L1 + H7N6F1E1L3 + H7N6F1E2L2 ) ) |

| | | |
|---|---|---|
| **A2F0GS** | Sialylation per galactose within non-fucosylated diantennary glycans | A2F0GS = ( ( 0/2 * ( H3N4 + H4N4 + H3N5 + H5N4 + H4N5 + H5N5 ) + 1/2 * ( H4N4L1 + H4N4E1 + H5N4L1 + H5N4E1 + H4N5E1 + H5N5E1 ) + 2/2 * ( H5N4L2 + H5N4E1L1 + H5N4E2 + H5N5E2 ) ) / ( H3N4 + H4N4 + H3N5 + H5N4 + H4N5 + H4N4L1 + H4N4E1 + H5N5 + H5N4L1 + H5N4E1 + H4N5E1 + H5N5E1 + H5N4L2 + H5N4E1L1 + H5N4E2 + H5N5E2 ) ) / ( ( 0/2 * ( H3N4 + H3N5 ) + 1/2 * ( H4N4 + H4N5 + H4N4L1 + H4N4E1 + H4N5E1 ) + 2/2 * ( H5N4 + H5N5 + H5N4L1 + H5N4E1 + H5N5E1 + H5N4L2 + H5N4E1L1 + H5N4E2 + H5N5E2 ) ) / ( H3N4 + H4N4 + H3N5 + H5N4 + H4N5 + H4N4L1 + H4N4E1 + H5N5 + H5N4L1 + H5N4E1 + H4N5E1 + H5N5E1 + H5N4L2 + H5N4E1L1 + H5N4E2 + H5N5E2 ) ) |
| **A3F0GS** | Sialylation per galactose within non-fucosylated triantennary glycans | A3F0GS = ( ( 0/3 * (0) + 1/3 * ( H6N5E1 ) + 2/3 * ( H6N5L2 + H6N5E1L1 + H6N5E2 ) + 3/3 * ( H6N5E1L2 + H6N5E2L1 + H6N5E3 ) ) / ( H6N5E1 + H6N5L2 + H6N5E1L1 + H6N5E2 + H6N5E1L2 + H6N5E2L1 + H6N5E3 ) ) / ( ( 0/3 * (0) + 1/3 * (0) + 2/3 * (0) + 3/3 * ( H6N5E1 + H6N5L2 + H6N5E1L1 + H6N5E2 + H6N5E1L2 + H6N5E2L1 + H6N5E3 ) ) / ( H6N5E1 + H6N5L2 + H6N5E1L1 + H6N5E2 + H6N5E1L2 + H6N5E2L1 + H6N5E3 ) ) |
| **A4F0GS** | Sialylation per galactose within non-fucosylated tetra-antennary glycans | A4F0GS = ( ( 0/4 * (0) + 1/4 * (0) + 2/4 * (0) + 3/4 * ( H7N6E1L2 + H7N6E2L1 ) + 4/4 * ( H7N6E1L3 + H7N6E2L2 + H7N6E3L1 ) ) / ( H4N6F1E1 + H7N6E1L2 + H7N6E2L1 + H7N6F1E1L2 + H7N6F1E2L1 + H7N6E1L3 + H7N6E2L2 + H7N6E3L1 + H7N6F1E1L3 + H7N6F1E2L2 ) ) / ( ( 0/4 * (0) + 1/4 * (0) + 2/4 * (0) + 3/4 * (0) + 4/4 * ( H7N6E1L2 + H7N6E2L1 + H7N6E1L3 + H7N6E2L2 + H7N6E3L1 ) ) / ( H4N6F1E1 + H7N6E1L2 + H7N6E2L1 + H7N6F1E1L2 + H7N6F1E2L1 + H7N6E1L3 + H7N6E2L2 + H7N6E3L1 + H7N6F1E1L3 + H7N6F1E2L2 ) ) |
| **A2FGS** | Sialylation per galactose within fucosylated diantennary glycans | A2FGS = ( ( 0/2 * ( H3N4F1 + H4N4F1 + H3N5F1 + H5N4F1 + H4N5F1 + H5N5F1 ) + 1/2 * ( H4N4F1E1 + H5N4F1L1 + H5N4F1E1 + H4N5F1E1 + H5N5F1E1 ) + 2/2 * ( H5N4F1L2 + H5N4F1E1L1 + H5N4F1E2 + H5N5F1E1L1 + H5N5F1E2 ) ) / ( H3N4F1 + H4N4F1 + H3N5F1 + H5N4F1 + H4N5F1 + H4N4F1E1 + H5N5F1 + H5N4F1L1 + H5N4F1E1 + H4N5F1E1 + H5N5F1E1 + H5N4F1L2 + H5N4F1E1L1 + H5N4F1E2 + H5N5F1E1L1 + H5N5F1E2 ) ) / ( ( 0/2 * ( H3N4F1 + H3N5F1 ) + 1/2 * ( H4N4F1 + H4N5F1 + H4N4F1E1 + H4N5F1E1 ) + 2/2 * ( H5N4F1 + H5N5F1 + H5N4F1L1 + H5N4F1E1 + H5N5F1E1 + H5N4F1L2 + H5N4F1E1L1 + H5N4F1E2 + H5N5F1E1L1 + H5N5F1E2 ) ) / ( H3N4F1 + H4N4F1 + H3N5F1 + H5N4F1 + H4N5F1 + H4N4F1E1 + H5N5F1 + H5N4F1L1 + H5N4F1E1 + H4N5F1E1 + H5N5F1E1 + H5N4F1L2 + H5N4F1E1L1 + H5N4F1E2 + H5N5F1E1L1 + H5N5F1E2 ) ) |
| **A3FGS** | Sialylation per galactose within fucosylated triantennary glycans | A3FGS = ( ( 0/3 * (0) + 1/3 * ( H6N5F1E1 ) + 2/3 * ( H6N5F1L2 + H6N5F1E1L1 + H6N5F1E2 ) + 3/3 * ( H6N5F1E1L2 + H6N5F1E2L1 + H6N5F1E3 + H6N5F2E2L1 ) ) / ( H6N5F1E1 + H6N5F1L2 + H6N5F1E1L1 + H6N5F1E2 + H6N5F1E1L2 + H6N5F1E2L1 + H6N5F1E3 + H6N5F2E2L1 ) ) / ( ( 0/3 * (0) + 1/3 * (0) + 2/3 * (0) + 3/3 * ( H6N5F1E1 + H6N5F1L2 + H6N5F1E1L1 + H6N5F1E2 + H6N5F1E1L2 + H6N5F1E2L1 + H6N5F1E3 + H6N5F2E2L1 ) ) / ( H6N5F1E1 + H6N5F1L2 + H6N5F1E1L1 + H6N5F1E2 + H6N5F1E1L2 + H6N5F1E2L1 + H6N5F1E3 + H6N5F2E2L1 ) ) |
| **A4FGS** | Sialylation per galactose within fucosylated tetra-antennary glycans | A4FGS = ( ( 0/4 * (0) + 1/4 * ( H4N6F1E1 ) + 2/4 * (0) + 3/4 * ( H7N6F1E1L2 + H7N6F1E2L1 ) + 4/4 * ( H7N6F1E1L3 + H7N6F1E2L2 ) ) / ( H4N6F1E1 + H7N6E1L2 + H7N6E2L1 + H7N6F1E1L2 + H7N6F1E2L1 + H7N6E1L3 + H7N6E2L2 + H7N6E3L1 + H7N6F1E1L3 + H7N6F1E2L2 ) ) / ( ( 0/4 * (0) + 1/4 * ( H4N6F1E1 ) + 2/4 * (0) + 3/4 * (0) + 4/4 * ( H7N6F1E1L2 + H7N6F1E2L1 + H7N6F1E1L3 + H7N6F1E2L2 ) ) / ( H4N6F1E1 + H7N6E1L2 + H7N6E2L1 + H7N6F1E1L2 + H7N6F1E2L1 + H7N6E1L3 + H7N6E2L2 + H7N6E3L1 + H7N6F1E1L3 + H7N6F1E2L2 ) ) |

| | | |
|---|---|---|
| **α2,3-linked sialylation (L)** | | |
| **A2L** | α2,3-sialylation per antenna within diantennary glycans | A2L = ( 0/2 * ( H3N4 + H3N4F1 + H4N4 + H3N5 + H4N4F1 + H5N4 + H3N5F1 + H4N5 + H5N4F1 + H4N4E1 + H4N5F1 + H5N5 + H4N4F1E1 + H5N4E1 + H5N5F1 + H4N5E1 + H5N4F1E1 + H4N5F1E1 + H5N5E1 + H5N4E2 + H5N5F1E1 + H5N4F1E2 + H5N5E2 + H5N5F1E2 ) + 1/2 * ( H4N4L1 + H5N4L1 + H5N4F1L1 + H5N4E1L1 + H5N4F1E1L1 + H5N5F1E1L1 ) + 2/2 * ( H5N4L2 + H5N4F1L2 ) ) / ( H3N4 + H3N4F1 + H4N4 + H3N5 + H4N4F1 + H5N4 + H3N5F1 + H4N5 + H4N4L1 + H5N4F1 + H4N4E1 + H4N5F1 + H5N5 + H5N4L1 + H4N4F1E1 + H5N4E1 + H5N5F1 + H4N5E1 + H5N4F1L1 + H5N4F1E1 + H4N5F1E1 + H5N5E1 + H5N4L2 + H5N4E1L1 + H5N4E2 + H5N5F1E1 + H5N4F1L2 + H5N4F1E1L1 + H5N4F1E2 + H5N5E2 + H5N5F1E1L1 + H5N5F1E2 ) |
| **A3L** | α2,3-sialylation per antenna within triantennary glycans | A3L = ( 0/3 * ( H6N5E1 + H6N5F1E1 + H6N5E2 + H6N5F1E2 + H6N5E3 + H6N5F1E3 ) + 1/3 * ( H6N5E1L1 + H6N5F1E1L1 + H6N5E2L1 + H6N5F1E2L1 + H6N5F2E2L1 ) + 2/3 * ( H6N5L2 + H6N5F1L2 + H6N5E1L2 + H6N5F1E1L2 ) + 3/3 * (0) ) / ( H6N5E1 + H6N5F1E1 + H6N5L2 + H6N5E1L1 + H6N5E2 + H6N5F1L2 + H6N5F1E1L1 + H6N5F1E2 + H6N5E1L2 + H6N5E2L1 + H6N5E3 + H6N5F1E1L2 + H6N5F1E2L1 + H6N5F1E3 + H6N5F2E2L1 ) |
| **A4L** | α2,3-sialylation per antenna within tetra-antennary glycans | A4L = ( 0/4 * ( H4N6F1E1 ) + 1/4 * ( H7N6E2L1 + H7N6F1E2L1 + H7N6E3L1 ) + 2/4 * ( H7N6E1L2 + H7N6F1E1L2 + H7N6E2L2 + H7N6F1E2L2 ) + 3/4 * ( H7N6E1L3 + H7N6F1E1L3 ) + 4/4 * (0) ) / ( H4N6F1E1 + H7N6E1L2 + H7N6E2L1 + H7N6F1E1L2 + H7N6F1E2L1 + H7N6E1L3 + H7N6E2L2 + H7N6E3L1 + H7N6F1E1L3 + H7N6F1E2L2 ) |
| **A2F0L** | α2,3-sialylation per antenna within non-fucosylated diantennary glycans | A2F0L = ( 0/2 * ( H3N4 + H4N4 + H3N5 + H5N4 + H4N5 + H4N4E1 + H5N5 + H5N4E1 + H4N5E1 + H5N5E1 + H5N4E2 + H5N5E2 ) + 1/2 * ( H4N4L1 + H5N4L1 + H5N4E1L1 ) + 2/2 * ( H5N4L2 ) ) / ( H3N4 + H4N4 + H3N5 + H5N4 + H4N5 + H4N4L1 + H4N4E1 + H5N5 + H5N4L1 + H5N4E1 + H4N5E1 + H5N5E1 + H5N4L2 + H5N4E1L1 + H5N4E2 + H5N5E2 ) |
| **A3F0L** | α2,3-sialylation per antenna within non-fucosylated triantennary glycans | A3F0L = ( 0/3 * ( H6N5E1 + H6N5E2 + H6N5E3 ) + 1/3 * ( H6N5E1L1 + H6N5E2L1 ) + 2/3 * ( H6N5L2 + H6N5E1L2 ) + 3/3 * (0) ) / ( H6N5E1 + H6N5L2 + H6N5E1L1 + H6N5E2 + H6N5E1L2 + H6N5E2L1 + H6N5E3 ) |
| **A4F0L** | α2,3-sialylation per antenna within non-fucosylated tetra-antennary glycans | A4F0L = ( 0/4 * (0) + 1/4 * ( H7N6E2L1 + H7N6E3L1 ) + 2/4 * ( H7N6E1L2 + H7N6E2L2 ) + 3/4 * ( H7N6E1L3 ) + 4/4 * (0) ) / ( H4N6F1E1 + H7N6E1L2 + H7N6E2L1 + H7N6F1E1L2 + H7N6F1E2L1 + H7N6E1L3 + H7N6E2L2 + H7N6E3L1 + H7N6F1E1L3 + H7N6F1E2L2 ) |
| **A2FL** | α2,3-sialylation per antenna within fucosylated diantennary glycans | A2FL = ( 0/2 * ( H3N4F1 + H4N4F1 + H3N5F1 + H5N4F1 + H4N5F1 + H4N4F1E1 + H5N5F1 + H5N4F1E1 + H4N5F1E1 + H5N5F1E1 + H5N4F1E2 + H5N5F1E2 ) + 1/2 * ( H5N4F1L1 + H5N4F1E1L1 + H5N5F1E1L1 ) + 2/2 * ( H5N4F1L2 ) ) / ( H3N4F1 + H4N4F1 + H3N5F1 + H5N4F1 + H4N5F1 + H4N4F1E1 + H5N5F1 + H5N4F1L1 + H5N4F1E1 + H4N5F1E1 + H5N5F1E1 + H5N4F1L2 + H5N4F1E1L1 + H5N4F1E2 + H5N5F1E1L1 + H5N5F1E2 ) |

| | | |
|---|---|---|
| **A3FL** | α2,3-sialylation per antenna within fucosylated triantennary glycans | A3FL = ( 0/3 * ( H6N5F1E1 + H6N5F1E2 + H6N5F1E3 ) + 1/3 * ( H6N5F1E1L1 + H6N5F1E2L1 + H6N5F2E2L1 ) + 2/3 * ( H6N5F1L2 + H6N5F1E1L2 ) + 3/3 * (0) ) / ( H6N5F1E1 + H6N5F1L2 + H6N5F1E1L1 + H6N5F1E2 + H6N5F1E1L2 + H6N5F1E2L1 + H6N5F1E3 + H6N5F2E2L1 ) |
| **A4FL** | α2,3-sialylation per antenna within fucosylated tetra-antennary glycans | A4FL = ( 0/4 * ( H4N6F1E1 ) + 1/4 * ( H7N6F1E2L1 ) + 2/4 * ( H7N6F1E1L2 + H7N6F1E2L2 ) + 3/4 * ( H7N6F1E1L3 ) + 4/4 * (0) ) / ( H4N6F1E1 + H7N6E1L2 + H7N6E2L1 + H7N6F1E1L2 + H7N6F1E2L1 + H7N6E1L3 + H7N6E2L2 + H7N6E3L1 + H7N6F1E1L3 + H7N6F1E2L2 ) |
| **A2GL** | α2,3-sialylation per galactose within diantennary glycans | A2GL = ( ( 0/2 * ( H3N4 + H3N4F1 + H4N4 + H3N5 + H4N4F1 + H5N4 + H3N5F1 + H4N5 + H5N4F1 + H4N4E1 + H4N5F1 + H5N5 + H4N4F1E1 + H5N4E1 + H5N5F1 + H4N5E1 + H5N4F1E1 + H4N5F1E1 + H5N5E1 + H5N4E2 + H5N5F1E1 + H5N4F1E2 + H5N5E2 + H5N5F1E2 ) + 1/2 * ( H4N4L1 + H5N4L1 + H5N4F1L1 + H5N4E1L1 + H5N4F1E1L1 + H5N5F1E1L1 ) + 2/2 * ( H5N4L2 + H5N4F1L2 ) ) / ( H3N4 + H3N4F1 + H4N4 + H3N5 + H4N4F1 + H5N4 + H3N5F1 + H4N5 + H4N4L1 + H5N4F1 + H4N4E1 + H4N5F1 + H5N5 + H5N4L1 + H4N4F1E1 + H5N4E1 + H5N5F1 + H4N5E1 + H5N4F1L1 + H5N4F1E1 + H4N5F1E1 + H5N5E1 + H5N4L2 + H5N4E1L1 + H5N4E2 + H5N5F1E1 + H5N4F1L2 + H5N4F1E1L1 + H5N4F1E2 + H5N5E2 + H5N5F1E1L1 + H5N5F1E2 ) ) / ( ( 0/2 * ( H3N4 + H3N4F1 + H3N5 + H3N5F1 ) + 1/2 * ( H4N4 + H4N4F1 + H4N5 + H4N4L1 + H4N4E1 + H4N5F1 + H4N4F1E1 + H4N5E1 + H4N5F1E1 ) + 2/2 * ( H5N4 + H5N4F1 + H5N5 + H5N4L1 + H5N4E1 + H5N5F1 + H5N4F1L1 + H5N4F1E1 + H5N5E1 + H5N4L2 + H5N4E1L1 + H5N4E2 + H5N5F1E1 + H5N4F1L2 + H5N4F1E1L1 + H5N4F1E2 + H5N5E2 + H5N5F1E1L1 + H5N5F1E2 ) ) / ( H3N4 + H3N4F1 + H4N4 + H3N5 + H4N4F1 + H5N4 + H3N5F1 + H4N5 + H4N4L1 + H5N4F1 + H4N4E1 + H4N5F1 + H5N5 + H5N4L1 + H4N4F1E1 + H5N4E1 + H5N5F1 + H4N5E1 + H5N4F1L1 + H5N4F1E1 + H4N5F1E1 + H5N5E1 + H5N4L2 + H5N4E1L1 + H5N4E2 + H5N5F1E1 + H5N4F1L2 + H5N4F1E1L1 + H5N4F1E2 + H5N5E2 + H5N5F1E1L1 + H5N5F1E2 ) ) |
| **A3GL** | α2,3-sialylation per galactose within triantennary glycans | A3GL = ( ( 0/3 * ( H6N5E1 + H6N5F1E1 + H6N5E2 + H6N5F1E2 + H6N5E3 + H6N5F1E3 ) + 1/3 * ( H6N5E1L1 + H6N5F1E1L1 + H6N5E2L1 + H6N5F1E2L1 + H6N5F2E2L1 ) + 2/3 * ( H6N5L2 + H6N5F1L2 + H6N5E1L2 + H6N5F1E1L2 ) + 3/3 * (0) ) / ( H6N5E1 + H6N5F1E1 + H6N5L2 + H6N5E1L1 + H6N5E2 + H6N5F1L2 + H6N5F1E1L1 + H6N5F1E2 + H6N5E1L2 + H6N5E2L1 + H6N5E3 + H6N5F1E1L2 + H6N5F1E2L1 + H6N5F1E3 + H6N5F2E2L1 ) ) / ( ( 0/3 * (0) + 1/3 * (0) + 2/3 * (0) + 3/3 * ( H6N5E1 + H6N5F1E1 + H6N5L2 + H6N5E1L1 + H6N5E2 + H6N5F1L2 + H6N5F1E1L1 + H6N5F1E2 + H6N5E1L2 + H6N5E2L1 + H6N5E3 + H6N5F1E1L2 + H6N5F1E2L1 + H6N5F1E3 + H6N5F2E2L1 ) ) / ( H6N5E1 + H6N5F1E1 + H6N5L2 + H6N5E1L1 + H6N5E2 + H6N5F1L2 + H6N5F1E1L1 + H6N5F1E2 + H6N5E1L2 + H6N5E2L1 + H6N5E3 + H6N5F1E1L2 + H6N5F1E2L1 + H6N5F1E3 + H6N5F2E2L1 ) ) |
| **A4GL** | α2,3-sialylation per galactose within tetra-antennary glycans | A4GL = ( ( 0/4 * ( H4N6F1E1 ) + 1/4 * ( H7N6E2L1 + H7N6F1E2L1 + H7N6E3L1 ) + 2/4 * ( H7N6E1L2 + H7N6F1E1L2 + H7N6E2L2 + H7N6F1E2L2 ) + 3/4 * ( H7N6E1L3 + H7N6F1E1L3 ) + 4/4 * (0) ) / ( H4N6F1E1 + H7N6E1L2 + H7N6E2L1 + H7N6F1E1L2 + H7N6F1E2L1 + H7N6E1L3 + H7N6E2L2 + H7N6E3L1 + H7N6F1E1L3 + H7N6F1E2L2 ) ) / ( ( 0/4 * (0) + 1/4 * ( H4N6F1E1 ) + 2/4 * (0) + 3/4 * (0) + 4/4 * ( H7N6E1L2 + H7N6E2L1 + H7N6F1E1L2 + H7N6F1E2L1 |

| | | |
|---|---|---|
| | | + H7N6E1L3 + H7N6E2L2 + H7N6E3L1 + H7N6F1E1L3 + H7N6F1E2L2 ) ) / ( H4N6F1E1 + H7N6E1L2 + H7N6E2L1 + H7N6F1E1L2 + H7N6F1E2L1 + H7N6E1L3 + H7N6E2L2 + H7N6E3L1 + H7N6F1E1L3 + H7N6F1E2L2 ) ) |
| **A2F0GL** | α2,3-sialylation per galactose within non-fucosylated diantennary glycans | A2F0GL = ( ( 0/2 * ( H3N4 + H4N4 + H3N5 + H5N4 + H4N5 + H4N4E1 + H5N5 + H5N4E1 + H4N5E1 + H5N5E1 + H5N4E2 + H5N5E2 ) + 1/2 * ( H4N4L1 + H5N4L1 + H5N4E1L1 ) + 2/2 * ( H5N4L2 ) ) / ( H3N4 + H4N4 + H3N5 + H5N4 + H4N5 + H4N4L1 + H4N4E1 + H5N5 + H5N4L1 + H5N4E1 + H4N5E1 + H5N5E1 + H5N4L2 + H5N4E1L1 + H5N4E2 + H5N5E2 ) ) / ( ( 0/2 * ( H3N4 + H3N5 ) + 1/2 * ( H4N4 + H4N5 + H4N4L1 + H4N4E1 + H4N5E1 ) + 2/2 * ( H5N4 + H5N5 + H5N4L1 + H5N4E1 + H5N5E1 + H5N4L2 + H5N4E1L1 + H5N4E2 + H5N5E2 ) ) / ( H3N4 + H4N4 + H3N5 + H5N4 + H4N5 + H4N4L1 + H4N4E1 + H5N5 + H5N4L1 + H5N4E1 + H4N5E1 + H5N5E1 + H5N4L2 + H5N4E1L1 + H5N4E2 + H5N5E2 ) ) |
| **A3F0GL** | α2,3-sialylation per galactose within non-fucosylated triantennary glycans | A3F0GL = ( ( 0/3 * ( H6N5E1 + H6N5E2 + H6N5E3 ) + 1/3 * ( H6N5E1L1 + H6N5E2L1 ) + 2/3 * ( H6N5L2 + H6N5E1L2 ) + 3/3 * (0) ) / ( H6N5E1 + H6N5L2 + H6N5E1L1 + H6N5E2 + H6N5E1L2 + H6N5E2L1 + H6N5E3 ) ) / ( ( 0/3 * (0) + 1/3 * (0) + 2/3 * (0) + 3/3 * ( H6N5E1 + H6N5L2 + H6N5E1L1 + H6N5E2 + H6N5E1L2 + H6N5E2L1 + H6N5E3 ) ) / ( H6N5E1 + H6N5L2 + H6N5E1L1 + H6N5E2 + H6N5E1L2 + H6N5E2L1 + H6N5E3 ) ) |
| **A4F0GL** | α2,3-sialylation per galactose within non-fucosylated tetra-antennary glycans | A4F0GL = ( ( 0/4 * (0) + 1/4 * ( H7N6E2L1 + H7N6E3L1 ) + 2/4 * ( H7N6E1L2 + H7N6E2L2 ) + 3/4 * ( H7N6E1L3 ) + 4/4 * (0) ) / ( H4N6F1E1 + H7N6E1L2 + H7N6E2L1 + H7N6F1E1L2 + H7N6F1E2L1 + H7N6E1L3 + H7N6E2L2 + H7N6E3L1 + H7N6F1E1L3 + H7N6F1E2L2 ) ) / ( ( 0/4 * (0) + 1/4 * (0) + 2/4 * (0) + 3/4 * (0) + 4/4 * ( H7N6E1L2 + H7N6E2L1 + H7N6E1L3 + H7N6E2L2 + H7N6E3L1 ) ) / ( H4N6F1E1 + H7N6E1L2 + H7N6E2L1 + H7N6F1E1L2 + H7N6F1E2L1 + H7N6E1L3 + H7N6E2L2 + H7N6E3L1 + H7N6F1E1L3 + H7N6F1E2L2 ) ) |
| **A2FGL** | α2,3-sialylation per galactose within fucosylated diantennary glycans | A2FGL = ( ( 0/2 * ( H3N4F1 + H4N4F1 + H3N5F1 + H5N4F1 + H4N5F1 + H4N4F1E1 + H5N5F1 + H5N4F1E1 + H4N5F1E1 + H5N5F1E1 + H5N4F1E2 + H5N5F1E2 ) + 1/2 * ( H5N4F1L1 + H5N4F1E1L1 + H5N5F1E1L1 ) + 2/2 * ( H5N4F1L2 ) ) / ( H3N4F1 + H4N4F1 + H3N5F1 + H5N4F1 + H4N5F1 + H4N4F1E1 + H5N5F1 + H5N4F1L1 + H5N4F1E1 + H4N5F1E1 + H5N5F1E1 + H5N4F1L2 + H5N4F1E1L1 + H5N4F1E2 + H5N5F1E1L1 + H5N5F1E2 ) ) / ( ( 0/2 * ( H3N4F1 + H3N5F1 ) + 1/2 * ( H4N4F1 + H4N5F1 + H4N4F1E1 + H4N5F1E1 ) + 2/2 * ( H5N4F1 + H5N5F1 + H5N4F1L1 + H5N4F1E1 + H5N5F1E1 + H5N4F1L2 + H5N4F1E1L1 + H5N4F1E2 + H5N5F1E1L1 + H5N5F1E2 ) ) / ( H3N4F1 + H4N4F1 + H3N5F1 + H5N4F1 + H4N5F1 + H4N4F1E1 + H5N5F1 + H5N4F1L1 + H5N4F1E1 + H4N5F1E1 + H5N5F1E1 + H5N4F1L2 + H5N4F1E1L1 + H5N4F1E2 + H5N5F1E1L1 + H5N5F1E2 ) ) |
| **A3FGL** | α2,3-sialylation per galactose within fucosylated triantennary glycans | A3FGL = ( ( 0/3 * ( H6N5F1E1 + H6N5F1E2 + H6N5F1E3 ) + 1/3 * ( H6N5F1E1L1 + H6N5F1E2L1 + H6N5F2E2L1 ) + 2/3 * ( H6N5F1L2 + H6N5F1E1L2 ) + 3/3 * (0) ) / ( H6N5F1E1 + H6N5F1L2 + H6N5F1E1L1 + H6N5F1E2 + H6N5F1E1L2 + H6N5F1E2L1 + H6N5F1E3 + H6N5F2E2L1 ) ) / ( ( 0/3 * (0) + 1/3 * (0) + 2/3 * (0) + 3/3 * ( H6N5F1E1 + H6N5F1L2 + H6N5F1E1L1 + H6N5F1E2 + H6N5F1E1L2 + H6N5F1E2L1 + H6N5F1E3 + H6N5F2E2L1 ) ) / ( H6N5F1E1 + H6N5F1L2 + H6N5F1E1L1 + H6N5F1E2 + H6N5F1E1L2 + H6N5F1E2L1 + H6N5F1E3 + H6N5F2E2L1 ) ) |

| | | |
|---|---|---|
| **A4FGL** | α2,3-sialylation per galactose within fucosylated tetra-antennary glycans | A4FGL = ( ( 0/4 * ( H4N6F1E1 ) + 1/4 * ( H7N6F1E2L1 ) + 2/4 * ( H7N6F1E1L2 + H7N6F1E2L2 ) + 3/4 * ( H7N6F1E1L3 ) + 4/4 * (0) ) / ( H4N6F1E1 + H7N6E1L2 + H7N6E2L1 + H7N6F1E1L2 + H7N6F1E2L1 + H7N6E1L3 + H7N6E2L2 + H7N6E3L1 + H7N6F1E1L3 + H7N6F1E2L2 ) ) / ( ( 0/4 * (0) + 1/4 * ( H4N6F1E1 ) + 2/4 * (0) + 3/4 * (0) + 4/4 * ( H7N6F1E1L2 + H7N6F1E2L1 + H7N6F1E1L3 + H7N6F1E2L2 ) ) / ( H4N6F1E1 + H7N6E1L2 + H7N6E2L1 + H7N6F1E1L2 + H7N6F1E2L1 + H7N6E1L3 + H7N6E2L2 + H7N6E3L1 + H7N6F1E1L3 + H7N6F1E2L2 ) ) |
| **α2,6-linked sialylation (E)** | | |
| **A2E** | α2,6-sialylation per antenna within diantennary glycans | A2E = ( 0/2 * ( H3N4 + H3N4F1 + H4N4 + H3N5 + H4N4F1 + H5N4 + H3N5F1 + H4N5 + H4N4L1 + H5N4F1 + H4N5F1 + H5N5 + H5N4L1 + H5N5F1 + H5N4F1L1 + H5N4L2 + H5N4F1L2 ) + 1/2 * ( H4N4E1 + H4N4F1E1 + H5N4E1 + H4N5E1 + H5N4F1E1 + H4N5F1E1 + H5N5E1 + H5N4E1L1 + H5N5F1E1 + H5N4F1E1L1 + H5N5F1E1L1 ) + 2/2 * ( H5N4E2 + H5N4F1E2 + H5N5E2 + H5N5F1E2 ) ) / ( H3N4 + H3N4F1 + H4N4 + H3N5 + H4N4F1 + H5N4 + H3N5F1 + H4N5 + H4N4L1 + H5N4F1 + H4N4E1 + H4N5F1 + H5N5 + H5N4L1 + H4N4F1E1 + H5N4E1 + H5N5F1 + H4N5E1 + H5N4F1L1 + H5N4F1E1 + H4N5F1E1 + H5N5E1 + H5N4L2 + H5N4E1L1 + H5N4E2 + H5N5F1E1 + H5N4F1L2 + H5N4F1E1L1 + H5N4F1E2 + H5N5E2 + H5N5F1E1L1 + H5N5F1E2 ) |
| **A3E** | α2,6-sialylation per antenna within triantennary glycans | A3E = ( 0/3 * ( H6N5L2 + H6N5F1L2 ) + 1/3 * ( H6N5E1 + H6N5F1E1 + H6N5E1L1 + H6N5F1E1L1 + H6N5E1L2 + H6N5F1E1L2 ) + 2/3 * ( H6N5E2 + H6N5F1E2 + H6N5E2L1 + H6N5F1E2L1 + H6N5F2E2L1 ) + 3/3 * ( H6N5E3 + H6N5F1E3 ) ) / ( H6N5E1 + H6N5F1E1 + H6N5L2 + H6N5E1L1 + H6N5E2 + H6N5F1L2 + H6N5F1E1L1 + H6N5F1E2 + H6N5E1L2 + H6N5E2L1 + H6N5E3 + H6N5F1E1L2 + H6N5F1E2L1 + H6N5F1E3 + H6N5F2E2L1 ) |
| **A4E** | α2,6-sialylation per antenna within tetra-antennary glycans | A4E = ( 0/4 * (0) + 1/4 * ( H4N6F1E1 + H7N6E1L2 + H7N6F1E1L2 + H7N6E1L3 + H7N6F1E1L3 ) + 2/4 * ( H7N6E2L1 + H7N6F1E2L1 + H7N6E2L2 + H7N6F1E2L2 ) + 3/4 * ( H7N6E3L1 ) + 4/4 * (0) ) / ( H4N6F1E1 + H7N6E1L2 + H7N6E2L1 + H7N6F1E1L2 + H7N6F1E2L1 + H7N6E1L3 + H7N6E2L2 + H7N6E3L1 + H7N6F1E1L3 + H7N6F1E2L2 ) |
| **A2F0E** | α2,6-sialylation per antenna within non-fucosylated diantennary glycans | A2F0E = ( 0/2 * ( H3N4 + H4N4 + H3N5 + H5N4 + H4N5 + H4N4L1 + H5N5 + H5N4L1 + H5N4L2 ) + 1/2 * ( H4N4E1 + H5N4E1 + H4N5E1 + H5N5E1 + H5N4E1L1 ) + 2/2 * ( H5N4E2 + H5N5E2 ) ) / ( H3N4 + H4N4 + H3N5 + H5N4 + H4N5 + H4N4L1 + H4N4E1 + H5N5 + H5N4L1 + H5N4E1 + H4N5E1 + H5N5E1 + H5N4L2 + H5N4E1L1 + H5N4E2 + H5N5E2 ) |
| **A3F0E** | α2,6-sialylation per antenna within non-fucosylated triantennary glycans | A3F0E = ( 0/3 * ( H6N5L2 ) + 1/3 * ( H6N5E1 + H6N5E1L1 + H6N5E1L2 ) + 2/3 * ( H6N5E2 + H6N5E2L1 ) + 3/3 * ( H6N5E3 ) ) / ( H6N5E1 + H6N5L2 + H6N5E1L1 + H6N5E2 + H6N5E1L2 + H6N5E2L1 + H6N5E3 ) |
| **A4F0E** | α2,6-sialylation per antenna within non-fucosylated tetra-antennary glycans | A4F0E = ( 0/4 * (0) + 1/4 * ( H7N6E1L2 + H7N6E1L3 ) + 2/4 * ( H7N6E2L1 + H7N6E2L2 ) + 3/4 * ( H7N6E3L1 ) + 4/4 * (0) ) / ( H4N6F1E1 + H7N6E1L2 + H7N6E2L1 + H7N6F1E1L2 + H7N6F1E2L1 + H7N6E1L3 + H7N6E2L2 + H7N6E3L1 + H7N6F1E1L3 + H7N6F1E2L2 ) |

| | | |
|---|---|---|
| **A2FE** | α2,6-sialylation per antenna within fucosylated diantennary glycans | A2FE = ( 0/2 * ( H3N4F1 + H4N4F1 + H3N5F1 + H5N4F1 + H4N5F1 + H5N5F1 + H5N4F1L1 + H5N4F1L2 ) + 1/2 * ( H4N4F1E1 + H5N4F1E1 + H4N5F1E1 + H5N5F1E1 + H5N4F1E1L1 + H5N5F1E1L1 ) + 2/2 * ( H5N4F1E2 + H5N5F1E2 ) ) / ( H3N4F1 + H4N4F1 + H3N5F1 + H5N4F1 + H4N5F1 + H4N4F1E1 + H5N5F1 + H5N4F1L1 + H5N4F1E1 + H4N5F1E1 + H5N5F1E1 + H5N4F1L2 + H5N4F1E1L1 + H5N4F1E2 + H5N5F1E1L1 + H5N5F1E2 ) |
| **A3FE** | α2,6-sialylation per antenna within fucosylated triantennary glycans | A3FE = ( 0/3 * ( H6N5F1L2 ) + 1/3 * ( H6N5F1E1 + H6N5F1E1L1 + H6N5F1E1L2 ) + 2/3 * ( H6N5F1E2 + H6N5F1E2L1 + H6N5F2E2L1 ) + 3/3 * ( H6N5F1E3 ) ) / ( H6N5F1E1 + H6N5F1L2 + H6N5F1E1L1 + H6N5F1E2 + H6N5F1E1L2 + H6N5F1E2L1 + H6N5F1E3 + H6N5F2E2L1 ) |
| **A4FE** | α2,6-sialylation per antenna within fucosylated tetra-antennary glycans | A4FE = ( 0/4 * (0) + 1/4 * ( H4N6F1E1 + H7N6F1E1L2 + H7N6F1E1L3 ) + 2/4 * ( H7N6F1E2L1 + H7N6F1E2L2 ) + 3/4 * (0) + 4/4 * (0) ) / ( H4N6F1E1 + H7N6E1L2 + H7N6E2L1 + H7N6F1E1L2 + H7N6F1E2L1 + H7N6E1L3 + H7N6E2L2 + H7N6E3L1 + H7N6F1E1L3 + H7N6F1E2L2 ) |
| **A2GE** | α2,6-sialylation per galactose within diantennary glycans | A2GE = ( ( 0/2 * ( H3N4 + H3N4F1 + H4N4 + H3N5 + H4N4F1 + H5N4 + H3N5F1 + H4N5 + H4N4L1 + H5N4F1 + H4N5F1 + H5N5 + H5N4L1 + H5N5F1 + H5N4F1L1 + H5N4L2 + H5N4F1L2 ) + 1/2 * ( H4N4E1 + H4N4F1E1 + H5N4E1 + H4N5E1 + H5N4F1E1 + H4N5F1E1 + H5N5E1 + H5N4E1L1 + H5N5F1E1 + H5N4F1E1L1 + H5N5F1E1L1 ) + 2/2 * ( H5N4E2 + H5N4F1E2 + H5N5E2 + H5N5F1E2 ) ) / ( H3N4 + H3N4F1 + H4N4 + H3N5 + H4N4F1 + H5N4 + H3N5F1 + H4N5 + H4N4L1 + H5N4F1 + H4N4E1 + H4N5F1 + H5N5 + H5N4L1 + H4N4F1E1 + H5N4E1 + H5N5F1 + H4N5E1 + H5N4F1L1 + H5N4F1E1 + H4N5F1E1 + H5N5E1 + H5N4L2 + H5N4E1L1 + H5N4E2 + H5N5F1E1 + H5N4F1L2 + H5N4F1E1L1 + H5N4F1E2 + H5N5E2 + H5N5F1E1L1 + H5N5F1E2 ) ) / ( ( 0/2 * ( H3N4 + H3N4F1 + H3N5 + H3N5F1 ) + 1/2 * ( H4N4 + H4N4F1 + H4N5 + H4N4L1 + H4N4E1 + H4N5F1 + H4N4F1E1 + H4N5E1 + H4N5F1E1 ) + 2/2 * ( H5N4 + H5N4F1 + H5N5 + H5N4L1 + H5N4E1 + H5N5F1 + H5N4F1L1 + H5N4F1E1 + H5N5E1 + H5N4L2 + H5N4E1L1 + H5N4E2 + H5N5F1E1 + H5N4F1L2 + H5N4F1E1L1 + H5N4F1E2 + H5N5E2 + H5N5F1E1L1 + H5N5F1E2 ) ) / ( H3N4 + H3N4F1 + H4N4 + H3N5 + H4N4F1 + H5N4 + H3N5F1 + H4N5 + H4N4L1 + H5N4F1 + H4N4E1 + H4N5F1 + H5N5 + H5N4L1 + H4N4F1E1 + H5N4E1 + H5N5F1 + H4N5E1 + H5N4F1L1 + H5N4F1E1 + H4N5F1E1 + H5N5E1 + H5N4L2 + H5N4E1L1 + H5N4E2 + H5N5F1E1 + H5N4F1L2 + H5N4F1E1L1 + H5N4F1E2 + H5N5E2 + H5N5F1E1L1 + H5N5F1E2 ) ) |
| **A3GE** | α2,6-sialylation per galactose within triantennary glycans | A3GE = ( ( 0/3 * ( H6N5L2 + H6N5F1L2 ) + 1/3 * ( H6N5E1 + H6N5F1E1 + H6N5E1L1 + H6N5F1E1L1 + H6N5E1L2 + H6N5F1E1L2 ) + 2/3 * ( H6N5E2 + H6N5F1E2 + H6N5E2L1 + H6N5F1E2L1 + H6N5F2E2L1 ) + 3/3 * ( H6N5E3 + H6N5F1E3 ) ) / ( H6N5E1 + H6N5F1E1 + H6N5L2 + H6N5E1L1 + H6N5E2 + H6N5F1L2 + H6N5F1E1L1 + H6N5F1E2 + H6N5E1L2 + H6N5E2L1 + H6N5E3 + H6N5F1E1L2 + H6N5F1E2L1 + H6N5F1E3 + H6N5F2E2L1 ) ) / ( ( 0/3 * (0) + 1/3 * (0) + 2/3 * (0) + 3/3 * ( H6N5E1 + H6N5F1E1 + H6N5L2 + H6N5E1L1 + H6N5E2 + H6N5F1L2 + H6N5F1E1L1 + H6N5F1E2 + H6N5E1L2 + H6N5E2L1 + H6N5E3 + H6N5F1E1L2 + H6N5F1E2L1 + H6N5F1E3 + H6N5F2E2L1 ) ) / ( H6N5E1 + H6N5F1E1 + H6N5L2 + H6N5E1L1 + H6N5E2 + H6N5F1L2 + H6N5F1E1L1 + H6N5F1E2 + H6N5E1L2 + H6N5E2L1 + H6N5E3 + H6N5F1E1L2 + H6N5F1E2L1 + H6N5F1E3 + H6N5F2E2L1 ) ) |

| | | |
|---|---|---|
| **A4GE** | α2,6-sialylation per galactose within tetra-antennary glycans | A4GE = ( ( 0/4 * (0) + 1/4 * ( H4N6F1E1 + H7N6E1L2 + H7N6F1E1L2 + H7N6E1L3 + H7N6F1E1L3 ) + 2/4 * ( H7N6E2L1 + H7N6F1E2L1 + H7N6E2L2 + H7N6F1E2L2 ) + 3/4 * ( H7N6E3L1 ) + 4/4 * (0) ) / ( H4N6F1E1 + H7N6E1L2 + H7N6E2L1 + H7N6F1E1L2 + H7N6F1E2L1 + H7N6E1L3 + H7N6E2L2 + H7N6E3L1 + H7N6F1E1L3 + H7N6F1E2L2 ) ) / ( ( 0/4 * (0) + 1/4 * ( H4N6F1E1 ) + 2/4 * (0) + 3/4 * (0) + 4/4 * ( H7N6E1L2 + H7N6E2L1 + H7N6F1E1L2 + H7N6F1E2L1 + H7N6E1L3 + H7N6E2L2 + H7N6E3L1 + H7N6F1E1L3 + H7N6F1E2L2 ) ) / ( H4N6F1E1 + H7N6E1L2 + H7N6E2L1 + H7N6F1E1L2 + H7N6F1E2L1 + H7N6E1L3 + H7N6E2L2 + H7N6E3L1 + H7N6F1E1L3 + H7N6F1E2L2 ) ) |
| **A2F0GE** | α2,6-sialylation per galactose within non-fucosylated diantennary glycans | A2F0GE = ( ( 0/2 * ( H3N4 + H4N4 + H3N5 + H5N4 + H4N5 + H4N4L1 + H5N5 + H5N4L1 + H5N4L2 ) + 1/2 * ( H4N4E1 + H5N4E1 + H4N5E1 + H5N5E1 + H5N4E1L1 ) + 2/2 * ( H5N4E2 + H5N5E2 ) ) / ( H3N4 + H4N4 + H3N5 + H5N4 + H4N5 + H4N4L1 + H4N4E1 + H5N5 + H5N4L1 + H5N4E1 + H4N5E1 + H5N5E1 + H5N4L2 + H5N4E1L1 + H5N4E2 + H5N5E2 ) ) / ( ( 0/2 * ( H3N4 + H3N5 ) + 1/2 * ( H4N4 + H4N5 + H4N4L1 + H4N4E1 + H4N5E1 ) + 2/2 * ( H5N4 + H5N5 + H5N4L1 + H5N4E1 + H5N5E1 + H5N4L2 + H5N4E1L1 + H5N4E2 + H5N5E2 ) ) / ( H3N4 + H4N4 + H3N5 + H5N4 + H4N5 + H4N4L1 + H4N4E1 + H5N5 + H5N4L1 + H5N4E1 + H4N5E1 + H5N5E1 + H5N4L2 + H5N4E1L1 + H5N4E2 + H5N5E2 ) ) |
| **A3F0GE** | α2,6-sialylation per galactose within non-fucosylated triantennary glycans | A3F0GE = ( ( 0/3 * ( H6N5L2 ) + 1/3 * ( H6N5E1 + H6N5E1L1 + H6N5E1L2 ) + 2/3 * ( H6N5E2 + H6N5E2L1 ) + 3/3 * ( H6N5E3 ) ) / ( H6N5E1 + H6N5L2 + H6N5E1L1 + H6N5E2 + H6N5E1L2 + H6N5E2L1 + H6N5E3 ) ) / ( ( 0/3 * (0) + 1/3 * (0) + 2/3 * (0) + 3/3 * ( H6N5E1 + H6N5L2 + H6N5E1L1 + H6N5E2 + H6N5E1L2 + H6N5E2L1 + H6N5E3 ) ) / ( H6N5E1 + H6N5L2 + H6N5E1L1 + H6N5E2 + H6N5E1L2 + H6N5E2L1 + H6N5E3 ) ) |
| **A4F0GE** | α2,6-sialylation per galactose within non-fucosylated tetra-antennary glycans | A4F0GE = ( ( 0/4 * (0) + 1/4 * ( H7N6E1L2 + H7N6E1L3 ) + 2/4 * ( H7N6E2L1 + H7N6E2L2 ) + 3/4 * ( H7N6E3L1 ) + 4/4 * (0) ) / ( H4N6F1E1 + H7N6E1L2 + H7N6E2L1 + H7N6F1E1L2 + H7N6F1E2L1 + H7N6E1L3 + H7N6E2L2 + H7N6E3L1 + H7N6F1E1L3 + H7N6F1E2L2 ) ) / ( ( 0/4 * (0) + 1/4 * (0) + 2/4 * (0) + 3/4 * (0) + 4/4 * ( H7N6E1L2 + H7N6E2L1 + H7N6E1L3 + H7N6E2L2 + H7N6E3L1 ) ) / ( H4N6F1E1 + H7N6E1L2 + H7N6E2L1 + H7N6F1E1L2 + H7N6F1E2L1 + H7N6E1L3 + H7N6E2L2 + H7N6E3L1 + H7N6F1E1L3 + H7N6F1E2L2 ) ) |
| **A2FGE** | α2,6-sialylation per galactose within fucosylated diantennary glycans | A2FGE = ( ( 0/2 * ( H3N4F1 + H4N4F1 + H3N5F1 + H5N4F1 + H4N5F1 + H5N5F1 + H5N4F1L1 + H5N4F1L2 ) + 1/2 * ( H4N4F1E1 + H5N4F1E1 + H4N5F1E1 + H5N5F1E1 + H5N4F1E1L1 + H5N5F1E1L1 ) + 2/2 * ( H5N4F1E2 + H5N5F1E2 ) ) / ( H3N4F1 + H4N4F1 + H3N5F1 + H5N4F1 + H4N5F1 + H4N4F1E1 + H5N5F1 + H5N4F1L1 + H5N4F1E1 + H4N5F1E1 + H5N5F1E1 + H5N4F1L2 + H5N4F1E1L1 + H5N4F1E2 + H5N5F1E1L1 + H5N5F1E2 ) ) / ( ( 0/2 * ( H3N4F1 + H3N5F1 ) + 1/2 * ( H4N4F1 + H4N5F1 + H4N4F1E1 + H4N5F1E1 ) + 2/2 * ( H5N4F1 + H5N5F1 + H5N4F1L1 + H5N4F1E1 + H5N5F1E1 + H5N4F1L2 + H5N4F1E1L1 + H5N4F1E2 + H5N5F1E1L1 + H5N5F1E2 ) ) / ( H3N4F1 + H4N4F1 + H3N5F1 + H5N4F1 + H4N5F1 + H4N4F1E1 + H5N5F1 + H5N4F1L1 + H5N4F1E1 + H4N5F1E1 + H5N5F1E1 + H5N4F1L2 + H5N4F1E1L1 + H5N4F1E2 + H5N5F1E1L1 + H5N5F1E2 ) ) |
| **A3FGE** | α2,6-sialylation per galactose within fucosylated triantennary glycans | A3FGE = ( ( 0/3 * ( H6N5F1L2 ) + 1/3 * ( H6N5F1E1 + H6N5F1E1L1 + H6N5F1E1L2 ) + 2/3 * ( H6N5F1E2 + H6N5F1E2L1 + H6N5F2E2L1 ) + 3/3 * ( H6N5F1E3 ) ) / ( H6N5F1E1 + H6N5F1L2 + H6N5F1E1L1 + H6N5F1E2 + H6N5F1E1L2 + H6N5F1E2L1 + H6N5F1E3 + H6N5F2E2L1 ) ) / ( ( 0/3 * (0) + 1/3 * |

| | | |
|---|---|---|
| | | (0) + 2/3 * (0) + 3/3 * ( H6N5F1E1 + H6N5F1L2 + H6N5F1E1L1 + H6N5F1E2 + H6N5F1E1L2 + H6N5F1E2L1 + H6N5F1E3 + H6N5F2E2L1 ) ) / ( H6N5F1E1 + H6N5F1L2 + H6N5F1E1L1 + H6N5F1E2 + H6N5F1E1L2 + H6N5F1E2L1 + H6N5F1E3 + H6N5F2E2L1 ) ) |
| **A4FGE** | α2,6-sialylation per galactose within fucosylated tetra-antennary glycans | A4FGE = ( ( 0/4 * (0) + 1/4 * ( H4N6F1E1 + H7N6F1E1L2 + H7N6F1E1L3 ) + 2/4 * ( H7N6F1E2L1 + H7N6F1E2L2 ) + 3/4 * (0) + 4/4 * (0) ) / ( H4N6F1E1 + H7N6E1L2 + H7N6E2L1 + H7N6F1E1L2 + H7N6F1E2L1 + H7N6E1L3 + H7N6E2L2 + H7N6E3L1 + H7N6F1E1L3 + H7N6F1E2L2 ) ) / ( ( 0/4 * (0) + 1/4 * ( H4N6F1E1 ) + 2/4 * (0) + 3/4 * (0) + 4/4 * ( H7N6F1E1L2 + H7N6F1E2L1 + H7N6F1E1L3 + H7N6F1E2L2 ) ) / ( H4N6F1E1 + H7N6E1L2 + H7N6E2L1 + H7N6F1E1L2 + H7N6F1E2L1 + H7N6E1L3 + H7N6E2L2 + H7N6E3L1 + H7N6F1E1L3 + H7N6F1E2L2 ) ) |

**Table S3. Data quality control.** The data quality was checked by measurement of technical replicates of a standard serum sample that were randomly distributed on the plates assessed by calculating the mean, SD (standard deviation), and the relative SD (RSD) for all glycan traits (including directly detected glycan traits and derived glycan traits).

| Derectly detected glycan traits | Average (Relative abundance) | SD | RSD (mean value is 8.35% for all direct traits) | | Derived glycan traits | Average (Relative abundance) | SD | RSD (mean value is 4.06% for all derived traits) |
|---|---|---|---|---|---|---|---|---|
| H5N2 | 0.3836 | 0.0230 | 5.99% | | TM | 0.0203 | 0.0011 | 5.46% |
| H3N3F1 | 0.0761 | 0.0210 | 27.54% | | THy | 0.0098 | 0.0007 | 7.49% |
| H3N4 | 0.1905 | 0.0176 | 9.24% | | TC | 0.9686 | 0.0013 | 0.13% |
| H6N2 | 0.6295 | 0.0250 | 3.97% | | MHy | 2.0621 | 0.0944 | 4.58% |
| H4N3F1 | 0.0697 | 0.0079 | 11.34% | | MM | 6.9643 | 0.0497 | 0.71% |
| H3N3E1 | 0.1407 | 0.0115 | 8.20% | | CA1 | 0.0070 | 0.0003 | 5.03% |
| H5N3 | 0.1067 | 0.0165 | 15.47% | | CA2 | 0.8374 | 0.0054 | 0.64% |
| H3N4F1 | 2.2242 | 0.1430 | 6.43% | | CA3 | 0.1269 | 0.0059 | 4.63% |
| H4N4 | 0.3056 | 0.0120 | 3.91% | | CA4 | 0.0209 | 0.0012 | 5.63% |
| H3N5 | 0.1748 | 0.0237 | 13.57% | | CF | 0.3695 | 0.0078 | 2.10% |
| H7N2 | 0.1841 | 0.0171 | 9.30% | | CFa | 0.0012 | 0.0001 | 6.06% |
| H3N3F1E1 | 0.0695 | 0.0084 | 12.13% | | CB | 0.1121 | 0.0024 | 2.18% |
| H5N3F1 | 0.0478 | 0.0075 | 15.63% | | CG | 0.9635 | 0.0026 | 0.27% |
| H4N3E1 | 0.3670 | 0.0177 | 4.82% | | CS | 0.8587 | 0.0042 | 0.49% |
| H6N3 | 0.0785 | 0.0041 | 5.18% | | TA2FS0 | 12.1743 | 0.3198 | 2.63% |
| H4N4F1 | 4.4296 | 0.1428 | 3.22% | | A1F0 | 0.7494 | 0.0159 | 2.12% |
| H5N4 | 0.3472 | 0.0120 | 3.44% | | A2F0 | 0.6375 | 0.0054 | 0.84% |
| H3N5F1 | 0.8342 | 0.0835 | 10.01% | | A3F0 | 0.5785 | 0.0114 | 1.97% |

| | | | | | | | | |
|---|---|---|---|---|---|---|---|---|
| H4N5 | 0.2138 | 0.0105 | 4.91% | | A4F0 | 0.4281 | 0.1027 | 23.99% |
| H5N3L1 | 0.0427 | 0.0062 | 14.42% | | A1F | 0.2510 | 0.0133 | 5.28% |
| H8N2 | 0.3525 | 0.0182 | 5.18% | | A2F | 0.3605 | 0.0061 | 1.70% |
| H4N3F1E1 | 0.1004 | 0.0089 | 8.88% | | A3F | 0.4180 | 0.0120 | 2.87% |
| H4N4L1 | 0.0392 | 0.0102 | 26.07% | | A4F | 0.5652 | 0.1022 | 18.08% |
| H5N3E1 | 0.2663 | 0.0167 | 6.25% | | A3Fa | 0.0094 | 0.0003 | 3.15% |
| H5N4F1 | 2.6586 | 0.2309 | 8.69% | | A2S0F | 0.8972 | 0.0084 | 0.94% |
| H4N4E1 | 0.4194 | 0.0147 | 3.51% | | A1L0F | 0.2510 | 0.0133 | 5.28% |
| H6N4 | 0.0269 | 0.0066 | 24.57% | | A2L0F | 0.3548 | 0.0061 | 1.71% |
| H4N5F1 | 1.4819 | 0.0389 | 2.62% | | A3L0F | 0.2187 | 0.0045 | 2.06% |
| H5N5 | 0.1690 | 0.0119 | 7.07% | | A2E0F | 0.8693 | 0.0025 | 0.29% |
| H9N2 | 0.4851 | 0.0479 | 9.88% | | A3E0F | 0.5767 | 0.0227 | 3.93% |
| H5N4L1 | 0.2649 | 0.0113 | 4.27% | | A1SF | 0.2510 | 0.0133 | 5.28% |
| H6N3E1 | 0.2210 | 0.0108 | 4.89% | | A2SF | 0.2527 | 0.0050 | 1.97% |
| H4N4F1E1 | 0.6917 | 0.0250 | 3.62% | | A3SF | 0.4180 | 0.0120 | 2.87% |
| H5N4E1 | 4.7762 | 0.1267 | 2.65% | | A4SF | 0.5652 | 0.1022 | 18.08% |
| H5N5F1 | 0.5731 | 0.0225 | 3.93% | | A2LF | 0.4096 | 0.0169 | 4.14% |
| H4N5E1 | 0.3046 | 0.0142 | 4.66% | | A3LF | 0.5071 | 0.0202 | 3.97% |
| H5N4F1L1 | 0.5665 | 0.0097 | 1.71% | | A4LF | 0.3820 | 0.0199 | 5.22% |
| H6N4L1 | 0.0828 | 0.0067 | 8.13% | | A1EF | 0.2510 | 0.0133 | 5.28% |
| H4N7 | 0.0839 | 0.0142 | 16.96% | | A2EF | 0.2370 | 0.0050 | 2.10% |
| H5N4F1E1 | 3.3589 | 0.2284 | 6.80% | | A3EF | 0.4147 | 0.0120 | 2.89% |
| H4N5F1E1 | 0.3828 | 0.0178 | 4.65% | | A4EF | 0.5652 | 0.1022 | 18.08% |
| H5N5E1 | 1.0621 | 0.0362 | 3.41% | | A2B | 0.1336 | 0.0032 | 2.37% |
| H5N4L2 | 0.4238 | 0.0248 | 5.85% | | A2F0B | 0.0427 | 0.0019 | 4.37% |

| | | | | | | | | |
|---|---|---|---|---|---|---|---|---|
| H5N4E1L1 | 4.1897 | 0.1461 | 3.49% | | A2FB | 0.2915 | 0.0062 | 2.12% |
| H5N4E2 | 38.6230 | 0.5938 | 1.54% | | A2S0B | 0.2506 | 0.0063 | 2.51% |
| H5N5F1E1 | 2.1881 | 0.0777 | 3.55% | | A2SB | 0.1097 | 0.0026 | 2.35% |
| H6N5E1 | 0.5117 | 0.0306 | 5.98% | | A2F0S0B | 0.3999 | 0.0135 | 3.39% |
| H5N4F1L2 | 1.0980 | 0.1301 | 11.85% | | A2F0SB | 0.0331 | 0.0014 | 4.18% |
| H4N6F1E1 | 0.6992 | 0.1501 | 21.47% | | A2FS0B | 0.2344 | 0.0072 | 3.08% |
| H5N4F1E1L1 | 1.6740 | 0.0527 | 3.15% | | A2FSB | 0.3335 | 0.0081 | 2.44% |
| H4N7E1 | 0.6067 | 0.0295 | 4.86% | | A2G | 0.9070 | 0.0020 | 0.22% |
| H5N4F1E2 | 4.0093 | 0.0615 | 1.53% | | A4G | 0.7532 | 0.0749 | 9.95% |
| H6N5F1E1 | 0.2404 | 0.0132 | 5.47% | | A2F0G | 0.9805 | 0.0006 | 0.07% |
| H5N5E2 | 0.3038 | 0.0126 | 4.15% | | A4F0G | 0.4281 | 0.1027 | 23.99% |
| H6N5L2 | 0.0993 | 0.0071 | 7.10% | | A2FG | 0.7759 | 0.0081 | 1.04% |
| H5N5F1E1L1 | 0.1380 | 0.0068 | 4.95% | | A4FG | 0.3378 | 0.0069 | 2.03% |
| H6N5E1L1 | 0.5685 | 0.0433 | 7.62% | | A2S0G | 0.5085 | 0.0216 | 4.25% |
| H5N5F1E2 | 3.0111 | 0.0562 | 1.87% | | A2SG | 0.9863 | 0.0005 | 0.05% |
| H6N5E2 | 0.5099 | 0.0298 | 5.84% | | A2F0S0G | 0.5544 | 0.0153 | 2.76% |
| H6N5F1L2 | 0.1325 | 0.0120 | 9.04% | | A2FS0G | 0.5044 | 0.0204 | 4.05% |
| H6N5F1E1L1 | 0.3050 | 0.0198 | 6.50% | | A2F0SG | 0.9924 | 0.0005 | 0.05% |
| H6N5F1E2 | 0.3750 | 0.0288 | 7.69% | | A2FSG | 0.9684 | 0.0008 | 0.09% |
| H8N6F2 | 0.1075 | 0.0117 | 10.87% | | A2S | 0.7451 | 0.0048 | 0.64% |
| H6N5E1L2 | 0.5549 | 0.0746 | 13.44% | | A3S | 0.9039 | 0.0021 | 0.23% |
| H6N5E2L1 | 2.9630 | 0.3386 | 11.43% | | A4S | 0.6840 | 0.0647 | 9.46% |
| H6N5E3 | 1.9973 | 0.1442 | 7.22% | | A2F0S | 0.9097 | 0.0064 | 0.71% |
| H6N5F1E1L2 | 0.4405 | 0.0198 | 4.50% | | A3F0S | 0.8958 | 0.0036 | 0.41% |
| H6N5F1E2L1 | 3.2778 | 0.0601 | 1.83% | | A4F0S | 0.3833 | 0.0926 | 24.16% |

| | | | | | | | | |
|---|---|---|---|---|---|---|---|---|
| H6N5F1E3 | 0.2212 | 0.0185 | 8.36% | | A2FS | 0.4600 | 0.0046 | 1.00% |
| H6N5F2E2L1 | 0.1152 | 0.0069 | 5.95% | | A3FS | 0.9170 | 0.0051 | 0.56% |
| H7N6E1L2 | 0.1863 | 0.0310 | 16.62% | | A4FS | 0.3128 | 0.0062 | 1.98% |
| H7N6E2L1 | 0.1716 | 0.0267 | 15.57% | | A2L | 0.0602 | 0.0005 | 0.79% |
| H7N6F1E1L2 | 0.1181 | 0.0114 | 9.67% | | A3L | 0.2629 | 0.0043 | 1.64% |
| H7N6F1E2L1 | 0.0845 | 0.0090 | 10.69% | | A4L | 0.3275 | 0.0562 | 17.14% |
| H7N6E1L3 | 0.1556 | 0.0314 | 20.17% | | A2F0L | 0.0515 | 0.0016 | 3.15% |
| H7N6E2L2 | 0.2307 | 0.0479 | 20.75% | | A3F0L | 0.2238 | 0.0079 | 3.55% |
| H7N6E3L1 | 0.1090 | 0.0112 | 10.29% | | A4F0L | 0.1986 | 0.0500 | 25.15% |
| H7N6F1E1L3 | 0.1250 | 0.0091 | 7.24% | | A2FL | 0.0749 | 0.0044 | 5.82% |
| H7N6F1E2L2 | 0.1830 | 0.0087 | 4.73% | | A3FL | 0.3171 | 0.0028 | 0.87% |
| | | | | | A4FL | 0.1286 | 0.0067 | 5.18% |
| | | | | | A2E | 0.6849 | 0.0048 | 0.70% |
| | | | | | A3E | 0.6437 | 0.0044 | 0.68% |
| | | | | | A4E | 0.3586 | 0.0147 | 4.10% |
| | | | | | A2F0E | 0.8549 | 0.0056 | 0.65% |
| | | | | | A3F0E | 0.6757 | 0.0038 | 0.56% |
| | | | | | A4F0E | 0.1847 | 0.0428 | 23.17% |
| | | | | | A2FE | 0.3867 | 0.0086 | 2.23% |
| | | | | | A3FE | 0.5994 | 0.0036 | 0.59% |
| | | | | | A4FE | 0.1732 | 0.0261 | 15.08% |
| | | | | | A2GS | 0.8219 | 0.0061 | 0.74% |
| | | | | | A3GS | 0.9039 | 0.0021 | 0.23% |
| | | | | | A4GS | 0.9091 | 0.0035 | 0.38% |
| | | | | | A2F0GS | 0.9280 | 0.0058 | 0.62% |

| | | | | | | | | |
|---|---|---|---|---|---|---|---|---|
| | | | | | A3F0GS | 0.8958 | 0.0036 | 0.41% |
| | | | | | A4F0GS | 0.8947 | 0.0015 | 0.17% |
| | | | | | A2FGS | 0.5919 | 0.0072 | 1.21% |
| | | | | | A3FGS | 0.9170 | 0.0051 | 0.56% |
| | | | | | A4FGS | 0.9261 | 0.0051 | 0.55% |
| | | | | | A2GL | 0.0665 | 0.0006 | 0.96% |
| | | | | | A3GL | 0.2629 | 0.0043 | 1.64% |
| | | | | | A4GL | 0.4311 | 0.0271 | 6.28% |
| | | | | | A2F0GL | 0.0524 | 0.0016 | 3.01% |
| | | | | | A3F0GL | 0.2238 | 0.0079 | 3.55% |
| | | | | | A4F0GL | 0.4635 | 0.0060 | 1.29% |
| | | | | | A2FGL | 0.0967 | 0.0044 | 4.54% |
| | | | | | A3FGL | 0.3171 | 0.0028 | 0.87% |
| | | | | | A4FGL | 0.3904 | 0.0409 | 10.49% |
| | | | | | A2GE | 0.7554 | 0.0060 | 0.79% |
| | | | | | A3GE | 0.6437 | 0.0044 | 0.68% |
| | | | | | A4GE | 0.4819 | 0.0176 | 3.65% |
| | | | | | A2F0GE | 0.8720 | 0.0052 | 0.59% |
| | | | | | A3F0GE | 0.6757 | 0.0038 | 0.56% |
| | | | | | A4F0GE | 0.4308 | 0.0054 | 1.25% |
| | | | | | A2FGE | 0.4972 | 0.0122 | 2.46% |
| | | | | | A3FGE | 0.5994 | 0.0036 | 0.59% |
| | | | | | A4FGE | 0.5388 | 0.0266 | 4.94% |

**Table S4. Associations of serum N-glycans with EC (complete list of tests performed).** The associations were determined with logistic regression. Age was included as a covariate in the models for the disease-related tests. Odds ratios (OR) are calculated on scaled data. The p-values considered significant with the significance threshold of 4.31E-04 (=0.05/116 derived glycan traits) and indicated in bold in the table. The p-values and OR are reported for the derived glycan traits. HC, healthy controls; EC, endometrial cancer.

| Derived glycan traits | beta | SEM | T | p (logistics regression) | OR | p (U test) |
|---|---|---|---|---|---|---|
| TM | 0.86 | 0.32 | 2.65 | 8.01E-03 | 2.36 | 3.51E-03 |
| **THy** | **-3.51** | **0.86** | **-4.08** | **4.48E-05** | **0.03** | **1.37E-09** |
| TC | 0.24 | 0.27 | 0.88 | 3.78E-01 | 1.27 | 3.26E-01 |
| **MHy** | **2.93** | **0.67** | **4.38** | **1.20E-05** | **18.82** | **4.29E-10** |
| MM | 0.41 | 0.28 | 1.43 | 1.51E-01 | 1.50 | 8.16E-02 |
| CA1 | 0.93 | 0.35 | 2.68 | 7.47E-03 | 2.54 | **6.37E-05** |
| CA2 | -0.74 | 0.31 | -2.37 | 1.76E-02 | 0.48 | 6.71E-03 |
| CA3 | 1.28 | 0.37 | 3.41 | 6.38E-04 | 3.58 | **6.05E-05** |
| **CA4** | **-1.46** | **0.41** | **-3.59** | **3.32E-04** | **0.23** | **1.41E-05** |
| **CF** | **-2.50** | **0.60** | **-4.19** | **2.80E-05** | **0.08** | **6.10E-09** |
| CFa | -0.55 | 0.29 | -1.91 | 5.63E-02 | 0.58 | 1.58E-01 |
| CB | -0.97 | 0.32 | -3.04 | 2.36E-03 | 0.38 | 1.20E-03 |
| CG | 0.38 | 0.27 | 1.40 | 1.62E-01 | 1.47 | 2.59E-01 |
| CS | 1.27 | 0.37 | 3.43 | 6.07E-04 | 3.57 | **2.88E-05** |
| TA2FS0 | -1.27 | 0.37 | -3.46 | 5.50E-04 | 0.28 | **2.32E-05** |
| **A1F0** | **1.63** | **0.44** | **3.73** | **1.95E-04** | **5.12** | **2.33E-06** |
| **A2F0** | **2.12** | **0.51** | **4.12** | **3.72E-05** | **8.33** | **6.78E-08** |
| A3F0 | 0.62 | 0.29 | 2.11 | 3.45E-02 | 1.86 | 4.43E-02 |

| | | | | | | |
|---|---|---|---|---|---|---|
| A4F0 | 9.76 | 4.19 | 2.33 | 1.99E-02 | >100 | **2.09E-12** |
| **A1F** | **-1.63** | **0.44** | **-3.73** | **1.94E-04** | **0.20** | **2.33E-06** |
| **A2F** | **-2.12** | **0.51** | **-4.12** | **3.73E-05** | **0.12** | **6.78E-08** |
| A3F | -0.62 | 0.29 | -2.12 | 3.43E-02 | 0.54 | 4.43E-02 |
| A4F | -9.79 | 4.20 | -2.33 | 1.98E-02 | 0.00 | **2.09E-12** |
| A3Fa | -1.02 | 0.35 | -2.94 | 3.25E-03 | 0.36 | 1.11E-02 |
| A2S0F | -0.83 | 0.33 | -2.52 | 1.18E-02 | 0.43 | 2.76E-03 |
| **A1L0F** | **-1.63** | **0.44** | **-3.73** | **1.94E-04** | **0.20** | **2.33E-06** |
| **A2L0F** | **-2.15** | **0.52** | **-4.11** | **4.02E-05** | **0.12** | **6.78E-08** |
| **A3L0F** | **-2.47** | **0.59** | **-4.22** | **2.44E-05** | **0.08** | **1.36E-08** |
| A2E0F | -0.06 | 0.25 | -0.23 | 8.21E-01 | 0.95 | 9.51E-01 |
| **A3E0F** | **-1.84** | **0.46** | **-4.01** | **6.06E-05** | **0.16** | **4.63E-07** |
| **A1SF** | **-1.63** | **0.44** | **-3.73** | **1.94E-04** | **0.20** | **2.33E-06** |
| **A2SF** | **-2.08** | **0.50** | **-4.13** | **3.65E-05** | **0.12** | **7.77E-08** |
| A3SF | -0.62 | 0.29 | -2.12 | 3.43E-02 | 0.54 | 4.43E-02 |
| A4SF | -9.79 | 4.20 | -2.33 | 1.98E-02 | 0.00 | **2.09E-12** |
| A2LF | -0.79 | 0.32 | -2.49 | 1.29E-02 | 0.46 | 4.69E-02 |
| A3LF | -0.39 | 0.28 | -1.43 | 1.53E-01 | 0.67 | 3.15E-01 |
| A4LF | -0.25 | 0.26 | -0.95 | 3.44E-01 | 0.78 | 4.54E-01 |
| **A1EF** | **-1.63** | **0.44** | **-3.73** | **1.94E-04** | **0.20** | **2.33E-06** |
| **A2EF** | **-2.01** | **0.49** | **-4.10** | **4.06E-05** | **0.13** | **1.09E-07** |
| A3EF | -0.51 | 0.28 | -1.79 | 7.35E-02 | 0.60 | 1.38E-01 |
| A4EF | -9.79 | 4.20 | -2.33 | 1.98E-02 | 0.00 | **2.09E-12** |
| A2B | -0.91 | 0.31 | -2.95 | 3.17E-03 | 0.40 | 1.69E-03 |
| A2F0B | 0.36 | 0.29 | 1.23 | 2.20E-01 | 1.43 | 3.51E-01 |

| | | | | | | |
|---|---|---|---|---|---|---|
| A2FB | 0.03 | 0.26 | 0.11 | 9.14E-01 | 1.03 | 4.77E-01 |
| A2S0B | 0.29 | 0.27 | 1.08 | 2.78E-01 | 1.34 | 1.00E-01 |
| A2SB | -0.83 | 0.30 | -2.74 | 6.14E-03 | 0.43 | 3.24E-03 |
| A2F0S0B | 1.02 | 0.38 | 2.67 | 7.68E-03 | 2.78 | 2.00E-03 |
| A2F0SB | 0.40 | 0.31 | 1.31 | 1.89E-01 | 1.50 | 3.45E-01 |
| A2FS0B | 0.02 | 0.26 | 0.09 | 9.31E-01 | 1.02 | 4.40E-01 |
| A2FSB | 0.02 | 0.25 | 0.09 | 9.29E-01 | 1.02 | 5.81E-01 |
| A2G | 0.78 | 0.30 | 2.58 | 1.00E-02 | 2.19 | 9.31E-03 |
| A4G | 265.34 | 80261.81 | 0.00 | 9.97E-01 | >100 | **1.75E-12** |
| A2F0G | 1.57 | 0.46 | 3.40 | 6.82E-04 | 4.79 | **3.39E-05** |
| A4F0G | 9.76 | 4.19 | 2.33 | 1.99E-02 | >100 | **2.09E-12** |
| A2FG | -0.04 | 0.26 | -0.16 | 8.75E-01 | 0.96 | 4.54E-01 |
| A4FG | -0.29 | 0.28 | -1.05 | 2.92E-01 | 0.75 | 3.82E-02 |
| A2S0G | -0.45 | 0.31 | -1.47 | 1.41E-01 | 0.64 | 3.38E-02 |
| **A2SG** | **3.98** | **0.97** | **4.09** | **4.34E-05** | **53.34** | **6.79E-11** |
| A2F0S0G | -0.13 | 0.28 | -0.47 | 6.37E-01 | 0.88 | 2.80E-01 |
| A2FS0G | -0.51 | 0.31 | -1.63 | 1.04E-01 | 0.60 | 1.98E-02 |
| **A2F0SG** | **4.39** | **1.15** | **3.81** | **1.42E-04** | **81.02** | **3.13E-10** |
| A2FSG | 0.87 | 0.30 | 2.87 | 4.17E-03 | 2.38 | 1.15E-03 |
| **A2S** | **1.66** | **0.44** | **3.74** | **1.84E-04** | **5.28** | **1.62E-06** |
| **A3S** | **2.77** | **0.69** | **4.00** | **6.23E-05** | **15.97** | **1.68E-08** |
| A4S | 259.70 | 79698.04 | 0.00 | 9.97E-01 | >100 | **1.92E-12** |
| A2F0S | 0.40 | 0.26 | 1.50 | 1.32E-01 | 1.49 | 4.05E-02 |
| **A3F0S** | **1.64** | **0.46** | **3.57** | **3.56E-04** | **5.17** | **2.73E-05** |
| A4F0S | 9.60 | 4.10 | 2.34 | 1.92E-02 | >100 | **2.49E-12** |

| | | | | | | |
|---|---|---|---|---|---|---|
| A2FS | 0.49 | 0.28 | 1.72 | 8.46E-02 | 1.63 | 7.53E-02 |
| A3FS | 1.58 | 0.50 | 3.19 | 1.41E-03 | 4.88 | **7.44E-05** |
| A4FS | -0.49 | 0.30 | -1.66 | 9.69E-02 | 0.61 | 8.99E-03 |
| A2L | -0.85 | 0.31 | -2.77 | 5.54E-03 | 0.43 | 1.10E-03 |
| A3L | -0.66 | 0.30 | -2.24 | 2.54E-02 | 0.51 | 2.88E-03 |
| A4L | 624.88 | 101446.81 | 0.01 | 9.95E-01 | >100 | **1.92E-12** |
| **A2F0L** | **-2.15** | **0.54** | **-3.95** | **7.83E-05** | **0.12** | **6.33E-08** |
| A3F0L | -0.31 | 0.28 | -1.12 | 2.63E-01 | 0.73 | 4.69E-02 |
| A4F0L | 8.63 | 3.42 | 2.53 | 1.15E-02 | >100 | **5.00E-12** |
| A2FL | 0.54 | 0.29 | 1.87 | 6.21E-02 | 1.72 | 2.11E-02 |
| A3FL | -0.22 | 0.26 | -0.84 | 4.03E-01 | 0.81 | 3.64E-01 |
| **A4FL** | **4.08** | **1.06** | **3.86** | **1.14E-04** | **59.22** | **1.72E-09** |
| **A2E** | **2.07** | **0.52** | **3.96** | **7.38E-05** | **7.91** | **1.17E-07** |
| **A3E** | **3.46** | **0.91** | **3.81** | **1.39E-04** | **31.67** | **3.38E-09** |
| A4E | 17.75 | 11.87 | 1.49 | 1.35E-01 | >100 | **1.61E-12** |
| A2F0E | 1.00 | 0.32 | 3.08 | 2.05E-03 | 2.71 | **2.34E-04** |
| A3F0E | 1.72 | 0.49 | 3.49 | 4.77E-04 | 5.59 | **8.72E-07** |
| A4F0E | 7.96 | 2.89 | 2.75 | 5.88E-03 | >100 | **2.09E-12** |
| A2FE | 0.41 | 0.27 | 1.51 | 1.32E-01 | 1.51 | 1.19E-01 |
| **A3FE** | **3.46** | **0.92** | **3.74** | **1.87E-04** | **31.66** | **7.07E-09** |
| **A4FE** | **-3.82** | **0.95** | **-4.04** | **5.40E-05** | **0.02** | **1.11E-10** |
| **A2GS** | **2.01** | **0.54** | **3.73** | **1.88E-04** | **7.46** | **9.28E-07** |
| **A3GS** | **2.77** | **0.69** | **4.00** | **6.23E-05** | **15.97** | **1.68E-08** |
| **A4GS** | **-4.50** | **1.19** | **-3.80** | **1.45E-04** | **0.01** | **9.94E-12** |
| A2F0GS | 0.10 | 0.25 | 0.40 | 6.91E-01 | 1.10 | 2.39E-01 |

| | | | | | | |
|---|---|---|---|---|---|---|
| **A3F0GS** | **1.64** | **0.46** | **3.57** | **3.56E-04** | **5.17** | **2.73E-05** |
| A4F0GS | 0.12 | 0.26 | 0.48 | 6.32E-01 | 1.13 | 9.80E-01 |
| A2FGS | 0.92 | 0.36 | 2.58 | 9.98E-03 | 2.51 | 1.20E-03 |
| A3FGS | 1.58 | 0.50 | 3.19 | 1.41E-03 | 4.88 | **7.44E-05** |
| **A4FGS** | **-4.74** | **1.24** | **-3.83** | **1.27E-04** | **0.01** | **8.67E-11** |
| A2GL | -1.23 | 0.36 | -3.43 | 6.09E-04 | 0.29 | **5.17E-05** |
| A3GL | -0.66 | 0.30 | -2.24 | 2.54E-02 | 0.51 | 2.88E-03 |
| A4GL | 10.36 | 4.31 | 2.41 | 1.61E-02 | >100 | **5.94E-12** |
| **A2F0GL** | **-2.22** | **0.56** | **-3.95** | **7.69E-05** | **0.11** | **5.52E-08** |
| A3F0GL | -0.31 | 0.28 | -1.12 | 2.63E-01 | 0.73 | 4.69E-02 |
| A4F0GL | -0.11 | 0.27 | -0.43 | 6.69E-01 | 0.89 | 3.15E-01 |
| A2FGL | 0.54 | 0.29 | 1.85 | 6.48E-02 | 1.72 | 1.23E-02 |
| A3FGL | -0.22 | 0.26 | -0.84 | 4.03E-01 | 0.81 | 3.64E-01 |
| A4FGL | 9.43 | 4.01 | 2.35 | 1.88E-02 | >100 | **5.45E-12** |
| **A2GE** | **2.98** | **0.77** | **3.86** | **1.13E-04** | **19.66** | **3.91E-08** |
| **A3GE** | **3.46** | **0.91** | **3.81** | **1.39E-04** | **31.67** | **3.38E-09** |
| A4GE | -1779.02 | 152685.85 | -0.01 | 9.91E-01 | 0.00 | **1.92E-12** |
| A2F0GE | 0.82 | 0.30 | 2.73 | 6.43E-03 | 2.27 | 1.37E-03 |
| A3F0GE | 1.72 | 0.49 | 3.49 | 4.77E-04 | 5.59 | **8.72E-07** |
| A4F0GE | 0.18 | 0.26 | 0.70 | 4.84E-01 | 1.20 | 2.29E-01 |
| A2FGE | 0.74 | 0.32 | 2.32 | 2.02E-02 | 2.09 | 2.55E-03 |
| **A3FGE** | **3.46** | **0.92** | **3.74** | **1.87E-04** | **31.66** | **7.07E-09** |
| A4FGE | -10.05 | 4.68 | -2.15 | 3.16E-02 | 0.00 | **5.45E-12** |

**Table S5. The diagnostic value of the optimized glycan panel built with the four most important (discriminative and biologically reliable) derived glycan traits using different models/algorithms.**

| Model/Algorithm (used in the multivariate ROC exploratory analyses) | AUC | 95%CI | Average accuracy |
|---|---|---|---|
| Random forest | 0.993 | 0.955-1 | 0.959 |
| Linear SVM | 0.987 | 0.946-1 | 0.940 |
| PLS | 0.989 | 0.950-1 | 0.957 |

**Table S6. Associations of serum N-glycan traits and classical gynecologic tumor markers with the differentiation type (well or poorly differentiated) of EC.** The associations were determined with logistic regression. Age was included as a covariate in the models. Odds ratios (OR) are calculated on scaled data.

| Derived glycan traits or classical gynecologic tumor markers | beta | SEM | T | p (logistics regression) | OR |
|---|---|---|---|---|---|
| **Derived glycan traits** | | | | | |
| THy | -2.24 | 0.95 | -2.36 | **0.018** | 0.11 |
| TC | 1.58 | 0.75 | 2.10 | **0.035** | 4.86 |
| **Classical gynecologic tumor markers** | | | | | |
| CA125 | 0.82 | 0.63 | 1.30 | 0.194 | / |
| CA199 | -0.67 | 0.93 | -0.73 | 0.468 | / |
| CEA | -0.14 | 0.63 | -0.22 | 0.828 | / |

### Supplementary Figures

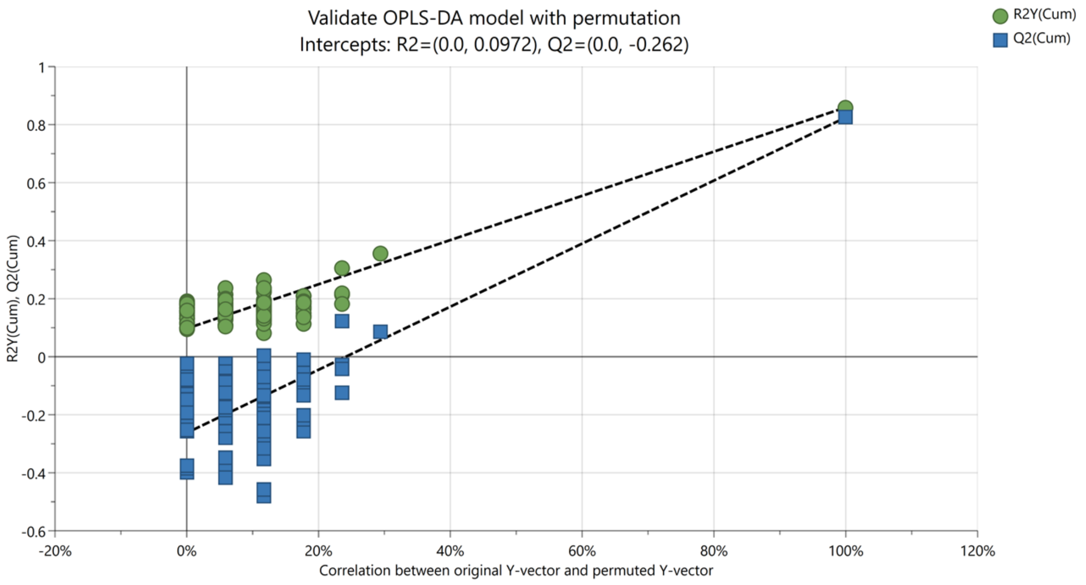


**Figure S1. The OPLS-DA model's permutation (cross-validation) test.** Permutation number = 200. OPLS-DA, orthogonal partial least squares - discriminant analysis.

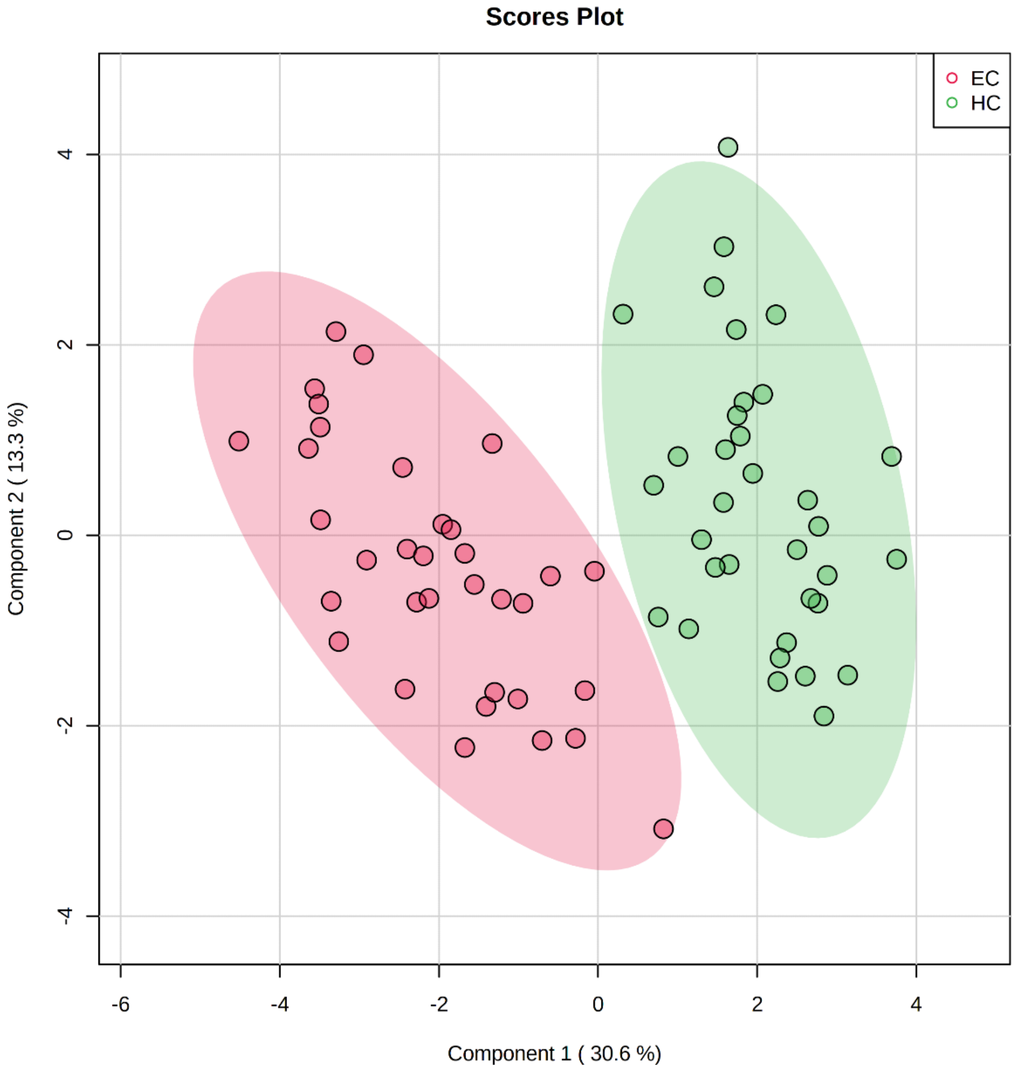


**Figure S2. Scores plot of the sparse partial least squares - discriminant analysis (sPLS-DA) between endometrial cancer (EC, red circle) and healthy controls (HC, green circle) groups.** The scores plot is based on derived glycan traits. Areas of 95% confidence interval are highlighted in red and green for EC and HC, respectively.

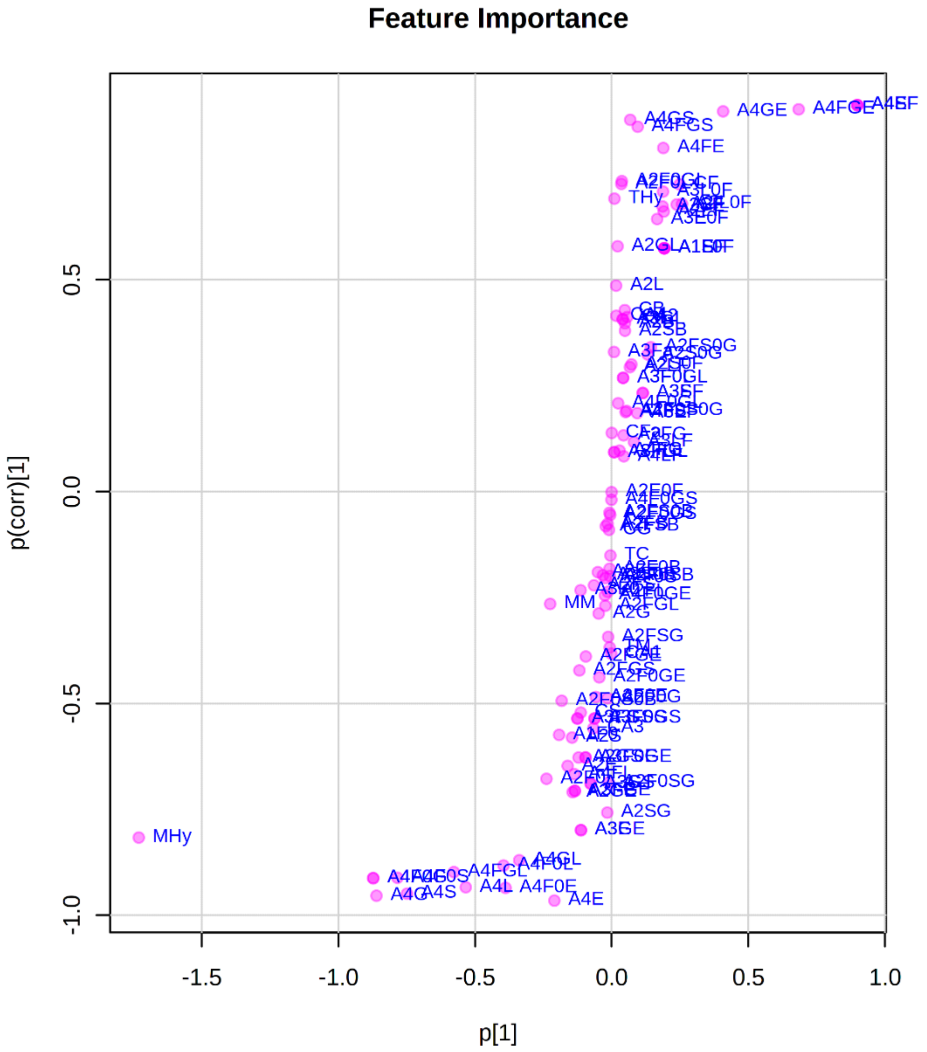


**Figure S3. The S-plot of the orthogonal partial least squares - discriminant analysis (OPLS-DA).**

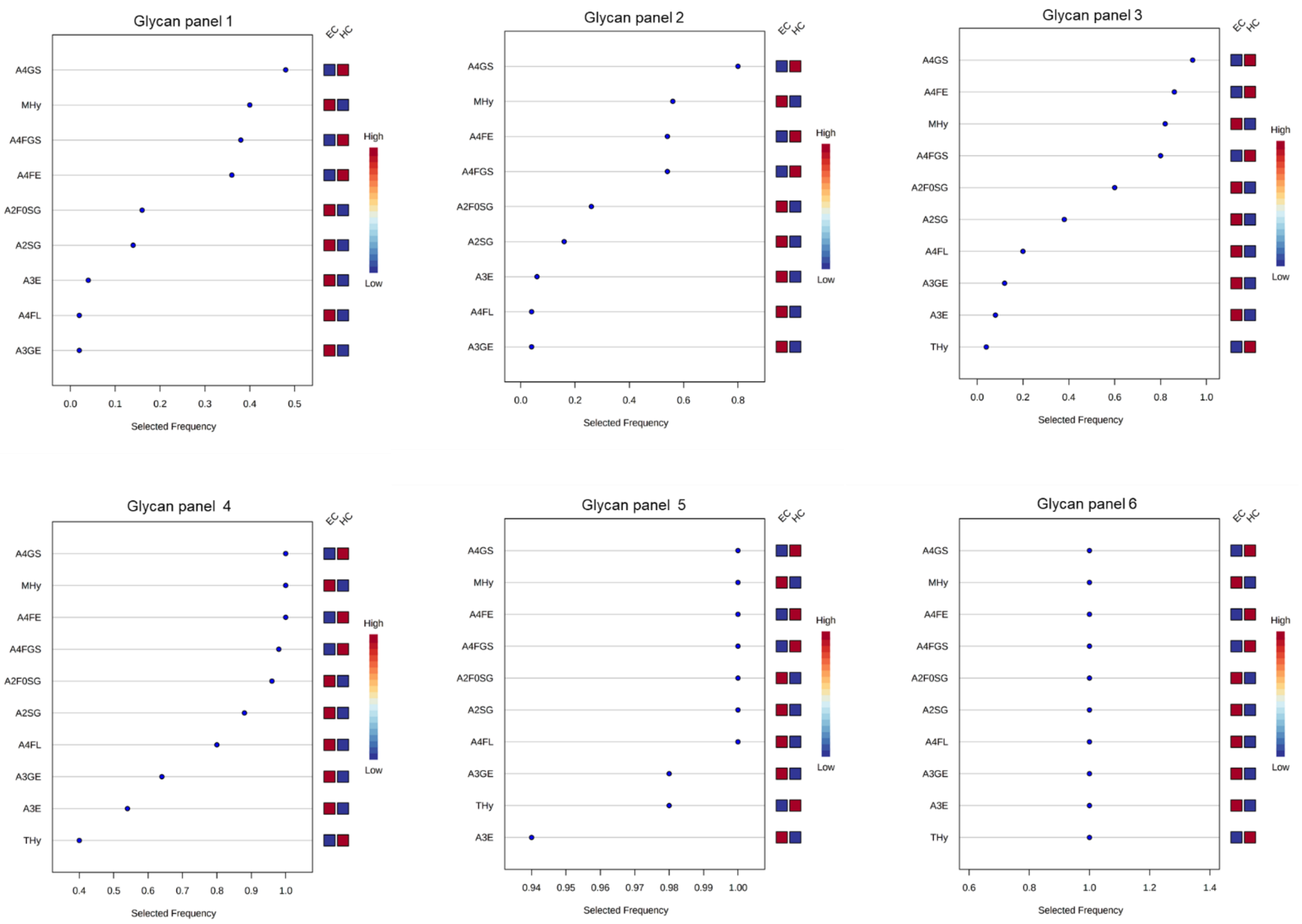


**Figure S4. The most frequently selected derived glycan traits when building the glycan panels (panel 1-panel 6, built with 2, 3, 5, 10, 20, or 33 derived glycan traits).** They were selected and evaluated by the multivariate receiver operating characteristic (ROC) exploratory analyses.

The exploratory analysis didn’t point out which specific derived glycan traits were used to construct each glycan panel, but it indicated the most frequently selected derived glycan traits when building the panels, which facilitate the building of the optimal glycan panel.

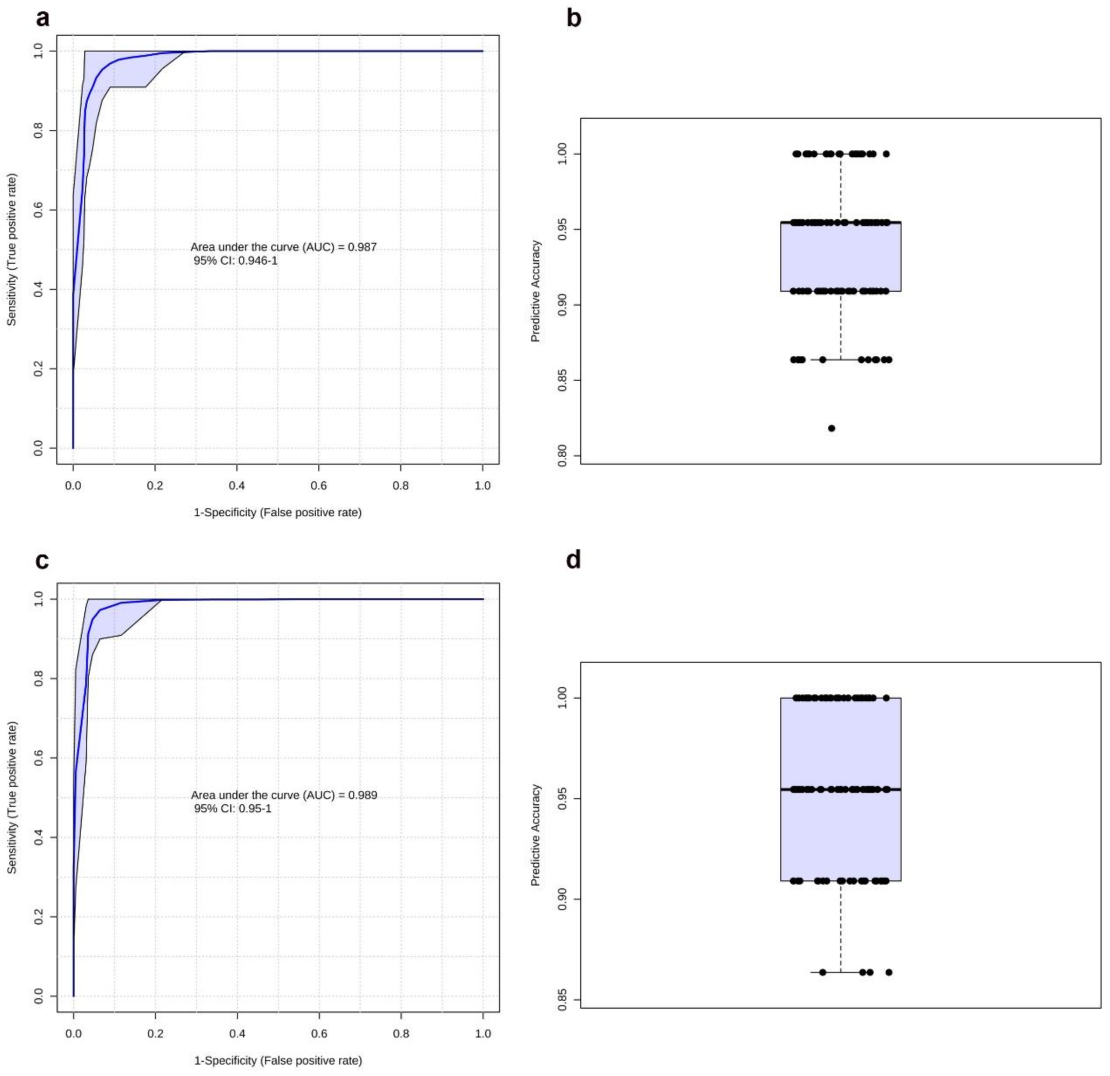


**Figure S5. Diagnostic performance of the optimized glycan panel built with the four most important (discriminative and biologically reliable) glycan traits based on the support vector machines (SVM) model/algorithm and partial least squares (PLS) model/algorithm for endometrial cancer. (A)** receiver operating characteristic (ROC) curve and **(B)** the predictive accuracy based on the linear SVM model/algorithm. **(C)** ROC curve and **(D)** the predictive accuracy based on the PLS model/algorithm.

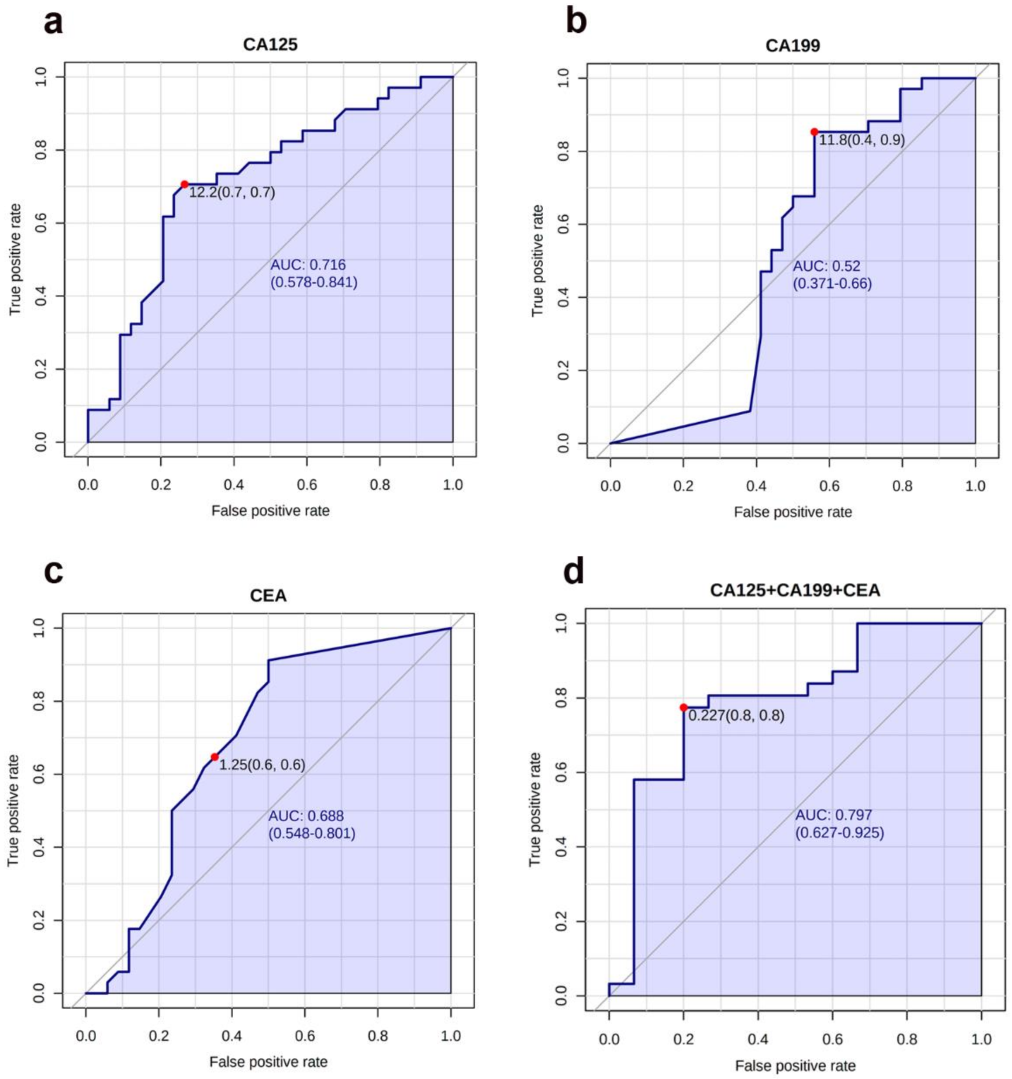


**Figure S6. Diagnostic performance of the classical gynecologic tumor markers for endometrial cancer.** The diagnostic performance of **(A)** carbohydrate antigen 125 [CA125], **(B)** CA199, **(C)** carcinoembryonic antigen [CEA], and **(D)** the combination of the three markers.
